# Clinical, *in vitro,* and *in vivo* evidence of *WAPL* as a novel cohesinopathy gene and phenotypic driver of 10q22.3q23.2 genomic disorder

**DOI:** 10.64898/2026.02.23.26346364

**Authors:** Philip M. Boone, Serkan Erdin, Abucar Mohamed, Sadegheh Haghshenas, Kamli N. W. Faour, Emeline Kao, Jack Fu, Chiara Auwerx, Ricardo Harripaul, Bimal Jana, Danielle Springer, Grey Hallstrom, Celine E.F. de Esch, Erica Denhoff, Lauren Holmes, Kiana Mohajeri, John Lemanski, Jennifer Kerkhof, Haley McConkey, Jessica Rzasa, Madison J. McCune, Michael A. Levy, Julia Grafstein, Matthew Larson, Zsabre Wright, Roberta L Beauchamp, Diane Lucente, Rami Abou Jamra, Neena Agrawal, Pankaj Agrawal, Erica F. Andersen, Emanuela Argilli, Renee Araiza, Sonia Ballal, Megan F. Baxter, Gaber Bergant, Astrid Bertsche, Riya Bhavsar, Debora R. Bortola, Viktoria Bothe, Charlotte Brasch-Andersen, Dominique Braun MPhil, Ange-Line Bruel, Catherine Buchanan, Nicholas D. Burt, Laura M.L. Carvalho, Luigi Chiriatti, Benjamin Cogne, Ryan Collins, Amy Crunk, Benjamin Currall, Andree Delahaye-Duriez, Julian Delanne, Anne-Sophie Denommé-Pichon, Koenraad Devriendt, Aloysius Domingo, Laura Duncan, Laurence Faivre, Laura Famularo, Anne Fulton, Casie Genetti, Tamar Harel, Marketa Havlovicova, Jenny Higgs, Marine Houlier, Maria Iascone, LaDonna Immken, Bertrand Isidor, Frank J. Kaiser, Kaycee Karbone, Margaret Kenna, Amjad Khan, Lara Kristina Kimmig, Tjitske Kleefstra, Eva-Maria Kraus, Ana C.V. Krepischi, Ilona Krey, Roger Ladda, Louise Lanoue, Cedric Le Caignec, Zoe K. Lewis, Gloria Lima, Sally Ann Lynch, Milan Macek, Olivier Maier, Silvia Maitz, Alison Male, Marcela Malikova, Victoria McKay, Oana Moldovan, Danielle Monteil, Mariana Moysés Oliveira, Jeeva Munasinghe, Sachiko Nakamori, Sonja Neuser, Mathilde Nizon, Xander Nuttle, Kathryn O’Keefe, Laura Orec, Ilaria Parenti, Borut Peterlin, Rolph Pfundt, Jill Pouncey, Francesca Clementina Radio, Leema Robert, Lance Rodan, Hallel Rosenberg-Fogler, Jill A. Rosenfeld, Hana Safraou, Monica Salani, Sophia Schliesske, Eleanor G. Seaby, Susan Sell, A. Eliot Shearer, Elliott Sherr, Amelle Shillington, Dorothea Siebold, Margje Sinnema, Laura Smith, Alexander P.A. Stegmann, Cathy Stevens, Servi Stevens, Eric Surette, Marco Tartaglia, Jenny C. Taylor, Michelle L. Thompson, Pernille M. Tørring, Frederic Tran Mau Them, Olga Tsoulaki, Muhammad Umair, Els Vanhoutte, Marie Vincent, Antonio Vitobello, Lydia von Wintzingerode, Amy Watt, Marketa Wayhelova, Ingrid M. Wentzensen, William Wilson, Monica H. Wojcik, Bo Yuan, Giuseppe Zampino, Siddharth Srivastava, Dominik S. Westphal, Korbinian M. Riedhammer, Eric Joyce, Rachita Yadav, James Gusella, Derek J. C. Tai, Bekim Sadikovic, Karl E. Pfeifer, Michael E. Talkowski

## Abstract

Cohesin is a fundamental genome-organizing complex that orchestrates three-dimensional chromosome folding and gene expression via DNA loop extrusion. Alterations to genes encoding cohesin subunits and cohesin loaders cause Mendelian disorders, including Cornelia de Lange syndrome (CdLS). By contrast, disruption of factors that remove cohesin from DNA, including *WAPL* and its binding partners *PDS5A* and *PDS5B*, have not yet been associated with human disease. Here, we explored the relevance of these cohesin release factors in Mendelian disease by establishing a rare disease cohort of deeply phenotyped individuals with heterozygous, predicted damaging variants in *WAPL* (n=27), *PDS5A* (n=8), and *PDS5B* (n=8), by modeling *WAPL* deficiency in human cell lines and mice, and by aggregating rare disease association statistics from consortia studies. We identified a *WAPL*-related disorder characterized by developmental delay, intellectual disability, and risk of other developmental anomalies including clubfoot. Similarities between individuals with damaging *WAPL* variants and those with large, recurrent 10q22.3q23.2 (10q) deletions (which encompass *WAPL*) nominate *WAPL* as a driver gene within this genomic disorder region. While carriers of *PDS5A* or *PDS5B* variants exhibited features of developmental disorders, neither cohort-based statistics nor case phenotyping associated these genes with specific phenotypes. We used CRISPR engineering to generate truncating variants in *WAPL*, as well the 7.8 Mb 10q deletion or duplication in human iPSCs and induced neurons. Transcriptomic analyses identified differentially expressed genes in both models, with highly significant overlap between *WAPL* haploinsufficiency and 10q deletion signatures. Mice with 50% residual *Wapl* expression exhibited mild deficits of growth and learning/memory, whereas those with 25% residual *Wapl* expression displayed birth defects and postnatal lethality, revealing a dosage liability threshold below the level of heterozygosity. In summary, we delineated a novel genetic condition caused by cohesin release factor deficiency, nominated *WAPL* as a driver gene within a genomic disorder region, and further illuminated dosage sensitivity of human cohesin.

## Introduction

The three-dimensional (3D) folding of human chromosomes is essential for proper regulation of gene expression throughout development, for example by bringing together promoters and tissue-specific distal enhancers (Ball et al., 2014; Hafner and Boettiger, 2023; Liu et al., 2023). Among the proteins that shape the genome in 3D are cohesins, which are critical for forming topologically-associating domains (TADs) (Rowley and Corces, 2018), CTCF-anchored chromatin loops (Sanborn et al., 2015), and other structures (Cuadrado et al., 2019; Yan et al., 2013). Mutations in cohesin genes are the cause of ‘cohesinopathies,’ a family of Mendelian conditions that typically present as syndromic neurodevelopmental disorders (NDDs). Most notable among these is Cornelia de Lange syndrome (CdLS; OMIM PS122470), a severe NDD and malformation syndrome caused by heterozygous or hemizygous nullimorphic, hypomorphic, or dominant negative variants in genes encoding the core cohesin ring, cohesin-associated proteins, the cohesion loader MAU2, or – most commonly – the cohesin loading and extrusion factor NIPBL. These variants are presumed to cause disease via alterations to genome organization, transcription, and other downstream effects (Garcia et al., 2021; Liu et al., 2009; Sakata et al., 2025; Weiss et al., 2021).

Cohesin binds chromatin with the aid of a loading complex (NIPBL and MAU2), then extrudes loops of DNA to form 3D genome structures (Ciosk et al., 2000; Davidson et al., 2019; Wendt et al., 2008) (Fig. 1a). Cohesin loading and loop extrusion are countered by cohesin removal/resetting that occurs by way of a release complex (WAPL bound to PDS5A or PDS5B) (Chan et al., 2012; Gandhi et al., 2006; Huis in ’t Veld et al., 2014; Kueng et al., 2006; Silva et al., 2020; Tedeschi et al., 2013; Zhang et al., 2021). These opposing forces render genome organization a dynamic and cyclical process (Gabriele et al., 2022; Mach et al., 2022).

**Figure 1.**
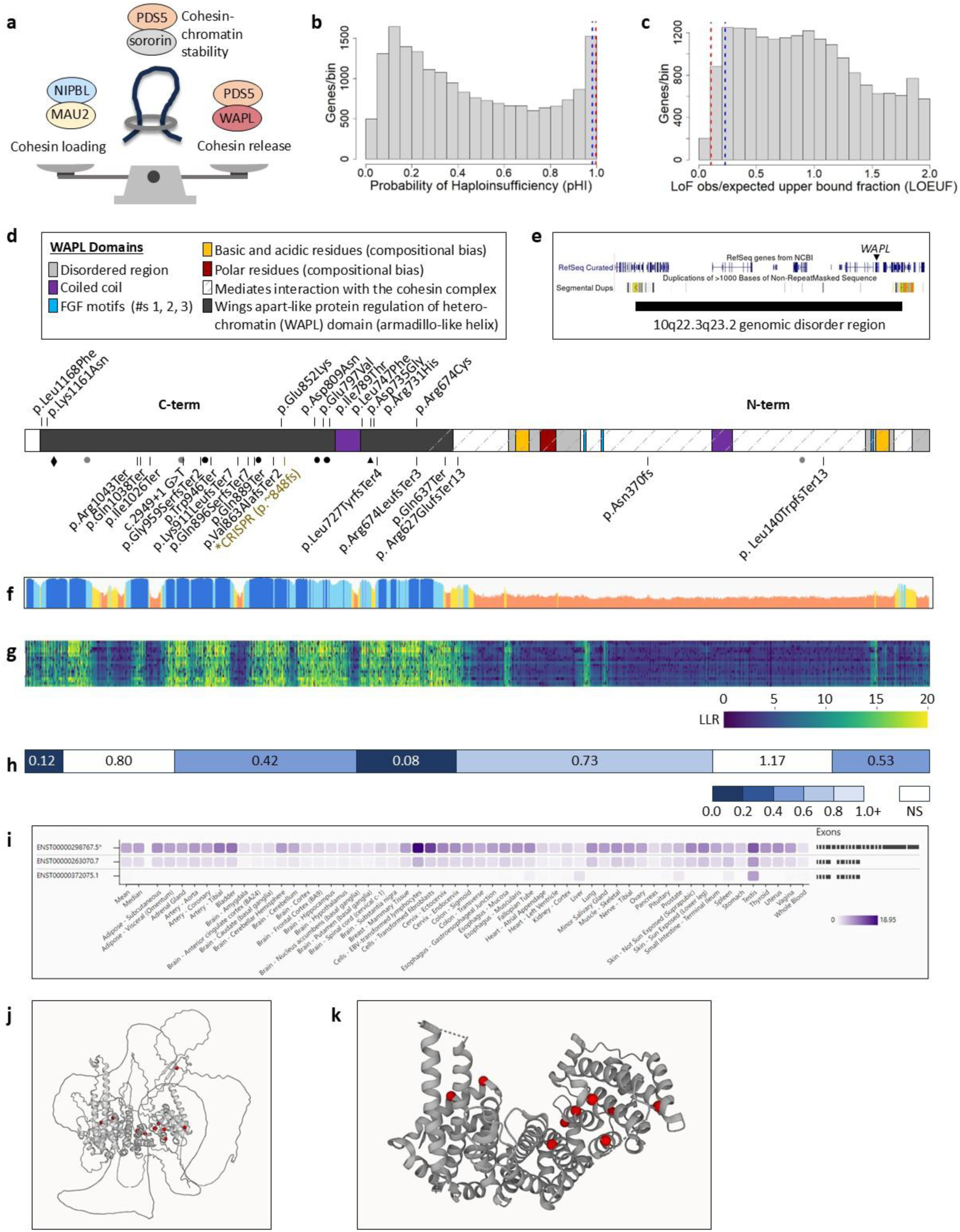
Disease-causing variants in *WAPL*, a cohesin release factor, were predicted then discovered. **a.** Simplified depiction of cohesin balance. Cohesin-mediated genome organization is initiated by loading of the cohesin ring (dark grey) by NIPBL and MAU2 onto chromatin (dark blue), followed by loop extrusion. WAPL resets this process by removing cohesin from chromatin. PDS5 facilitates both cohesin release by WAPL and cohesin stability by sororin, among other functions (Zhang et al., 2021). **b-c.** Probability of haploinsufficiency (pHaplo, b) of 18,641 autosomal genes, via (Collins et al., 2022), and loss of function observed/expected upper bound fraction (LOEUF, c) of 18,567 genes, via gnomAD (Karczewski et al., 2020). Red lines, average of known autosomal dominant CdLS genes *NIPBL*, *RAD21*, and *SMC3*. Blue lines, average of cohesin release factor genes *WAPL*, *PDS5A*, and *PDS5B*. See also Table S1. **d.** 27 *WAPL* variants identified in subjects including predicted damaging missense (ms, top) and loss-of-function (pLoF, bottom), plotted in protein space in genomic orientation. Domains of the canonical protein (1190 amino acids) via UniProt (https://www.uniprot.org) and Ensembl (https://ensembl.org). gnomAD pLoF variants passing quality filters are denoted by black (frameshift, stop gain) and grey (splice) circles. The diamond denotes the point after which escape from nonsense-mediated decay would be predicted (codon 1151). The triangle is the start position for minor 3′ isoforms in (h). The cut site for CRISPR/Cas9 experiments is in gold text. The p.Val863AlafsTer2 variant was seen in two individuals. Missense and pLoF variants both cluster in the C-terminal half of WAPL (p=0.0020 and p=0.0024, respectively by two-tailed binomial test). **e.** Location of *WAPL* within the 7.8 Mb 10q22.3q23.2 reciprocal genomic disorder region (black bar; via ClinGen Pathogenic CNV track of the UCSC Genome Browser (https://genome.ucsc.edu/)). **f.** AlphaFold per-residue structural confidence (pLDDT, or predicted local distance difference test), via (Kwon et al., 2024). Y-axis range is 0-100, with scores <50 (orange) being a reasonably strong predictor of disorder (https://alphafold.ebi.ac.uk/). **g.** Deep protein language model-derived predicted pathogenicity of every possible missense change in *WAPL*, via (Brandes et al., 2023). Y-axis, amino acid. X-axis, residue. Higher LLR (log-likelihood ratio) indicates more likely deleteriousness **h.** Regional missense constraint in amino acid space, via gnomAD v2.1.1 (https://gnomad.broadinstitute.org/). Scores indicate the fraction of expected amino acid variation in a population cohort. **i.** *WAPL* transcript expression by tissue, via gnomAD. C-terminal isoforms exist but are more lowly expressed. **j-k.** 3D structures of WAPL via AlphaFold (j) (https://www.alphafold.ebi.ac.uk/entry/Q7Z5K2) that demonstrates low-confidence structure for its N-terminal half, and via crystallography of its C-terminal half (k) (Ouyang et al., 2013) (pdb ID 4k6j, amino acids 631-1190). Case missense variants (red) show possible clustering. Plots created via (https://g2p.broadinstitute.org; (Kwon et al., 2024)).

Data from a very large-scale dosage sensitivity prediction analysis of rare copy number variants (rCNVs) across human phenotypes in almost 1M individuals (Collins et al., 2022) demonstrate that autosomal cohesinopathy genes, including the cohesin loader *NIPBL*, have both high predicted haploinsufficiency (pHaplo) scores and high triplosensitivity (pTriplo) scores, suggesting that these loci are bidirectionally dosage sensitive (Table S1). This is corroborated by the existence of known human phenotypes resulting from both heterozygous copy loss of *NIPBL* in CdLS and duplication of this gene in the 5p13 duplication syndrome (OMIM #613174) (Yan et al., 2009). These data, and the known dynamism between TAD formation and resetting, suggests that a disruption of cohesin balance in either direction may be pathological. Thus, it is surprising that neither WAPL nor its binding partners PDS5A and PDS5B – the three cohesin release factors – have been systematically assessed as potential disease genes (Yuan et al., 2019).

We hypothesized that cohesin release factor genes may also be disease genes, would herald a novel class of cohesinopathies, and that their clinical and molecular features could provide insights extending to CdLS and related syndromes. Furthermore, we hypothesized a potential intersection between cohesinopathies and genomic disorders (GDs) – genetic conditions caused by large, recurrent deletions and duplications of genomic intervals mediated by non-allelic homologous recombination between segmental duplications (Carvalho et al., 2010). Specifically, *WAPL* resides in the 10q22.3q23.2 GD region, which when deleted (OMIM **#**612242) or duplicated is associated with risk of NDDs, but for which no driver gene(s) has been confirmed. In the present work, we leveraged patient-level deep phenotyping, large-cohort disease association approaches, mouse models, and – to recapitulate cohesin release factor deficiency in a species-, zygosity-, and NDD-relevant system – CRISPR genome engineering in human iPSCs and neural cell lines. Our work nominates *WAPL* as a novel cohesinopathy gene and a driver of 10q22.3q23.2 GD and describes the phenotypic consequences and transcriptomic disturbances resulting from haploinsufficiency of this cohesin release factor.

## Results

### A comprehensive case series of patients harboring mutations in cohesin release factor genes

To explore the extent to which *de novo* and ultra-rare inherited variants in three core cohesin release factor genes cause human phenotypes, we identified, recruited, and phenotyped individuals with predicted LoF or damaging missense variants in these genes. We evaluated children and adults with heterozygous and predicted damaging variants in *WAPL* (n = 27), *PDS5A* (n = 8), and *PDS5B* (n = 8) (Fig. 1d; Tables 1, S4). Where parental information was available and inheritance could be determined, we discovered that the majority of *WAPL* variants (19 of 22), the majority of *PDS5A* variants (3 of 4), and all *PDS5B* variants (4 of 4), arose *de novo*. Inherited variants included a *WAPL* stop gain variant (WAPL case 9) from an asymptomatic mother, a *WAPL* frameshift variant (WAPL case 13) from a father with a history of learning difficulties, a *WAPL* missense variant (WAPL case 26) from a mother with a history of speech delay, a PDS5A missense variant (PDS5A case 6) from a mother with an unknown phenotype, and a *PDS5B* stop gain variant (PDS5B case 8) from an apparently healthy mother. No individuals were found with biallelic predicted damaging variants in any of these genes.

Clinical assessment of these individuals provided evidence of a *WAPL*-related set of phenotypes. Of 25 subjects with *WAPL* variants for whom detailed information was available, mild-moderate developmental delay/intellectual disability is present in most (n = 21) (Tables 1, S4; Fig. 2a). Note that Table 1, Table S4, Fig. 2, and Fig. S12 have had case-level data including age, sex, phenotype, and photos removed, per medRxiv guidelines. Many subjects have craniofacial dysmorphism (n = 15); although some facial features were shared by multiple individuals (e.g. retrognathia, high columella), there is not a typical set of features across all individuals. Many subjects also have behavioral features (n = 11), organ malformation (e.g., clubfoot (n = 4), cardiac (n = 7)), and variable neurological features (n = 14; including 4 with hypotonia, 2 with small head circumference).

**Figure 2.**
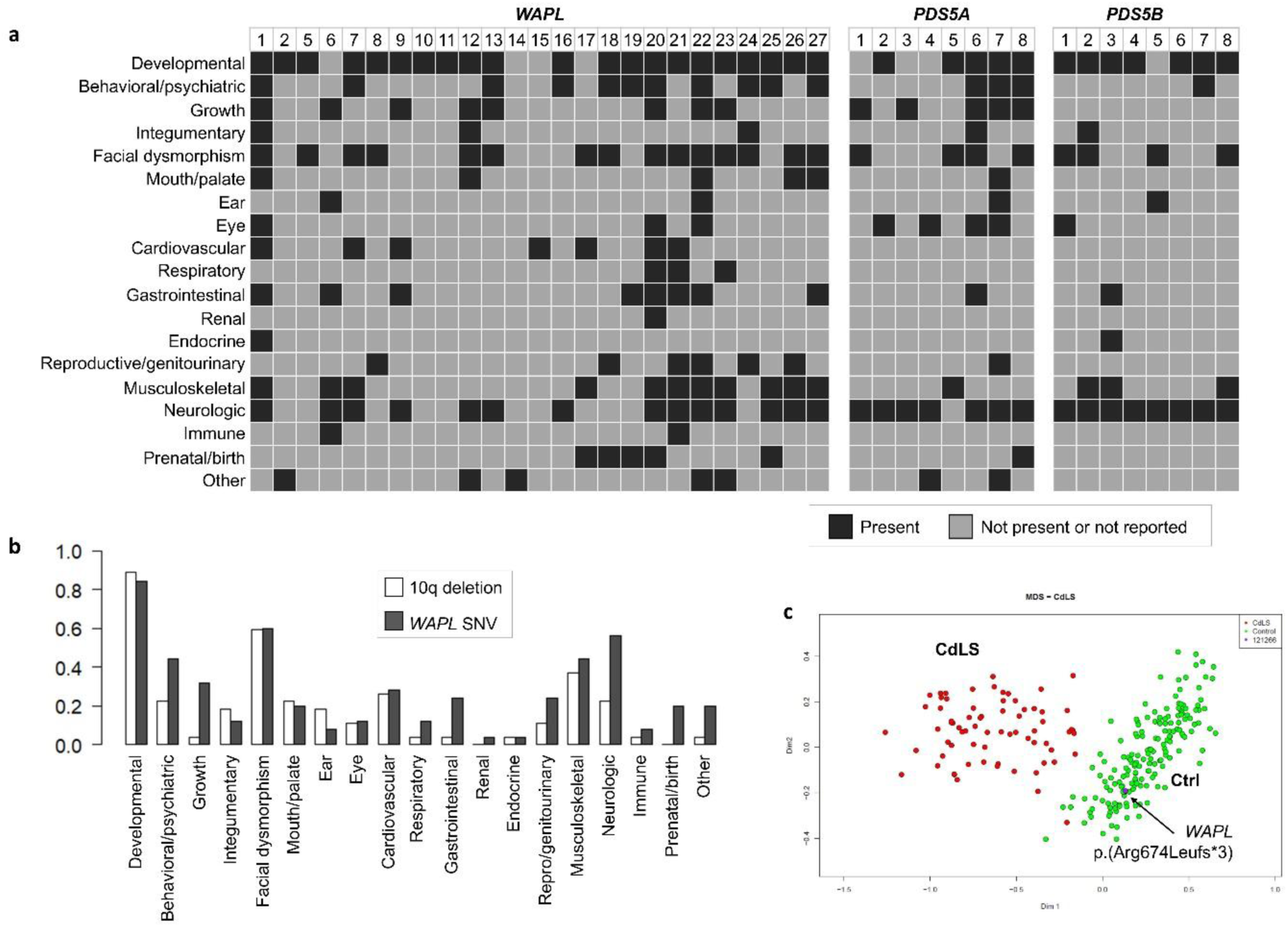
Cohesin release factor variants are associated with neurodevelopmental and other features. **a.** Phenotypes (rows) in subjects (columns) with heterozygous predicted damaging variation in *WAPL*, *PDS5A*, or *PDS5B*. Most individuals with *WAPL* SNVs have mild-moderate developmental problems, and facial dysmorphism and neurological issues are each present in about half of subjects. Many individuals have behavioral challenges, growth delays, and musculoskeletal defects (e.g., clubfoot). WAPL subjects 3-4 were removed from this figure for lack of phenotype detail. *PDS5A* and *PDS5B* cases are enriched for developmental and variable neurological issues, but they do not coalesce into defined syndromes. Phenotypes are described in Tables 1 and S4. **b.** A comprehensive literature review identified 27 individuals with recurrent 10q dels. Sibs were each retained rather than collapsed to one per family. The similar frequency of some features between 10q del patients and *WAPL* point mutation patients (e.g. developmental, dysmorphism, musculoskeletal) suggests that *WAPL* may be a driver of these phenotypes. **c.** Methylation signature of a pLoF *WAPL* variant p.(Arg674LeufsTer3) compared to known CdLS genes. Subjects with *WAPL* SNVs do not have a DNA methylation pattern that clusters with the overall methylation signature of CdLS.

**Table 1.**
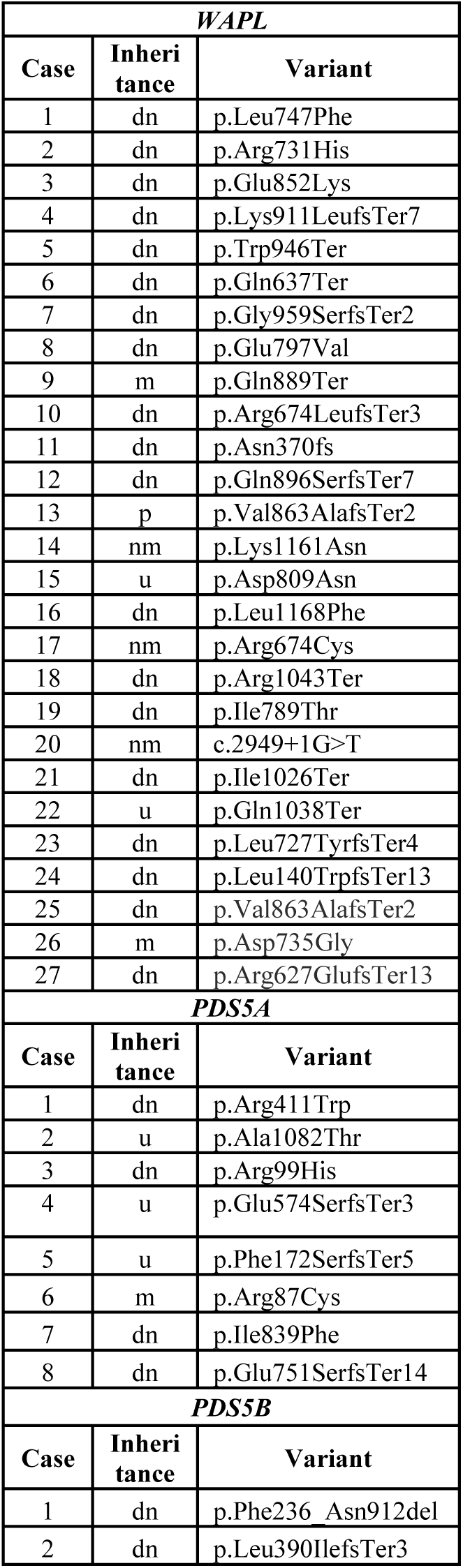
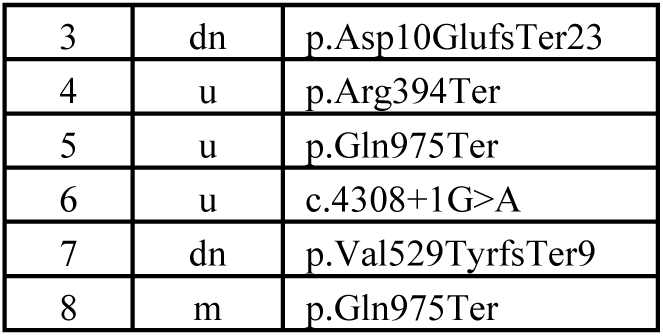
*WAPL*, *PDS5A*, *PDS5B* variants and associated phenotypes. Case-level phenotypes have been removed, per medRxiv lines. See Table S4 for additional information. dn, de novo. u, unknown. m, maternal. p, paternal. nm, non-maternal.

Subjects with variants in *PDS5A* or *PDS5B* presented with considerable morbidity, and every subject had neurodevelopmental and/or neurological features (Tables 1, S4; Fig. 2a; Fig. S12). Yet, these features were variable and less convincingly coalesced into unified syndromic presentations like those observed for *WAPL*. Thus, our subsequent analyses focused on the clinical, functional, and molecular signatures associated with *WAPL*-associated mutations.

Within *WAPL*, missense and pLoF variants cluster away from the N-terminal region and into subregions of the more deeply conserved and more structured C-terminal region (Fig. 1d, f-h, j-k) (Brandes et al., 2023). This may suggest a relative importance of C-terminal *WAPL* isoforms (Fig. 1i) and the C-terminal WAPL domain. Of note, owing to the lower conservation of the majority of the N-terminus, fewer possible N-terminal missense variants would be judged as pathogenic via conservation- and structure-based *in silico* deleteriousness prediction tools, which were prerequisites for inclusion in our study (Methods).

### WAPL as a driver of phenotypes in the 10q22.3q23.2 genomic disorder

*WAPL* is contained within chromosome 10q22.3q23.2, which is subject to recurrent deletions and duplications of ∼7.8 Mb between flanking low-copy repeats termed LCR3 and LCR4 (Balciuniene et al., 2007) (Fig. 1e). Case reports have associated these deletions with NDD, craniofacial dysmorphism, and other features including congenital heart disease and limb abnormalities, while duplications present with milder developmental effects. However, a consensus phenotype has not been delineated, and driver genes have been proposed but not confirmed (Azidane et al., 2024; Breckpot et al., 2012; Coelho Molck et al., 2017; Saito et al., 2012) (Table S5).

To inform on *WAPL* dosage sensitivity and to make phenotypic comparisons between 10q dels and *WAPL* single nucleotide variants (SNVs), we sought to define the 10q22.3q23.2 deletion phenotype. We screened our cross-disorder rare copy number variant (rCNV) microarray dataset composed of 458,326 cases (i.e., samples with at least one diagnosed condition) and 491,952 control individuals (Collins et al., 2022). Our analysis identified 41 carriers of the canonical 10q22.3q23.2 deletion, which were strongly associated with developmental phenotypes (OR = 20.9; two-sided Fisher test of proportions p = 1.8e-10). The most common HPO terms were abnormality of the nervous system (n = 24; 59%), neurodevelopmental abnormality (n = 21; 51%), abnormality of head or neck (n = 9; 22%), abnormality of the face (n = 8; 20%), abnormality of the cardiovascular system (n = 5; 12%), growth abnormality (n = 5), and atypical behavior (n = 5) (Fig. S17). We also identified 24 carriers of the canonical duplication. In line with the previously purported milder effect of the duplication, the enrichment of duplication carriers among cases was more modest (OR = 3.2; p-value = 0.013), although a similar pattern of affected organ systems with the deletion carriers was noted (Fig. S17). This suggests that the dosage of the region is under tight control. Next, we performed a literature review and identified 27 published cases of the deletion. The most common phenotypic features among these 27 cases are NDDs, facial dysmorphisms, musculoskeletal abnormalities (including clubfoot in 6 individuals), and congenital heart defects (Fig. 2b, Table S5).

Together, this comprehensive phenotypic analysis of 68 individuals carrying the recurrent 10q22.3q23.2 deletion points to a phenotype that is similar to that of patients with *WAPL* point mutations. On a more granular level, the involvement of several organ systems is of equivalent prevalence between individuals with *WAPL* SNVs and those with 10q dels (Fig. 2b); these include NDD, facial dysmorphisms, cardiovascular defects, and musculoskeletal anomalies, suggesting *WAPL* may be a driver of 10q GD features.

### Episignature of cohesin release factors

Global DNA methylation aberrancies have been found in peripheral blood of patients with CdLS (Aref-Eshghi et al., 2020), which are both biologically informative (i.e., evidence of downstream effects of cohesin loss on the epigenome) and clinically useful (i.e., as a diagnostic aid in cases of cohesin gene variants of uncertain significance). We applied EpiSign^TM^ analysis to subjects’ peripheral blood DNA (Table S6) to establish a genome-wide DNA methylation episignature. Three distinct analyses were conducted. Analysis 1 combined truncating variants affecting *WAPL* (including 10q dels), *PDS5A*, and *PDS5B*. Analysis 2 combined *WAPL* missense SNV and 10q del cases. Analysis 3 assessed only *WAPL* SNV (protein-truncating and missense) cases. Informative and variably methylated sites were selected, and their ability to distinguish case samples from controls was confirmed using hierarchical clustering and multidimensional scaling (MDS) (Fig. S1). The reproducibility of the identified profiles was assessed using leave-one-out cross-validation (LOOCV) (Fig. S2). The left-out sample clustered with the other cases in 11 of 14 LOOCV iterations of Analysis 1, in 8 of 10 iterations of Analysis 2, and 0 of 8 iterations of Analysis 3 (i.e., no methylation signature for *WAPL* SNVs alone was found). A support vector machine (SVM) classifier was developed by training case samples against their matched controls, along with 75% of control samples from the EpiSign Knowledge Database (EKD) and thousands of samples from individuals with other rare genetic disorders included in the EpiSign v5 clinical classifier. This was done for Analyses 1 (Fig. S3a) and 2 (Fig. S3b). The remaining 25% of control and other episignature disorder samples were used to test the model. Both models demonstrated high specificity, as the majority of testing samples from other episignature disorders (including CdLS) received very low methylation variant pathogenicity (MVP) scores.

In summary, we identified an overall cohesin release factor deficiency-associated DNA methylation signature and a DNA methylation signature combining *WAPL* SNV and 10q del samples. We did not identify a DNA methylation signature for *WAPL* SNVs alone. These DNA methylation patterns do not mimic the overall CdLS methylation pattern (Fig. 2c).

### In vitro human stem cell and neuronal disease modeling of 10q GD and postulated driver WAPL

We next sought to explore the functional genomic consequences of *WAPL* haploinsufficiency. We applied CRISPR engineering to isogenic human induced pluripotent stem cells (iPSCs) to generate biological replicate lines housing heterozygous (het) *WAPL* frameshift indel (fs) variants (7 het fs lines and 5 “paired” unedited wt lines; details in Methods) or the 10q recurrent genomic deletion (del) or duplication (dup) that contains *WAPL* (6 het del, 6 het dup, 6 paired wt) (Figs. 3a, S5; Table S7). Despite high efficiency of editing (see Methods), no *WAPL* biallelic fs clones were recovered, suggesting that complete loss of this gene is lethal in iPSCs. Of note, all replicates represent independently edited iPSCs with unedited lines exposed to the same CRISPR guides and sorted to single cells in the same experiment, with the exception of 10q dups, which were replicates of one edited line (Methods).

**Figure 3.**
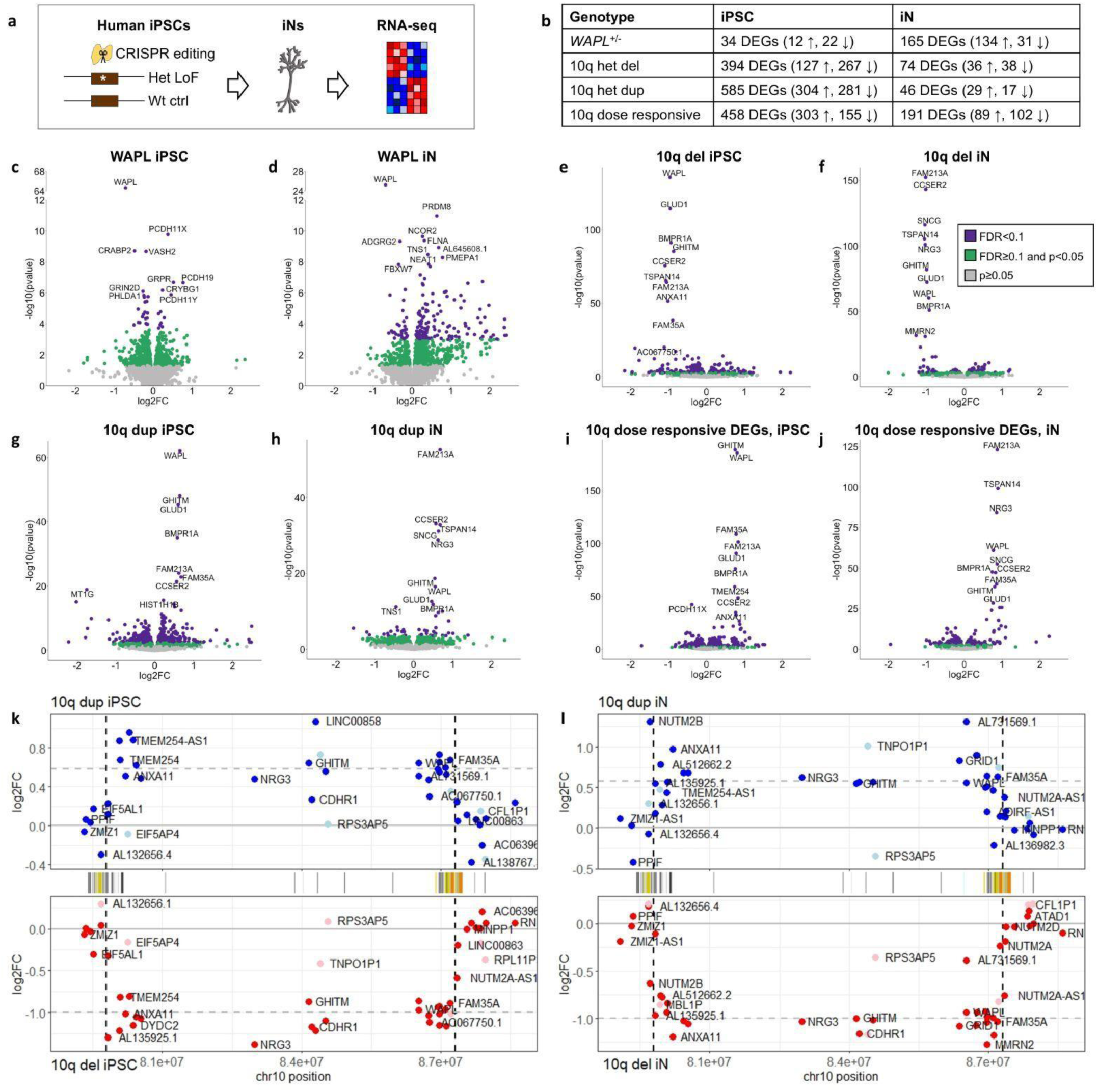
CRISPR-engineered human iPSC and iN models of *WAPL*^+/-^ and heterozygous 10q del or dup reveal disease-relevant transcriptional disturbance. **a.** The CRISPR-Cas9 system was employed to introduce heterozygous fs indels in *WAPL* or to generate the 7.8 Mb 10q del or dup. Edited iPSCs were differentiated into iNs. iPSCs and iNs were RNA-sequenced. **b.** Genotypes and number of differentially expressed genes (DEGs) at FDR <0.1. ↑ upregulated. ↓ downregulated. See panels (i-j) for description of “dose responsive.” **c-h.** Volcano plots showing DEGs. Purple, FDR<0.1. Green, FDR≥0.1 and p<0.05. Gray, p≥0.05. The x-axis range is limited to log_2_ of - 2 to 2 for clarity. **i-j.** Volcano plots of genes, genome-wide, that are reciprocally dosage sensitive in 10q iPSCs (i) and iNs (j). Data are derived from an analysis in which del, dup, and wt samples were assigned to a (−1,1,0) vector. With this definition, 10q region genes correlate positively with 10q copy number and have positive fold change (FC). **k-l.** Expression changes, plotted as log2 FC of genes within and flanking the 10q del and dup in iPSCs (k) and iNs (l). Deleted (bottom panels, red) or duplicated (top panels, blue) genes are expectedly up or down-regulated in both direction and magnitude, and flanking genes are generally unaltered in their expression (Table S10). Specifically, average fold changes for protein-coding genes, after removal of genes with paralogs (e.g. *NUTM2A*/*B*/*D*) and genes with low expression (baseMean <25), were as follows: del iPSC decrease by 50% of wt, dup iPSC increase by 53%, del iN decrease by 51%, and dup iN increase by 57%. All of these genes individually also have significantly altered expression at a significance threshold of p<0.05. Horizontal dashed lines show the predicted log_2_ FC for loss or gain of one copy. Pseudogenes are in light colors. Genes are plotted by their end coordinates. UCSC segmental dups track is shown in between del and dup plots.

Given that iPSCs are an undifferentiated, early, dividing cell type in which cohesin complex function is largely unknown, we also sought to characterize the relative roles of *WAPL* and 10q dels/dups in transcriptional regulation in the brain. Thus, all 30 lines were differentiated into induced glutamatergic neurons (iNs) (Fig. 3a). Total RNA-seq confirmed a decrease in expression of *WAPL* in het fs (by 39% compared to wt in iPSCs and by 38% in iNs) and 10q del (by 49% in iPSCs and by 48% iNs) lines, and an increase in 10q dup (by 56% in iPSCs and by 47% in iNs) lines (Fig. S6; statistics in figure legend). These levels of expression were corroborated by qPCR of mRNA. Allelic expression analysis confirmed that the fs indel alleles of all *WAPL*^+/-^ lines were diminished at the RNA level (by 92.6% of wt allele in *WAPL* iPSC and by 94.0% in *WAPL* iN), indicating effective elimination via nonsense-mediated decay (Fig. S6; statistics in figure legend).

### Transcriptional impact in WAPL and 10q cell models implicates developmental processes

We performed transcriptome analyses in our iPSC and iN models using total RNA sequencing (Figs. 3, S7-S10). Differentially expressed genes (DEGs) are summarized in Tables S8-S9 and Fig. 3b. In addition to DEGs attributable to each individual genotype (Fig. 3c-h), we combined 10q del and dup data to call 458 and 191 dose-responsive DEGs (e.g., those with opposing fold changes in del vs dup) in iPSCs and iNs, respectively (Fig. 3i-j). There was no evidence of unexpected expression effects (i.e., dosage compensation or position effect) of genes within or flanking the 10q region (Fig. 3k-l; Table S10).

We compared DEGs (p < 0.05) between cell types (iPSCs vs iNs) for each genotype (*WAPL*^+/-^, 10q del, 10q dup) (Figs. 4, S11). *WAPL* iPSC and iN DEGs overlapped (p = 4.46e-8), but the fold change (FC) of overlapping genes did not correlate (Spearman = -0.10, p = 3.34e-1) (Fig. 4a). 10q del iPSC and iN DEGs, excluding genes within the deleted segment, overlapped (p = 6.87e-5) and the FC of overlapping genes was correlated (Spearman = 0.60, p < 1e-100) (Figs. 4b, S11a). 10q dup iPSC and iN DEGs, excluding genes within the deleted segment, overlapped (p = 3.43e-3), but the FC of overlapping genes did not correlate (Spearman = 0.09, p = 2.85e-1) (Figs. 4c, S11b). 10q dose responsive DEGs, excluding genes within the deleted segment, overlapped (p = 8.06e-12) and the FC of overlapping genes was correlated (Spearman = 0.31, p = 5.88e-6). To summarize, *WAPL***^+/-^**, 10q del, 10q dup, and 10q dose responsive DEGs each exhibit partial overlap between iPSCs and iNs, suggesting shared but also cell type-specific transcriptomic effects.

**Figure 4.**
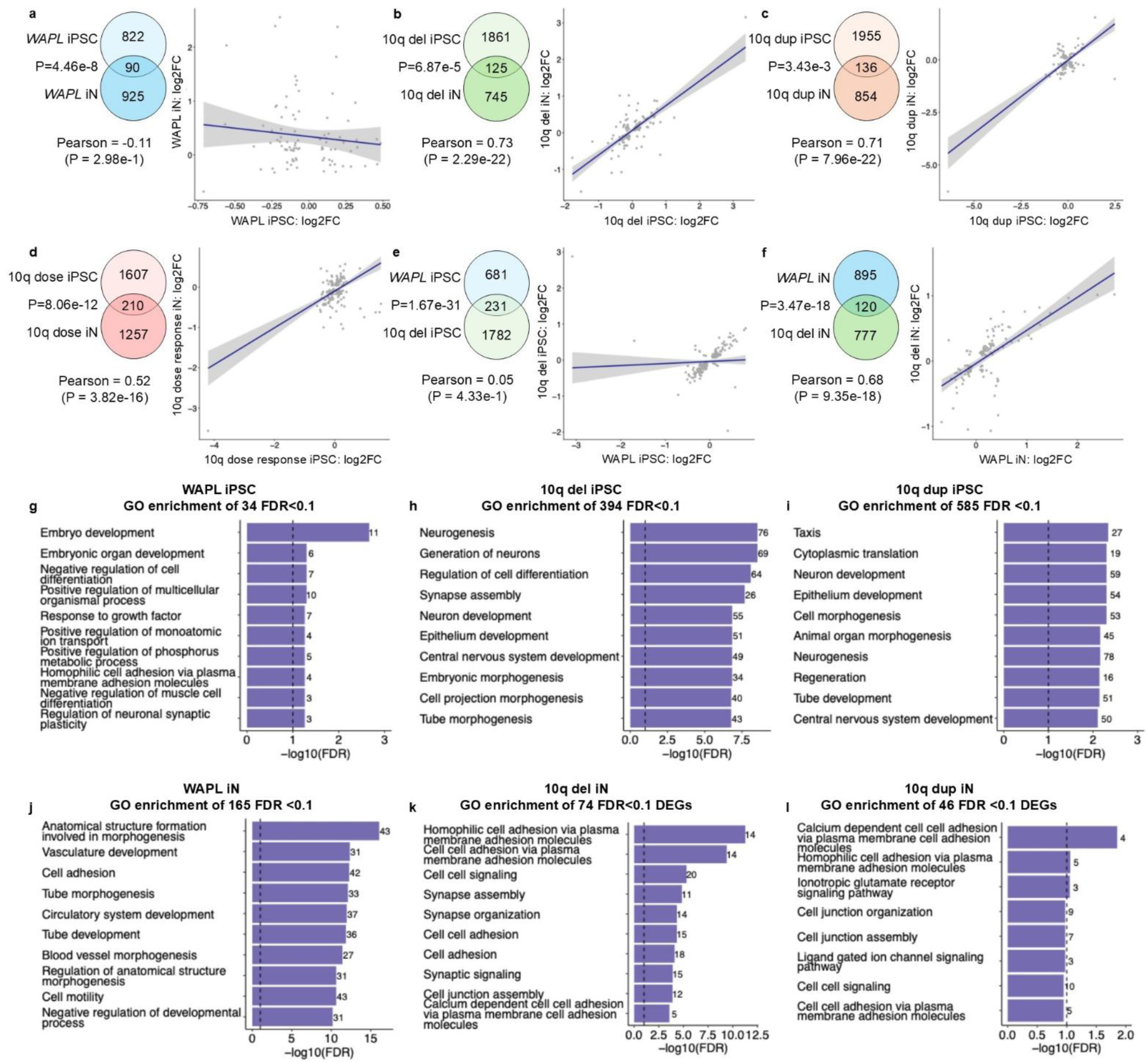
Signatures of altered gene expression in models of *WAPL*^+/-^ and heterozygous 10q del or dup. a-d. DEG comparisons between cell types within a given genotype. *WAPL***^+/-^** DEGs (a) significantly overlap between cell types (iPSCs and iNs); however, the fold change (FC) of the intersection of these genes is not correlated. 10q del (b), 10q dup (c), and 10q dose responsive (d) DEGs, with genes within the 10q GD region removed, each significantly overlap between iPSCs and iNs; the FC of the intersection of these genes is correlated for 10q del and 10q dose responsive DEGs. Overlap of DEGs at the p<0.05 threshold are shown as Venn diagrams, and the correlation of the FC of the intersection of these genes as dot plots. P values of gene overlap are from Fisher’s exact test. Linear regression lines are shown, and correlation statistics are listed below each dot plot. The axes are limited to -2 to 2 for clarity (see Table S9 for complete DEG list). **e-f.** DEG comparisons between genotypes within a given cell type. *WAPL*^+/-^ and 10q del DEGs significantly overlap and the FCs of the intersection of these genes are correlated in iPSCs (d) and iNs (e). **g-l.** The most significant biological processes enriched among DEGs (at a threshold of FDR<0.1) from each model, via gene ontology (GO) analysis. Dotted lines are FDR <0.1

To infer the disrupted biological processes in our cell models, we performed functional enrichment analyses of DEGs at FDR<0.1 against gene ontologies (GO; Fig. 4g-l). For example, in *WAPL*^+/-^ iPSCs, embryo development was the most enriched process; this may indicate that the developmental defects in the *WAPL*-related disorders have, at least partially, a very early origin. As another example, all iN models feature terms related to cell-cell interactions, suggesting potential relevance of this process to brain pathobiology upon *WAPL* or 10q mutation. The distinct terms enriched in iPSCs vs iNs may further suggest cell-type specific effects.

### WAPL as a driver of 10q molecular pathogenesis

To find evidence for or against *WAPL* being a driver gene of transcriptional disturbances in 10q del GD, we first compared DEGs at p < 0.05 between *WAPL*^+/-^ and 10q del. *WAPL*^+/-^ and 10q del DEGs overlapped significantly in iPSCs (p = 1.67e-31) and the FC of overlapping genes correlated strongly (Spearman = 0.75, p < 1e-100) (Fig. 4d). This was also true in iNs (overlap p = 3.47e-18; overlapping genes’ FC Spearman 0.67, p < 1e-100) (Fig. 4e). These results indicate similar transcriptional effects between *WAPL*^+/-^ and 10q del, providing evidence of *WAPL* as a strong driver of 10q molecular pathogenesis.

### Wapl dosage and early mouse development

We next sought evidence for an *in vivo* impact of *Wapl* haploinsufficiency, in mice. Previous mouse models indicate that complete loss of Wapl (*Wapl*^-/-^) is embryonic lethal (Kean et al., 2022; Oikawa et al., 2004; Tedeschi et al., 2013), consistent with the absence of homozygous *WAPL* LoF mutations in our case series and cell lines. *Wapl*^+/-^ mice are viable, fertile, and weaned at expected Mendelian frequencies, however embryos show mildly reduced body weight (Kean et al., 2022). It is of interest to determine whether these animals may serve as a model of human WAPL deficiency syndrome and/or of cohesin release deficiency more broadly, and to determine the tolerated window of cohesin balance in mice.

We first utilized *Wapl*^+/-^ mice exhibiting a 50% reduction in *Wapl* mRNA and Wapl protein compared with wild type (wt) (Kean et al., 2022). To determine the effects of reduced *Wapl* on neuromuscular development in neonates, we assessed p2 to p20 animals daily. *Wapl*^+/-^ mice outperformed wt littermates in regard to righting reflex (Fig. 5a; p_genotype_ = 0.03 by 3-way ANOVA test over p2-p4) and negative geotaxis (Fig. 5b; p_genotype_ = 0.0006 by 3-way ANOVA over p2-p6). Forepaw grasping reflex is acquired at similar ages in wt and mutant mice: 3.1±0.4 days and 3.0±0.3 days in wt and het females (p = 0.79, t-test) and 2.7±0.3 days and 3.3±0.4 days in wt and het males (p = 0.27) (values are ±SEM). We assayed muscle strength using the Four Limb Hang assay (Fig. 5c). Younger *Wapl^+/-^* neonates (days 5-7) outperform wt neonates (p_genotype_ = 0.02 by 3-way ANOVA over p5-p8) but behaviors of the two groups then become indistinguishable. Each day we assigned pups a gait score based on criteria described in Methods. *Wapl^+/-^* pups started walking and crawling as quickly as *Wapl^+/+^*pups (Fig. 5d; p_genotype_ = 0.49 by 3-way ANOVA). *Wapl^+/-^*pups are smaller than wt cohorts (Fig. 5e; p_genotype_ = 0.0001 by 3-way ANOVA). In summary, newborn heterozygotes are mildly smaller than wt but do not show early motor disadvantages.

**Figure 5.**
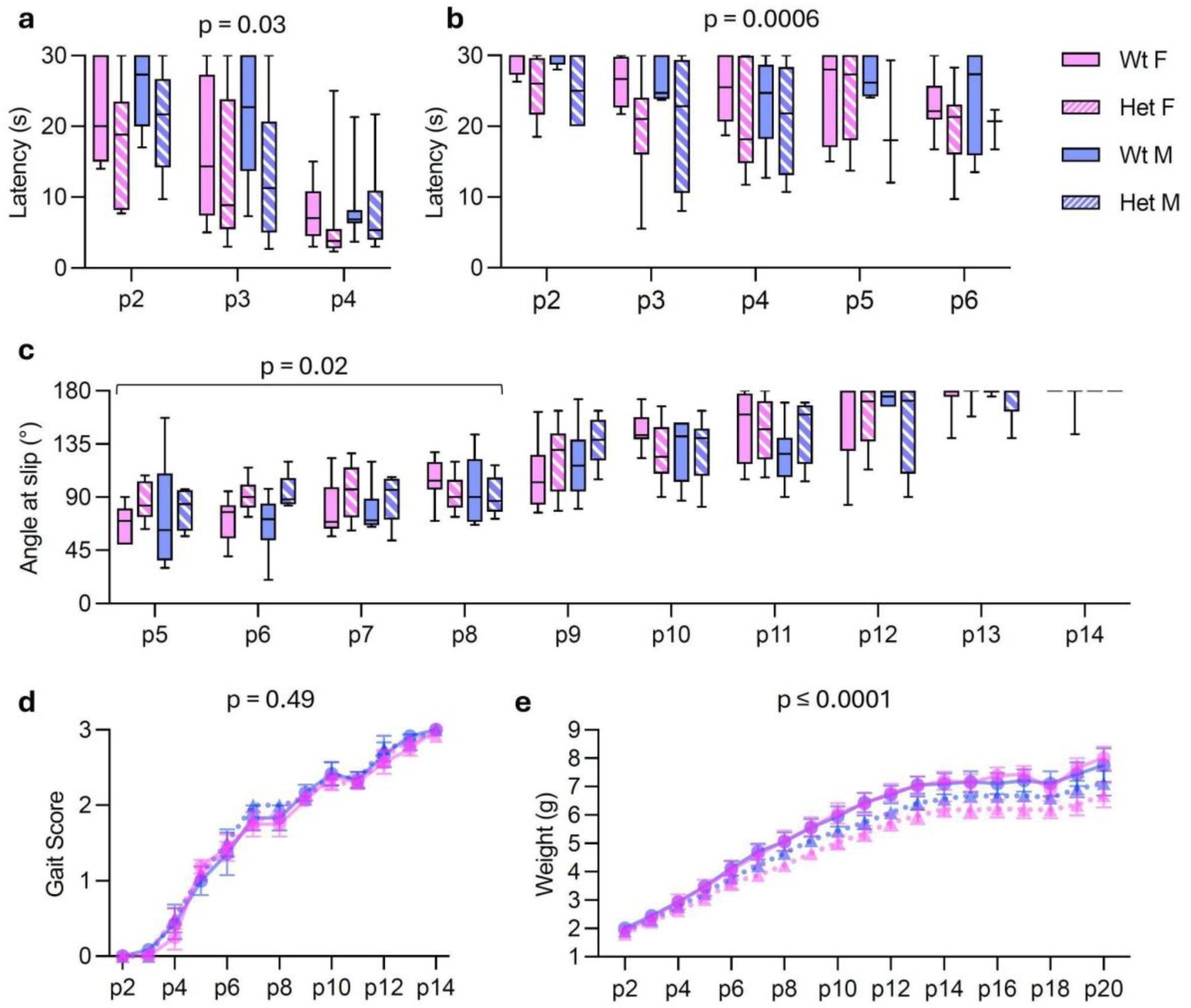
Analyses of neonatal behavior in *Wapl*^+/+^ and *Wapl*^+/-^ pups. **a.** Righting Reflex. *Wapl*^+/-^ pups performed better than their wt cohorts (p_genotype_ = 0.03). **b.** Geotaxis. *Wapl*^+/-^ pups outperformed their wt cohorts (p_genotype_ = 0.0006). **c.** Four Limb Hang. Younger *Wapl*^+/-^ pups (≤8 days) performed better than their wt cohorts (p_genotype_ = 0.02) but the effect of genotype diminishes with age. **d.** Gait. Genotype has no discernible effect. **e.** Growth curves. *Wapl*^+/-^pups are significantly lighter than wt (p_genotype_ = 0.0001). Data in (a-c) are presented as median, the 25-75% quartiles, and max/min values. Data in (d-e) are presented as mean ± SEM. *Wapl*^+/+^ female = solid magenta; *Wapl*^+/-^ female = hatched magenta; *Wapl*^+/+^ male = solid blue; *Wapl*^+/-^ male = hatched blue. For (d) and (e), circles/solid lines = wt; triangles/dashed lines = het. N = 32: 8 *Wapl*^+/+^ females; 12 *Wapl*^+/-^ females; 6 *Wapl*^+/+^ males; 6 *Wapl*^+/-^ males. Statistical significance was evaluated by 3-way ANOVA with genotype, sex, and age as independent variables.

### Wapl dosage and adult behaviors

In the Open Field Test, adult *Wapl*^+/-^ and wt animals did not travel different distances or speeds or spend different fractions of time moving (Fig. 6a-i, ii, iii). Also, *Wapl*^+/-^ and wt mice did not display different behaviors within the center region (Fig. 6a-iv, v, vi), indicating that neither exploratory behavior nor anxiety are genotype dependent. In the Y-Maze, spontaneous alternation is assessed as a measure of spatial, short-term memory. Mice prefer novel experiences to familiar ones; thus, higher alternation among the arms of the maze is associated with better short-term memory. Het females and males alternated 11% and 10% more than their wt counterparts (Fig. 6b; p_genotype_ = 0.002 by 2-way ANOVA). Consistent with results from the Open Field Test, we did not observe differences in the distances traveled, speeds, or mobility patterns of *Wapl*^+/-^ and wt mice during the Y-Maze (Fig. S14a). In the Morris Water Maze (MWM), a measure of learning and long-term memory, *Wapl^+/-^* mice, especially *Wapl^+/-^* females, show poorer performances relative to wt cohorts (p_genotype_ = 0.0002 by 3-way ANOVA) (Fig. 6c, Fig. S14b-d, g). The deficiency cannot be explained by impaired swimming ability or by impaired visual acuity (Fig. S14e-f). Thus, differences in latency are likely attributable to differences in learning. Looking at results for individual mice (Fig. S14g), a major difference is that *Wapl^+/-^* mice, but especially *Wapl^+/-^* female mice, perform inconsistently from day to day. Contextual and cued fear conditioning tests assess associative learning and are measures of hippocampus and amygdala function. Wt and het mice performed equally well in these tests (Fig. 6d). Lastly, we assayed motor coordination via the rotarod test. *Wapl*^+/-^ female and male mice were able to remain on rotarod at RPMs that are 15% and 19% higher than wt mice, respectively (p_genotype_<0.0001 by 2-way ANOVA) (Fig. 6e).

**Figure 6.**
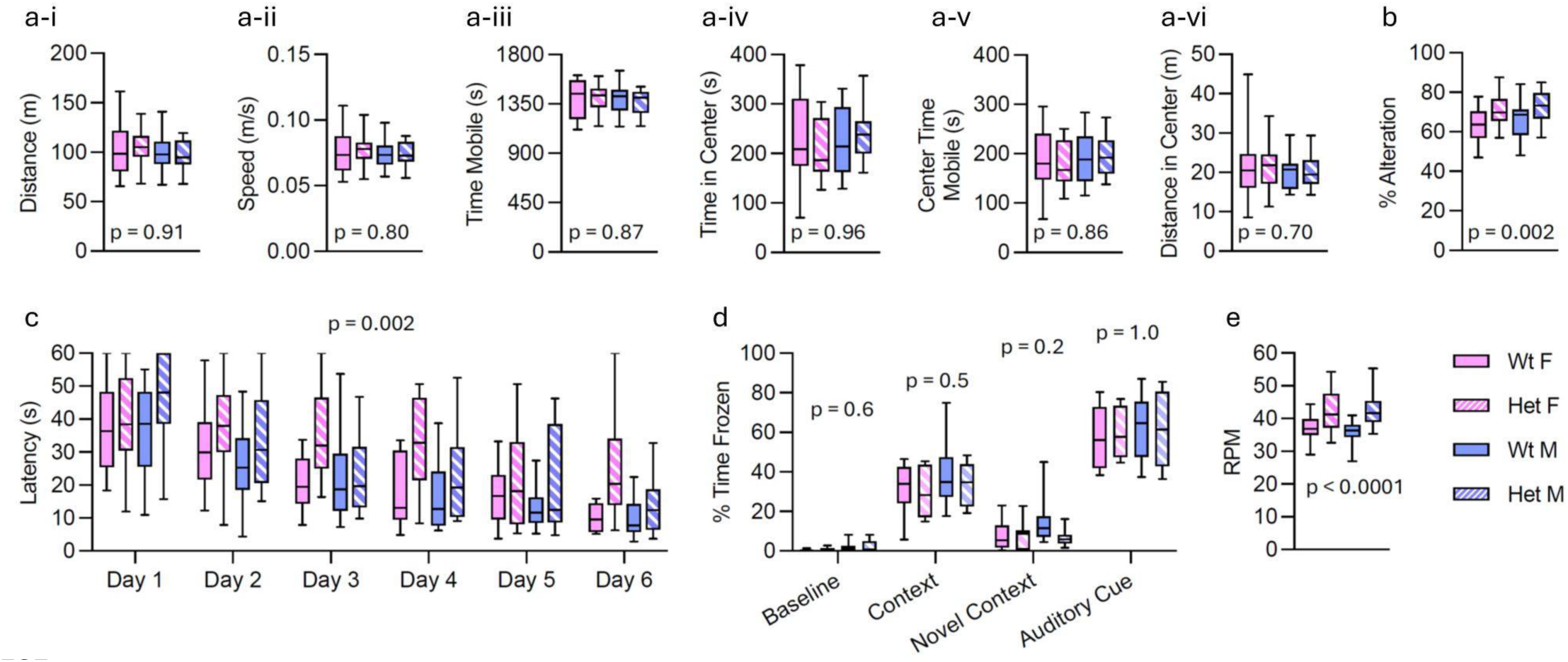
Analysis of adult behavior in *Wapl^+/+^* and *Wapl^+/-^* 5- to 7-month mice. **a**. Open Field Test, to assess mobility and exploratory behaviors. Total distance (a-i), speed (a-ii), time mobile (a-iii), time in the center region (a-iv), time immobile in the center region (a-v), and time immobile in the edge region (a-v) during the 30-minute test. Statistics are p_genotype_ from 2-way ANOVA. **b.** Y-Maze. *Wapl*^+/-^ female and male mice alternated 11% and 10% more than wt cohorts. Statistic is p_genotype_ from 2-way ANOVA. Additional metrics of this assay are shown in Fig. S14a. **c**. Morris Water Maze, to assess the ability of mice to use and remember visual clues to navigate to a hidden platform. Latency (time to platform) is measured on 6 consecutive days. *Wapl*^+/-^ mice performed significantly worse. Statistic is p_genotype_ from 3-way ANOVA. See Supplemental Fig. 14b for full summary of ANOVA analyses. Performance is especially poor in female *Wapl*^+/-^ (see Fig. S14c-d). **d**. Fear Conditioning. On day 1, we trained mice to associate environmental cues – context and an auditory signal – with a mild electric foot shock. After 24 hours, mice were re-exposed to the original context and then subsequently to the auditory cue in a novel context and their resultant freezing behaviors were used to evaluate associative learning. No significant differences in freezing were measured. Statistics are p_genotype_ from 2-way ANOVA. **e**. Rotarod. Het female and male mice performed 15% and 19% better that wt cohorts. Statistic is p_genotype_ from 2-way ANOVA. Data in (a-e) are presented as median, the 25-75% quartiles, and max/min values. In (a-c), N = 64: 16 *Wapl*^+/+^ females; 16 *Wapl*^+/-^ females; 16 *Wapl*^+/+^ males; 16 *Wapl*^+/-^ males. In (d), N = 32: 8 *Wapl*^+/+^ females; 8 *Wapl*^+/-^ females; 8 *Wapl*^+/+^ males; 8 *Wapl*^+/-^ males. In (e), N = 53: 12 *Wapl*^+/+^ females; 13 *Wapl*^+/-^ females; 15 *Wapl*^+/+^ males; 13 *Wapl*^+/-^ males.

The hippocampus plays important roles in spatial navigation and reference memory. Hippocampal lesions have been shown to result in poor MWM performance. To investigate hippocampi in our mice, we performed live MRI imaging on 3 *Wapl*^+/+^ and 6 *Wapl*^+/-^ adult female mice (Fig. S15a-b). We saw small decreases in total brain, cortex, or hippocampal volumes in hets (only whole brain volume was significant; p = 0.03 by t-test), without changes in morphology of these structures. Similar in magnitude were changes in body weight: adult *Wapl^+/-^* mice are 4-5% smaller than their wt cohorts (Fig. S13; statistics in figure).

In summary, adult *Wapl^+/-^* mice have equivalent mobility, visual acuity, anxiety/exploratory behavior, and brain morphology. They have mildly decreased brain volume and their neonatal lower body weight persists. They have poorer learning/long-term memory as measured by the MWM but behave like wt on the fear conditioning tests. Adult heterozygotes have superior motor coordination and spatial short-term memory/novelty preference as measured by the rotarod test and the Y maze, respectively.

### Wapl dosage and cardiovascular function

A minority of patients in our *WAPL* case series, and of those with 10q deletions, have cardiovascular defects. To assess heart function in mice, we performed echocardiograms on 32 mice (Table S11). We did not see evidence of the septal defects or patent ductus arteriosus observed in some *WAPL^+/-^* patients. We did observe 3 animals with mild pathologies. One wt male and one het female had reduced heart function as measured by reduced ejection fraction (54% and 53%, respectively, vs. 63% average for all mice). A second het female showed mildly elevated blood flow velocity through the left ventricular outflow tract, consistent with minor obstruction.

Comparing the frequency of pathologies between genotypes, Fisher’s exact test statistic value = 1. We used 2-way ANOVA to compare means for each measurement. Although *Wapl^+/-^* mice were significantly different for several measurements including left ventricular (LV) diastolic volume (p_genotype_ = 0.02), LV diastolic diameter (p_genotype_ = 0.02), LV posterior wall thickness at systole (p_genotype_ = 0.04), and aorta diameters at systole (p = 0.0004) and diastole (p = 0.04), the magnitude of difference between wt and mutant mice was generally modest.

### Reducing Wapl dosage to 25% results in discrete defects in embryonic development

We previously generated a hypomorphic *Wapl* allele, *Wapl^Flox^*(Kean et al., 2022). *Wapl^Flox/-^* compound heterozygotes produce Wapl protein at 25% of normal levels instead of the 50% levels produced in *Wapl^+/-^* animals. *Wapl^Flox/-^* mice die perinatally and so cannot be assayed for postnatal neuromuscular development or learning. To screen for developmental programs that are most sensitive to *Wapl* gene dosage, we analyzed e18.5 *Wapl^Flox/-^*embryos by micro-CT scanning, comparing with *Wapl^+/+^* and *Wapl^+/-^*cohorts. *Wapl^+/-^* embryos, consistent with their survival and mostly wt-like phenotypes, showed no overt dysmorphology. *Wapl^+/-^* embryos are slightly smaller but are at the appropriate developmental stage. Development of *Wapl^Flox/-^*is also relatively normal and embryos are at the appropriate developmental stage. However, we did observe three discrete phenotypes. In 5 of 6 embryos, lungs were small and alveolar maturation delayed (Fig. S15c). In 3 embryos, the right kidney, but not the right ureter, was missing (N = 2) or very small (N = 1) (Fig. S15d). These same three mice also displayed underdeveloped/missing medullar facial nucleus (Fig. S15e). In summary, there appears to be a dosage liability threshold for birth defects between 25% and 50% of wt *Wapl* level in mice.

### Genomic evidence of disease gene candidacy for cohesin release factors

To augment the targeted case series analyses and experimental approaches above, we sought large-scale population- and disease-cohort evidence to determine evidence for, or against, disease association for these cohesin release factors *WAPL*, *PDS5A*, and *PDS5B* with broader NDD phenotypes. Dosage sensitivity scores (pHaplo, pTriplo; Collins Cell 2022), as well as their correlations with measures of loss-of-function constraint (LOEUF) and missense constraint (MPC) from the genome aggregation database (gnomAD; (Chao et al., 2024; Karczewski et al., 2020)), suggest that cohesin release factor genes are broadly intolerant to functional alteration (Table S1, Fig. 1b-c). From these analyses, *WAPL* (LOEUF = 0.04; missense constraint Z = 4.27; pHaplo = 0.99, pTriplo = 0.95) was highly constrained, whereas other release complex subunits *PDS5A* (LOEUF = 0.33, missense constraint Z = 2.69; pHaplo = 0.99, pTriplo = 1) and *PDS5B* (LOEUF = 0.33, missense constraint Z = 2.35; pHaplo = 0.97, pTriplo = 0.94) were slightly more tolerant to alteration. Furthermore, *WAPL* is within the 10q22.3q23.2 GD region, a dosage-sensitive genomic locus.

The most ubiquitous feature of cohesinopathies, as well as the 10q22.3q23.2 GD, is neurodevelopmental impairment. We therefore surveyed a series of large-scale sequencing datasets for an association between variants in cohesin release factor genes and NDDs (Table S2). Specifically, we assessed a broad collection of individuals with developmental disorders (“DDD”, n = 31,058 case trios) (Kaplanis et al., 2020) and a large cohort of neurodevelopmental disorder cases (“NDD”) diagnosed with autism, some of whom have comorbid intellectual disability or developmental delay (“ID/DD”) (n = 38,680 affected trios, unpublished). Analyses included family-based and case-control study designs that incorporated all variant classes (PTVs, damaging missense, CNVs) into an integrated Bayesian association framework (Fu et al., 2022; He et al., 2013). Applying this model to the combined DDD/NDD data (n = 64,789 affected trios) identified a statistically significant association for *WAPL* (FDR = 5.14e-3), but not for *PDS5A* (FDR = 1.67e-1) or *PDS5B* (FDR = 3.38e-1). As known cohesinopathies are associated with additional features including birth defects, seizures, and behavioral issues, we performed similar analyses on aggregated data of schizophrenia (Singh et al., 2022), bipolar disorder (Palmer et al., 2022), and epilepsy (Epi25k Collaborative, 2024) (Table S2). None of these analyses identified a statistically significant enrichment of variants in the cohesin release factor genes (Table S2). Given the association of *WAPL* with NDDs, we also assessed genomic metrics to infer the potential mutational intolerance of *WAPL* as compared to other genes within the ∼7.8 Mb recurrent genomic disorder region at 10q22.3q23.2. Among all genes in the region, only two displayed evidence of substantial constraint (LOEUF ≤ 0.1; *WAPL* and *BMPR1A*), and *WAPL* represented the only gene to achieve genome-wide significant association with NDDs in the DDD cohort (p = 7.35e-4). These findings nominate *WAPL* as a driver of neurodevelopmental phenotypes in patients with 10q22.3q23.2 deletions.

These data collectively confirm our case series results that *WAPL*, but not *PDS5A* or *PDS5B*, is associated with autosomal dominant developmental disorders and a potential driver gene within the 10q genomic disorder region.

## Discussion

Genome organization is an essential nuclear property with implications across biological functions and scale (Misteli, 2020). The cohesin complex exemplifies both mechanistic control of chromosome topology and – via cataloged mutations in its component genes – the phenomenon of genome disorganization as a potential cause of disease (Piche et al., 2019). Cohesin is loaded and removed from chromatin in a cyclical manner. Yet, the relevance to human disease of cohesin release factors is unknown.

### Variants in WAPL define a syndrome of cohesin release factor deficiency

We investigated whether damaging variation in the genes encoding cohesin release factors, which play an opposing role to the cohesin loader/extrusion factor (and quintessential cohesinopathy gene) *NIPBL*, may also be disease genes. Phenotyping individuals with heterozygous predicted damaging variants in *WAPL* revealed a WAPL-related syndrome, which we propose to provisionally label “WAPL deficiency syndrome” (WDS). This condition is characterized principally by mild-moderate developmental delay/intellectual disability, but also in some individuals by craniofacial dysmorphism, organ malformations including clubfoot and mild congenital heart lesions, and other variable features. *WAPL* variants underlying the above cases were concentrated in C-terminal regions of the protein, suggesting the relative importance of C-terminal *WAPL* isoforms and the C-terminal WAPL domain. This is interesting to consider given differing reported roles for C- versus N-terminal regions of WAPL (Hara et al., 2014) and coincides with the C-terminal portion being more deeply conserved and apparently more structured (Brandes et al., 2023; Gao et al., 2023; Ouyang et al., 2013). Supporting the above, we found that heterozygous, predicted damaging variants in *WAPL* are statistically enriched among individuals with developmental disorders and neurodevelopmental delay.

WAPL participates in meiotic chromosome structuring/segregation (Crawley et al., 2016), however *WAPL* heterozygosity appears compatible with human reproduction given observed paternal and maternal inheritance in at least one case each. Immune defects and cancer did not seem to be enriched despite WAPL having roles in DNA damage repair (Benedict et al., 2020; Misulovin et al., 2018) and V(D)J recombination (Dai et al., 2021; Hill et al., 2020). Despite the opposing biological roles of their protein products, WDS is milder than and neither a mirror nor a mimic of classic *NIPBL*-related Cornelia de Lange syndrome. Incompletely penetrant features could suggest WDS is to some extent an error of developmental robustness (in other words, that *WAPL* dosage sensitivity inflection for these phenotypes in humans is located at or near hemizygosity (Green et al., 2017), and that WAPL haploinsufficiency could potentially unmask the effect of other genetic factors. Finally, we demonstrated genomic and phenotypic evidence of *WAPL* being a driver gene within the 10q22.3q23.2 reciprocal genomic disorder region.

### Phenotypes in subjects with PDS5A and PDS5B variants

While WAPL, PDS5A, and PDS5B are all involved in cohesin release, their specific roles are more nuanced. WAPL removes interphase cohesin from chromatin, acting to decrease cohesin’s residence time on chromatin, limit loop length, and guide cell-type specific gene regulation (Busslinger et al., 2017; Gassler et al., 2017; Haarhuis et al., 2017; Kiefer et al., 2023; Kueng et al., 2006; Liu et al., 2021; Tedeschi et al., 2013; Wutz et al., 2017). PDS5A and PDS5B (which in some species are a single protein, PDS5) serve as adaptors between WAPL and cohesin, yet they are not solely cohesin release factors (Zhang et al., 2021). PDS5 also inhibits cohesin ATPase activity (Murayama and Uhlmann, 2014; Petela et al., 2018), is involved in recognizing convergent CTCF proteins as boundary elements (Wutz et al., 2017), and recruits ESCO1 and ESCO2 which promote acetylation of the core cohesin protein SMC3 (Chan et al., 2013; Deardorff et al., 2012) to induce cohesin pausing and loop size restriction. Finally, PDS5 can be bound competitively by sororin (CDCA5), and PDS5-sororin has the opposite effect of PDS5-WAPL: maintenance of cohesin on chromatin (Morales et al., 2020; Nishiyama et al., 2010; Wutz et al., 2017). The net effect of PDS5 loss on genome organization, and whether redundant and/or differential roles exist for PDS5A versus PDS5B, are still being established (Al-Jomah et al., 2020; Carretero et al., 2013; Couturier et al., 2016; Cuadrado et al., 2022; Losada et al., 2005; Meisenberg et al., 2019; Ouyang et al., 2016; van Ruiten et al., 2022; Waizenegger et al., 2000; Wutz et al., 2017; Zhang et al., 2021).

In our study, subjects with variants in *PDS5A* or *PDS5B* possessed considerable morbidity, and no healthy individual has been described in the literature with a predicted damaging variant in these genes. Some trends emerged in our cases including neurodevelopmental and neurological features. Yet, the number of *PDS5A* and *PDS5B* cases was smaller than that of *WAPL* cases, and their features less convincingly converged into a unified Mendelian syndrome. Although speculative, it is possible that different missense variants confer different phenotypes given distinct roles of several PDS5 domains (Zhang et al., 2021); as a potential example of this, ‘Additional PDS5B case 4’ and ‘Additional PDS5B case 5’ (Table S4) were included because these cases involving severe neurodevelopmental and other features and share the same, rare *PDS5B* variant. Finally, damaging variants in these genes were not associated with any phenotype in our cohort-based approaches.

### Differential gene expression in CRISPR disease models

Various knockdown or knockout models of cohesin genes and studies of patient-derived cell lines or tissue have been undertaken. However, no engineered cellular models exist with the appropriate zygosity (heterozygous), species (human), and cell type (neuronal) to replicate mechanisms of brain disease in human cohesinopathies. Each of these aspects may be critically important for modeling human disease; for example, knockdown experiments where the target gene is not at heterozygous ploidy may yield different results if they fall at a different point along a dosage sensitivity inflection curve for that gene (Nadig et al., 2025). We thus used CRISPR to create heterozygous models in human iPSCs and iNs for *WAPL^+/-^* and 10q dels and dups.

RNA-seq demonstrated fewer DEGs in *WAPL* models than in published models of *NIPBL* loss, which is in line with WAPL deficiency syndrome being milder than classic CdLS. We determined pathways enriched among *WAPL* and 10q DEGs that could be relevant to disease biology, for example cell-cell adhesion in neuronal models. Finally, we found that the 10q del expression signature appeared to be partially driven by *WAPL*.

We found no evidence of expression buffering (i.e. dosage compensation) of any gene in the 10q22.3q23.2 interval, in the context of deletion or duplication of this region. Of note, LoF variants in *BMPR1A,* a gene which falls within the recurrent 10q region, cause a dominant polyposis syndrome (OMIM *601299). Yet, patients with 10q deletions are not reported to have polyposis unless they house larger deletions also eliminating the tumor suppressor gene *PTEN*, a gene beyond the recurrent deletion’s boundaries (OMIM #612242). A potential explanation for this phenomenon could be that *BMPR1A* exhibits dosage compensation when deleted; yet, our data argue against this hypothesis, at least in the cell lines we modeled (average *BMPR1A* expression in 10q del iPSCs decreased by 48% of wt and in iNs by 47%).

### Wapl heterozygous mice display only mild neuromuscular phenotypes

Phenotyping of *Wapl*^+/-^ mice was done to consider the extent to which these mice model human WAPL deficiency. More broadly, this was done to inform on organism-level dosage sensitivity of Wapl and the therapeutic window of cohesin balance in mice. Overall, *Wapl* appears to be less haploinsufficient in C57BL/6J mice than in humans where, for example, many subjects had motor delays. To further probe dosage sensitivity in mouse, we generated *Wapl^Flox/-^* animals that express 25% of wt levels of Wapl. This genotype is postnatal lethal and these mice exhibit some birth defects. The substantially milder phenotypes of *Wapl*^+/-^ mice and *WAPL*^+/-^ humans, compared to *Nipbl*^+/-^ or *NIPBL*^+/-^, not only informs on dosage sensitivity of these genes, but also may indicate a wider range in which *WAPL* could be perturbed during therapy for NIPBL-related disease (Kean et al., 2022; Luppino et al., 2022) while still resulting in a net improvement in phenotype. However, the *Wapl^Flox/-^* mice provide a potential caution regarding the bounds of such a range.

### Limitations and future possibilities

The penetrance and expressivity of *WAPL* variants will be clarified by future studies. Additional studies are needed to confirm disease associations of variants in cohesin release factor genes, and human cell models of *PDS5A* and *PDS5B* genes are needed to further elucidate these genes’ shared vs disparate and redundant vs independent roles. Further mechanistic studies of cohesin balance, cycling, and effect on the 3D genome in our models are needed to confirm pathomechanistic connections linking mutation, genome disorganization, gene misexpression, and disease. The roles of cohesin and WAPL during human neurodevelopment are of particular interest, both for informing on brain development and critical temporal windows for treating cohesinopathies.

### Summary

In summary, *WAPL* is a novel haploinsufficient disease gene associated with neurodevelopmental phenotypes, risk of congenital anomalies, and is a likely driver gene for the 10q recurrent deletion syndrome. Variants in *PDS5A* and *PDS5B*, which encode binding partners of WAPL, are found in patients with intellectual disability and/or other neurodevelopmental features, but we did not find convincing evidence that these genes underlie clinically homogeneous Mendelian syndromes. Disease modeling of WAPL deficiency syndrome in stem cell and neuronal cell lines allowed transcriptional effects of this condition – and its relation to the 10q genomic disorder – to be deciphered. The critical phenotypic threshold for *Wapl* in mice appears to lie between 25 and 50% dosage. Overall, we highlight that cohesin balance is bidirectionally dosage sensitive, as loss of cohesin loader or releaser causes human disease.

## Data Availability

All data produced in the present study are available upon reasonable request to the authors.

## Acknowledgements

This work was supported by the following grants, programs, and philanthropy: The United States NIH NIGMS (T32GM007748-41, GM061354), NINDS (K08NS117891), NHGRI (U01HG011755, U01HG011744), NSF GRFP (DGE1745303), NICHD (HD081256), and NIMH (R01MH115957); the Massachusetts General Hospital Eckerd Scholars Program and Center for Diversity and Inclusion SRTP Program; the Czech National Center for Medical Genomics (LM2018132), MH CZ DRO Motol (00064203) and NW24-06-00083; the Italian Ministry of Health (RF-2021-12374963 and PNRR-MR1-2022-12376811); the Boston Children’s Hospital (BCH) Rare Disease Collaborative; donation from Peter and Kathryn Wagner; donation from Julie and Frank Mairano; the BCH Office of Faculty Development/Basic & Clinical Translational Research Executive Committees; the government of Canada through Genome Canada and the Ontario Genomics Institute (OGI-188) awarded to B.S.; the German Research Foundation (DFG; RI 3503/3-1) awarded to K.M.R.; the Postdoc Mobility Fellowship from the Swiss National Science Foundation (P500-3_235131) to C.A.; and the São Paulo Research Foundation (FAPESP) (2022/03980-5 and 2025/00171-7) and Coordination for the Improvement of Higher Education Personnel (CAPES) (88887.113708/2025-00) to L.M.L.C. and by FASEP (2023/09879-7) to A.C.V.K. The SouthSeq project (U01HG007301) is supported by the Clinical Sequencing Evidence-Generating Research (CSER) consortium which is funded by the National Human Genome Research Institute (NHGRI) with co-funding from the National Institute on Minority Health and Health Disparities (NIMHD) and the National Cancer Institute (NCI). The authors thank: the patients and families who contributed to this study; Drs. Holly Harris, Caroline Dias, and Angela Lin for suggestions regarding phenotyping; Annabelle Tuttle, Allyson McCrary, Drs. Isabella Herman, Jennifer Posey, Moez Dawood, Emmi Helle, Jessica Chong, Michael Bamshad, Sue Moyer Harasink, Karen Gripp, Colby Marvin, Eva Schwaibold, Kathyrn Shively, Gabrielle Lemire, Maya Chopra, Kati Buckingham, Emily Wai Yi Cheng, Fowzan Alkuraya, Charlotte W. Ockeloen, Julia Foreman, Laura Orec, Pengfei Liu, Jason Flannick, and Shira Rockowitz for assistance with cases or identifying cases; Wenqi Shi and Adela Ye for suggestions regarding statistical tests; Dr. Judith Kassis for general advice; the CdLS Foundation USA and members of the BCH CdLS and Related Disorders Clinic for clinical support; Drs. Dennis Lal, AJ Campbell, and Sumaiya Iqbal for advice about 3D protein modeling; and Dr. Gabi Xavier, Maris Handley, and Pathik Sen for experimental assistance. This study makes use of data generated by the DECIPHER community (Firth et al., 2009). A full list of centers who contributed to the generation of the data is available from http://decipher.sanger.ac.uk and via email from. Funding for DECIPHER was provided by the Wellcome Trust. Those who carried out the original DECIPHER analysis and collection of the data bear no responsibility for the further analysis or interpretation of the data. This study makes use of data from the DDD study (Deciphering Developmental Disorders, 2015). The DDD was commissioned by the UK Health Innovation Challenge Fund.

## Declaration of generative AI and AI-assisted technologies in the manuscript preparation process

During the preparation of this work the authors used Google Gemini v2.0 Flash in order to generate HPO terms from lists of phenotypes. After using this tool/service, the authors reviewed and edited the content as needed and take full responsibility for the content of the published article.

## Methods

### Human subjects research

We enrolled patients, and obtained data, including genetic and phenotypic information, and biological samples under an IRB-approved research protocol at Boston Children’s Hospital (00040134) and under relevant research protocols at the collaborating institutions listed in Table S4. All procedures adhered to the principles of the Declaration of Helsinki and its later amendments.

### Identifying WAPL, PDS5A, PDS5B, and 10q22.3q23.2 cases

We queried multiple databases (Table S3) including GeneMatcher (Sobreira et al., 2015) and MatchMaker Exchange (Philippakis et al., 2015) to identify, recruit, and phenotype individuals with predicted damaging variants in *WAPL*, *PDS5A*, or *PDS5B*. Missense variants were included if predicted damaging/deleterious or likely/probably/possibly damaging by two or more of the following three tools: SIFT, PolyPhen, and AlphaMissense as obtained via the Ensembl Variant Effect Predictor (https://ensembl.org/info/docs/tools/vep/index.html). MPC scores (Chao et al., 2024) are also included in Table S4 but were not used to exclude cases. Additional cases were excluded because a variant affecting another gene could potentially explain the phenotype (n=3), or because the variant was present in >10 individuals in gnomAD (n=2), although information about these cases is retained in Table S4. Table S4 also lists two cases that do not fulfil the pathogenicity criteria above, because they shared the same rare *PDS5B* variant (c.523A>G p.Met175Val) and severe phenotypes. The phenotype of published patients with 10q deletions was identified by literature review.

### DNA methylation signature analysis

We applied EpiSign^TM^ analysis to establish a genome-wide DNA methylation episignature. All case subjects were de-identified and the research was conducted in accordance with relevant institutional approvals and ethical standards. The study followed the protocols of the Western University Research Ethics Board (REB 106302 and 116108). The methylation analysis followed a previously established protocol (Aref-Eshghi et al., 2020). In summary, DNA was extracted from the peripheral blood of individuals carrying *WAPL*, *PDS5A*, and *PDS5B* SNVs, and 10q dels, as well as from control subjects. After bisulfite conversion, the DNA samples were processed using Illumina Infinium MethylationEPIC BeadChip microarrays (San Diego, CA), which assess approximately 860,000 CpG sites across the human genome. Methylated and unmethylated signal intensities were analyzed using R v4.3.1, with normalization conducted via the SeSAMe package (v1.20.0; (Zhou et al., 2018)). Several probe types were excluded from further analysis, including those located on chromosomes X and Y, those containing SNPs near CpG interrogation or single-nucleotide extension sites, cross-reactive probes that bind to non-target regions, and probes flagged by Illumina for poor technical performance. Samples were also excluded if > 5% of their probes failed or if they were previously identified in the EpiSign Knowledge Database (EKD, https://episign.lhsc.on.ca/index.html) as being prone to batch effects. All remaining samples displayed typical bimodal distributions in genome-wide methylation density. Principal component analysis (PCA) was conducted to assess batch structure and detect any potential outliers, which resulted in the exclusion of additional samples as noted below.

Data analysis strategy: Since analyzing all study samples did not yield a consistent DNA methylation profile due to cohort heterogeneity, three separate analyses were conducted. In Analysis 1, 14 samples with truncating variants affecting *WAPL* (including 10q del), *PDS5A*, or *PDS5B* were included for methylation profiling. In Analysis 2, 10 samples with either *WAPL* SNVs or 10q deletions were analyzed. In Analysis 3, eight samples with *WAPL* SNV samples were used for profiling.

Selection of matched control samples: Using the MatchIt package (version 4.5.5; Ho, Imai et al Polit Anal 2007), control samples were randomly selected from the EKD, matched to the case samples based on age, sex, and array type. For Analyses 1, 2, and 3, sets of 56, 50, and 56 control samples were selected, respectively. After each round of control selection, PCA was conducted on the combined set of case and matched control samples. Any control samples identified as outliers in the first two principal components were excluded. This process was repeated iteratively until no additional outliers were detected.

Methylation profiling: For each probe, the methylation level (β-value) was calculated by dividing the intensity of the methylated signal by the combined intensity of both methylated and unmethylated signals. This β-value ranges from 0 (indicating no methylation) to 1 (indicating complete methylation). These β-values were then converted into M-values using a logit transformation prior to conducting linear regression analysis with the limma package (v3.58.1) (Ritchie et al., 2015). The analysis accounted for variations in blood cell type composition, which were estimated using Houseman’s algorithm (Houseman et al., 2012). Estimated cell proportions were included as covariates in the model matrix for the linear model. The p-values obtained from the model were moderated using the eBayes function in limma. For Analysis 2, multiple testing correction was performed using the Benjamini-Hochberg (BH) procedure, while for Analysis 1 and 3, non-adjusted p-values were utilized.

Probe selection and verification: To identify the cases with the most pronounced methylation differences and to select probes that best distinguish case samples from controls, the following method was used. Probes were ranked in descending order based on the product of the mean methylation difference and the negative logarithm of the p-value. From this ranking, the top 800 probes were selected. Of these, a subset with the highest areas under the receiver operating characteristic (ROC) curve—specifically 400, 400, and 267 probes for Analyses 1, 2, and 3, respectively—was chosen. To reduce redundancy, within each set, only one probe was kept from pairs showing high similarity (Pearson’s correlation coefficient greater than 0.6 for Analyses 1 and 2, and 0.65 for Analysis 3). In Analyses 1, 2, and 3, the number of selected probes was 211, 268, and 200, respectively. To evaluate the robustness of these selected probes, both hierarchical clustering using Ward’s method with Euclidean distances and multidimensional scaling (MDS) based on Euclidean distances were performed. To test the reproducibility of the methylation profiles, a leave-one-out cross-validation (LOOCV) approach was applied. In each iteration, one case sample was excluded, probes were reselected using the remaining samples, and an MDS plot was created to determine whether the excluded sample clustered with the remaining case samples and their matched controls.

Construction of the binary classifier: To improve the accuracy of subject classification, a binary support vector machine (SVM) was developed using the e1071 package. This model produces methylation variant pathogenicity (MVP) scores ranging from 0 to 1 for each sample, with higher scores indicating a closer resemblance to the identified episignature. The SVM was constructed by training the case samples against their matched controls, 75% of additional control samples, and 75% of individuals with other rare genetic disorders included in the EpiSign v5 clinical classifier from the EKD (Levy et al., 2022). The remaining 25% of these samples were used to test the model’s performance. MVP scores were also calculated for the remaining study samples that were not involved in probe selection or model training.

### Exome burden analysis

Exome burden analysis was performed based on two datasets: A published exome sequencing study of probands with developmental disorders (“DDD”, 31,058 case trios) for which single-nucleotide variant (SNV) and insertion/deletion (indel) *de novo* mutation counts were available (Kaplanis et al., 2020) and the latest freeze (unpublished) of exome sequencing data of neurodevelopmental disorder cases (“NDD”, 38,680 case trios and 23,749 isolated cases matched with 23,749 control samples) from the Autism Sequencing Consortium (ASC) (Buxbaum et al., 2012), for which *de novo*, inherited, and case-control SNV/indel and copy-number variants (CNVs) were available. The latter represents an aggregate of samples ascertained for autism with or without co-occurring intellectual disability or developmental delay (“ID/DD”) stemming from different sources with distinct ascertainment criteria (from self-reported to clinical diagnosis) and study designs (trios vs isolated cases). Details of data collection, alignment, and variant calling will be described in detail in a forthcoming manuscript describing this new freeze. Trios present in both the DDD and NDD datasets were identified and deduplicated, totaling 64,789 trio cases across both datasets.

Evidence for the association between genetic variation in a gene and a phenotype was obtained through the Bayesian transmission and de novo association (TADA) framework that incorporates evidence from different study designs and variant classes (Fu et al., 2022; He et al., 2013). To do so, SNV/indels were stratified into five classes: i) protein-truncating variants (PTVs) identified as high-confidence loss-of-function or “other splice” (OS) variants by LOFTEE (Karczewski et al., 2020); ii-iv) three classes of missense variants depending on whether the variant’s Missense deleteriousness Prediction by Constraint (MPC) (Chao et al., 2024) and AlphaMissense pathogenicity (AM) (Cheng et al., 2023) were deemed damaging (i.e., MPC > 2 and AM > 0.07), so that Mis2 variants met both criteria, Mis1 variants met one of the criteria, and Mis0 met none of the criteria; v) synonymous variants, identified by the Variant Effect Predictor (VEP) (McLaren et al., 2016). CNVs were stratified into deletions and duplications using previously described criteria (Fu et al., 2022). Bayes factors for each study design (*de novo*, inherited, case-control) and variant class (PTV, Mis2, Mis1, deletion, duplication) were computed and integrated, allowing to derive a single gene-level posterior probability used to compute a false discovery rate (FDR), as previously described (Fu et al., 2022). Significance threshold for evidence of an association between damaging variants in a gene and a phenotype was defined at FDR < 0.001, which has previously been shown to approximate exome-wide Bonferroni significance threshold (*P* < 2.8 × 10^−6^) (Fu et al., 2022).

### rCNV analyses

The rCNV data was derived from microarrays, and thus the breakpoint coordinates of CNVs are not exact with base pair resolution. CNVs were restricted to those ≥ 100kb. Coordinates of the 10q22.3q23.2 reciprocal genomic disorder region was lifted over from chr10:79,788,887-87,311,071 (hg38) to chr10:81,548,643-89,070,828 (hg19), and individuals in the rCNV dataset carrying an overlapping (≥80% reciprocal overlap) CNV were identified. Cases were defined as individuals with any HPO term. The count of total cases and controls was from (Collins Cell 2022). Odds ratios and associated P values were calculated using the two-sided Fisher test comparing case proportion to the entire rCNV cohort. Among the 65 10q del or dup carriers, all but one overlaps (≥1bp) *WAPL* (chr10:88,195,013-88,281,549; NCBI hg19).

To explore the HPO annotation for the 57 case 10q del and dup carriers, we utilized the HPO database, version October 22^nd^, 2025. This was done by counting the number of times a given HPO term appears listed under the phenotype description of a carrier, across all 10q del/dup carriers. There were a total of 37 unique HPO terms across CNV carriers. For plotting, we excluded “phenotypic abnormality” (HP:0000118), which was present in 30 deletion carriers and 17 duplication carriers, as it is an ancestor term and is too general. The remaining 36 terms were annotated with their 3^rd^ degree ancestor, which is used to categorize individual HPO terms.

### iPSC culture

Cell culture and other laboratory work were performed under MassGeneralBrigham biosafety protocol 2011B000500. For CRISPR experiments, we utilized an extensively characterized induced pluripotent stem cell (iPSC) line from a healthy human donor (GM08330) (https://coriell.org/0/Sections/Search/Sample_Detail.aspx?Ref=GM08330&Product=CC) (Sheridan et al., 2011). Prior to editing, the line was confirmed to have a 46, XY karyotype (via WiCell) and no pathogenic variants were detected via whole genome sequencing (not shown). iPSCs were cultured on Matrigel (Corning). Media was Essential 8 (E8) (Gibco) with 1% penicillin-streptomycin (Fisher Scientific). ROCK inhibitor (Y-27632 dihydrochloride; Biological Industries) was supplemented at 10uM for 24h after thawing or passaging and during FACS sorting. iPSCs were passaged with ReleSR (STEMCELL Technologies) and cryopreserved with mFreSR (STEMCELL Technologies) or NutriFreez D10 (Biological Industries). Mycoplasma testing with the V1 EZ-PCR protocol (Biological Industries) was negative (not shown).

### CRISPR gene editing

sgRNAs (IDT) for making fs indels (Table S12) were designed using a combination of the IDT CRISPR-Cas9 guide RNA design checker (https://www.idtdna.com/site/order/designtool/index/CRISPR_SEQUENCE) and the UCSC Genome Browser CRISPR targets track (https://genome.ucsc.edu/). sgRNAs for making 10q CNVs (Table S12) were designed based upon the principles of SCORE (Tai et al., 2016) as previously described (Nuttle et al., 2024). iPSCs were edited with ribonucleoproteins (RNPs) consisting of sgRNAs complexed with Alt-R S.p. HiFi Cas9 nuclease V3 (IDT) in duplex buffer (IDT) (see http://sfvideo.blob.core.windows.net/sitefinity/docs/default-source/user-guide-manual/alt-r-crispr-cas9-user-guide-ribonucleoprotein-transfections-recommended.pdf?sfvrsn=1c43407_8). RNPs were co-transfected with EGFP mRNA (CleanCap; TriLink Biotechnologies) using Lipofectamine STEM (Thermo-Fisher) diluted in Opti-MEM I (Gibco). Cells were sorted as single cells via the FACSAria system (BD), gated on GFP to enrich for transfected cells, at 48h post-transfection.

### Genotyping and pluripotency selection

Genotyping of iPSCs was performed at the time of post-FACS consolidation and repeated at the time of initial freezing of clonal lines. Frameshift (fs) indels were genotyped via PCR and Sanger sequencing as in (Boone et al., 2020). Edits were identified with assistance from the ICE CRISPR analysis tool (https://www.synthego.com/products/bioinformatics/crispr-analysis). Genotyping at post-FACS consolidation, demonstrated high editing efficiency for fs indels (67 wt/wt clones, 30 fs/wt, 7 in-frame indel/wt, 31 fs/in-frame indel, 7 in-frame/in-frame, 4 fs/fs); none of these fs/fs lines survived to initial freezing. Equal numbers of +1 register fs and -1 register fs clones were carried forward. 10q dels and dups were genotyped via droplet digital PCR as in (Boone et al., 2020), with a Thermo-Fisher Taq-Man target probe (ID Hs00599013_cn). Primers and the ddPCR amplicon are listed in Table S12. Genotyping results are shown in Fig. S5 and listed in Table S7. *WAPL* fs indels identified by Sanger sequencing of DNA were further confirmed via variant calling of RNA-seq data.

Edited iPSCs were enriched for the TRA-1-60 pluripotency marker via magnetic cell sorting on the MACS system (Miltenyi) prior to harvesting for RNA or neuronal differentiation. Six clones harboring an independently edited variant and six unedited clones per locus of interest were enriched, with the exception of 10q dups (g2B5); as dups are rare events, a single edited line was triplicated prior to pluripotency enrichment to derive biological pseudo-replicates (i.e. g2B5_1/2/3; Table S7). Each of these three replicates was subsequently duplicated to generate six lines (i.e. g2B5_1A/1B/2A/2B/3A/3B; Table S7). In the case of iPSCs, this duplication was done prior to RNA collection. In the case of iNs, this duplication was done following *ngn2* transduction. After pluripotency enrichment, iPSCs were transferred to Geltrex (Gibco) coated plates and harvested when they reached confluency 3 days later in TRIzol (Thermo Fisher) or as frozen cell pellets.

### Neuronal differentiation

Glutamatergic induced neurons (iNs) were generated via a doxycycline responsive lentiviral-mediated *Ngn2* transgene approach (Shi et al., 2012; Zhang et al., 2013) as described previously (Tai et al., 2022), with key timepoints as follows: iPSCs were transduced with rtTA-and pTetO-mNgn2-encoding lentiviruses on day -8. On day 0, E8 medium was switched to Neuronal Maintenance Medium (NMM) supplemented with doxycycline (Sigma-Aldrich) to start the induction. After 24 hours, iPSCs were re-plated with Accutase (Thermo Fisher) and selection was started using puromycin (Thermo Fisher) for 72 hours. On day 4, cells were counted and re-plated at 1 million cells per 6-well, onto poly-L-ornithine (Sigma-Aldrich) and laminin (Sigma) coated plates in NMM supplemented with doxycycline (Sigma-Aldrich) until Day 10 and NT-3 (Preprotech) and BDNF (Prospec Bio) until collection. Ara-C (Sigma) was added on day 5 and half medium changes were performed every three days. Morphology was consistent with successful differentiation in all cases. Harvest for RNA was at differentiation day 20 (10q) or 24 (*WAPL*).

### RNA

RNA was extracted from TRIzol with chloroform in Phasemaker tubes (Thermo Fisher), precipitated with isopropanol and RNAse-free glycogen (Roche), washed with 75% ethanol, quantitated using a Nanodrop spectrophotometer (Thermo), and QC’ed on a TapeStation 4200 (Agilent). cDNA was prepared using SuperScript IV reverse transcriptase (Thermo Fisher) and oligo (dT)_15_ (Promega), according to the SuperScript protocol (https://assets.thermofisher.com/TFS-Assets%2FLSG%2Fmanuals%2FSSIV_First_Strand_Synthesis_System_UG.pdf). qPCR was performed in technical triplicate or duplicate (consistent within an experiment) with SYBR Green I Master mix (Roche) and the LightCycler 480 instrument (Roche). A dilution series was included for each experiment in order to calculate reaction efficiency (Pfaffl, 2001). The thermocycler used the following program: 95°C x 5’ ramp rate 4.4°C/s; then 35 cycles of 95°C x 10s (ramp rate 4.4), 55C × 20s (ramp rate 2.2), and 72°C x 30s (ramp rate 4.4). qPCR primers are listed in Table S12. Statistics are t-tests based on (Livak and Schmittgen, 2001), using average primer efficiencies when calculating 2^-deltadeltaCt.

Total RNA-seq libraries were prepared using the v1 NEBNext rRNA Depletion Kit (NEB) according to the manufacturer’s instructions. Briefly, RNA samples were hybridized with rRNA depletion probes, digested with RNase H and DNase I, and fragmented using divalent cations and higher temperature. Random primers primed first-strand cDNA synthesis, followed by second-strand cDNA synthesis with dUTP replacing dTTP. Libraries were end-repaired, A-tailed, universal hairpin loop adapter ligated, and USER enzyme digested. Libraries were indexed via 11-cycles of PCR amplification and unique dual-indexed primer pairs. QC was undertaken on a TapeStation 4200 (Agilent) and qPCR with a Library Quantification Kit (Kapa Biosystems). Finally, libraries were pooled for sequencing on an Illumina NovaSeq6000 using an S4 300 cycles kit.

### RNA-seq data analysis

A total of 60 RNA sequencing (RNA-seq) libraries was generated from 30 human induced pluripotent stem cells (iPSC) and 30 *Ngn2*-induced neurons (iNs) and sequenced on an Illumina NovaSeq platform. The sequencing process produced an average of 44.87 million paired-end reads of 150 base pairs. Quality checking of the raw sequence reads was assessed by fastQC (version 0.10.1) (https://www.bioinformatics.babraham.ac.uk/projects/fastqc/). Adapter sequences and low-quality bases were trimmed from the raw sequence reads using Trimmomatic (version 0.36) with the following parameters: ILLUMINACLIP:adapter.fa:2:30:10 LEADING:3 TRAILING:3 SLIDINGWINDOW:4:20 MINLEN:50 (Bolger et al., 2014). After trimming, the average number of paired-end reads per library was reduced to 43.25 million. Trimmed sequence reads were aligned to the human reference genome (GRCh38, Ensembl build 92) using STAR (version 2.5.3) with the following parameters: --twopassMode Basic --outFilterMultimapNmax 1 --outFilterMismatchNoverLmax 0.05 --outSAMtype BAM Unsorted --outReadsUnmapped Fastx --readFilesCommand zcat --alignIntronMin 21 --alignIntronMax 0 --quantMode GeneCounts --alignEndsType EndToEnd. This process also generated gene-level counts for each library based on the human gene annotations for the same human reference genome (Dobin et al., 2013). Alignment quality was further assessed using custom scripts incorporating PicardTools (version 1.75) (https://broadinstitute.github.io/picard/), RNASeqQC (version 1.1.7) (DeLuca et al., 2012), RSeQC (version 2.3.3) (Wang et al., 2012) and SamTools (version 0.1.18) (Li et al., 2009). Differential expression analyses were separately performed for the following comparisons in iPSCs and iNs: *WAPL^+/-^* vs wt, 10q del vs wt, 10q dup vs wt. In each comparison, genes with at least 0.5 count-per-million (CPM) expression in at least 50% of the RNA-seq libraries were retained for further analyses. The CPM for gene *i* in library *j* was calculated using the following formula: 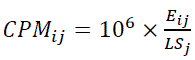, where *E*_ij_ represents the raw count expression of gene *i* in library *j*, and *LS_j_* is the library size of library j, which was defined as the number of uniquely mapped reads reported by STAR aligner.

Allelic expression: To validate the effect of CRISPR-induced edits into target genes, we checked the allelic expression of the reference and mutant alleles. To do so, we first computed the number of reads that support reference and alternate alleles at the edit site identified by FreeBayes (Garrison and Marth arXiv 2012) and the number of reads at the target location in control samples. To make the latter comparable with reference and alternate allele expression, we divided the number of reads in control samples by two. Next, we normalized these reads by the number of uniquely mapped reads retrieved from the STAR log file and multiplied it by 1 million (equivalent to CPM calculation). DE analysis: Differential expression analysis was performed between edited samples and control samples using DESeq2 (Love et al., 2014) with SVA (Leek et al., 2012) ∼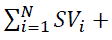, where SVs are surrogate variables (SVs) estimated by R/Bioconductor package SVA to remove unwanted variation in the data using ∼ genotype as the full model and ∼1 as the reduced model.

Genotype group reassignment: In both iPSC and iN, *WAPL* P1F10, thought to be an unedited sample based on Sanger genotyping of PCRed DNA, was found to have a hemizygous expression level of *WAPL*. Thus, this sample was reassigned as *WAPL*^+/-^, for all analyses. Variant calling from RNA-seq failed to definitively identify the identity of the suspected LoF variant in P1F10 (one of 63 reads showed a 10nt deletion in iPSCs; Table S7). To identify the outlying samples in each comparison, we first calculated Pearson correlation coefficients for each sample with other samples in the comparison using the DESeq2-normalized count data after identifying the genes that pass the expression threshold (CPM=0.5). Next, average Pearson correlation coefficient for each sample was calculated and then z-transformed (scaled). Samples with z ≤ 3 were identified as outliers. Following this approach, *WAPL*^+/-^ sample P1B5 and 10q del sample g1B11 were identified as an outlying sample and thus removed from *WAPL*^+/-^ iN RNA-seq analyses and 10q deliPSC RNA-seq analyses, respectively. Furthermore, these samples clustered away from other samples sharing their respective genotype on PCA.

Pathway enrichment: We performed pathway enrichment analysis of selected DEGs using one-tailed Fisher’s exact test for the curated lists of Gene Ontology (GO) terms and REACTOME terms from mSigDB (version 2025.1.Hs) (Liberzon et al., 2015). In this analysis, the list of genes that pass the expression threshold (CPM=0.5) in a given comparison was used as the background set for that comparison, and the pathway terms with at least 10 associated genes in the background list were analyzed. Multiple testing correction following Benjamini-Hochberg procedure was applied to the analyses of distinct pathway types (e.g. GO: Biological Process, REACTOME) for each comparison.

Overlap enrichment: Significance of overlapping DEGs between different comparisons was assessed using one-tailed Fisher’s exact test.

All RNA-seq related statistical analyses were performed using R Statistical Software (version 4.3.2).

### Immunoblotting

Total protein was isolated from frozen cell pellets using RIPA lysis buffer (Boston BioProducts) and cOmplete Protease Inhibitor Cocktail (Roche). Lysates were quantitated with the Pierce BCA Protein Assay Kit (Thermo Fisher). Lysates were prepared in NuPAGE LDS sample buffer containing 1X reducing agent and run on NuPAGE 3-8% tris-acetate gels (Thermo Fisher), followed by transfer to a 0.45um Immobilon-P PVDF membrane (Millipre Sigma) in 1X bis-tris transfer buffer (Boston BioProducts) containing 20% methanol. Membranes were blocked in Intercept blocking buffer (LICORbio) diluted 1:1 with PBS. Primary and secondary antibodies, diluted in 1:1 Intercept blocking buffer and TBST, are listed in Table S12. Proteins were visualized on an Odyssey CLx scanner (LICORbio) and quantitated using Image Studio 6.1 software (LICORbio).

### Mouse models

All mouse studies were performed according to NIH and PHS guidelines and only after protocols were reviewed and approved by the Eunice Kennedy Shriver National Institute of Child Health and Human Development Animal Care and Use Committee. Mice carrying the *Wapl* null allele (*Wapl*^-^) and the Wapl hypomorph allele (*Wapl*^Flox^) were generated as described (Kean et al., 2022). *Wapl*^-^ carries a 4,158 bp del that removes the *Wapl* promoter and exons 1 and 2. These *Wapl* alleles were generated on a 129X1/SvJ background and backcrossed >12 generations to C57BL/6J (strain 000664, Jackson labs) to generate the mice used in this study. Genotypes were determined by PCR analysis of gDNAs extracted from ear punch or tail biopsies as previously described (Kean et al., 2022). As *Wapl^flox^* decreases the expression of *Wapl* (Kean et al., 2022), *Wapl^+/+^* were used as wt controls. The four groups in each experiment were: wt females, het females, wt males, het males. Number of mice/group is indicated in figure legends.

### Mouse anatomic phenotyping

Embryo micro-CT: e18.5 embryos were collected into PBS with 20,000 U heparin/liter, transferred to cold PBS with 4% Paraformaldehyde, incubated at least 4 days with gentle rocking, and transferred to PBS with 0.02% sodium azide. Micro-CT scans were performed and data analyzed by the University of California Mouse Biology Program Embryo Phenotyping Core. Approximately 100 embryo tissues and organs were assessed on the frontal, sagittal, and transverse planes. Adult brain MRI: Brain MRIs were performed on 8-9-month-old mice using a 9.4-T, 30 cm horizontal Bruker Avance scanner (Bruker BioSpin, Billerica) operating on a PV360 platform. Each mouse was anesthetized in 1.5% isoflurane and its head positioned in an 86 mm transmit and a cryo cooled, 2x2 array, receive coil ensemble and centered in the scanner. We acquired 18 axial images (0.8 mm) of T2 weighted fast spin echo (Repetition Time [TR]/Echo Time [TE] = 2500/8.25 ms, in-plane resolution = 75 µm, number of averages (NA) = 8, echo train = 8) images encompassing the whole brain. We referenced coronal sections of adult mouse brains published by the Allen Brain Atlas to determine brain morphology. The MR images were segmented in ITK-SNAP to calculate volumes of the whole brain, cerebral cortex, and hippocampus. Adult echocardiography: Transthoracic echocardiography was performed on 14-month-old mice as described (Park et al., 2021). Data were analyzed by 2-way ANOVA using GraphPad Prism Software.

### Mouse neonatal motor/behavioral phenotyping

p2 to p20 neonates were assessed daily in hours 10-12 of a 14-hour light cycle by a single investigator blinded to genotype and following previously described protocols (Feather-Schussler and Ferguson, 2016). Righting: Righting was assessed by placing pups in a prone position, releasing them, and recording the time required to turn all 4 paws onto the ground surface. Three trials of maximum 30 seconds were conducted, and times averaged. Geotaxis: Geotaxis (or positional sensing) was assessed by placing pups facing downward on a 45° slope, releasing them, and recording the time to turn 90° and turn 180°. Three trials of maximum 30 seconds were conducted, and times averaged. Falls were recorded but not used in calculating performance. Forepaw grasp reflex: Forepaw grasping reflex was tested by holding each pup by the nape of the neck and brushing each forepaw with the blunt side of a straight-edge razor blade. Gait: Gait was assayed qualitatively using the following scoring system: 0 = no movement; 1 = crawling with asymmetric limb movement; 2 = slow crawling but symmetric limb movement; 3 = walking. Four Limb Hang: Muscle strength was assessed via the Four Limb Hang assay, starting at p5. Mice were placed on a fiberglass screen. After 3 seconds, the screen was slowly rotated and the angle at which the mouse slid off was recorded. Weight: Mice were weighed daily.

### Mouse adult motor/behavioral phenotyping

Adult behavioral and motor screening was performed in 5–7-month-old mice. Rotarod performance was assessed in hours 4-7 of a 14-hour light cycle. Visual acuity was assessed in hours 10-12 of a 14-hour light cycle. All other behavioral analyses were assessed starting at hour 2 of a 12-hour dark cycle. Open Field Test: Mice are removed from their home cage and individually placed into a 16” x 16” x 16” Perspex arena viewing chamber and allowed to explore for 30 minutes. Movements were recorded and analyzed using ANY-maze (Wood Dale, IL). The edge zone is defined as all areas inside the arena within 10 cm of any of the four chamber walls. The center is defined as any area not within the edge zone. Spontaneous Y maze: The Y maze has three arms separated by 120° angles. Two arms are 15.25 cm long x 12.7 cm high x 7.62 cm wide and a third arm is 20.32 cm long and with a start box at the end. The mouse was placed in the start box, the door opened, and the mouse was allowed to freely explore the three arms for 5 minutes. Movements were recorded and analyzed using ANY-maze. Morris Water Maze (MWM): The MWM test was performed in a circular pool (4 feet diameter, 30” high) (San Diego Instruments). Water was made opaque by adding non-toxic white paint (Crayola) and kept at 20°-30°C. A small plexiglass platform is hidden in the NW quadrant of the pool at 2 cm below the surface. Visual clues are placed around the pool to enable mice a chance to learn the platform location. Mice received 4 trials per day (60 seconds maximum per trial) over six consecutive days. Swim distance, swim speed, path length to platform, and latency to platform were analyzed with ANY-maze. After learning trials were completed on day 6, the platform was removed for a 90 second probe trial and the number of crossings over the location previously occupied by the platform and the times spent in each quadrant were assessed. Fear Conditioning: Mice were placed individually in a sound-attenuating chamber, 17 cm x 17 cm x 25 cm. Baseline freezing was measured for 120 sec. Auditory signals (2 x 4 kHz, dB tone) were delivered into the chamber for 15s, followed by a 2-sec foot shock (0.85 mA). After a break of 120 sec, the tone-shock procedure was repeated. Then the mouse was returned to its home cage. On day 2 (hour 24), the mouse was returned to the original chamber and contextual freezing was recorded. At hour 25, the mouse was placed in a new chamber and novel context freezing was measured for 120 seconds. Then auditory cues were applied and freezing is measured. Recording and analyses were done using ANY-maze software. Rotarod Performance: Motor coordination was assessed using a rotarod test (Ugo Basile SRL, Italy). Two to four mice were tested simultaneously with males and females always tested separately. Each mouse underwent training trials and then a test that lasted a maximum of 3 minutes as the rod accelerated from 2 to 60 rpm (acceleration = 19.33 rpm/min). Each test was repeated three times with a 40-minute rest interval between. Visual Acuity: Visual acuity was assessed using an Optodrum^PLUS^ (Stria.Tech) using Stria.Tech software. Contrast was locked at 99.72%. Both the parameters and test frequencies were selected by the Stria.Tech software. A trial was successful if the mouse passed a certain threshold for head-tracking. A mouse “passed” at a test frequency after two successful trials or “failed” after three unsuccessful trials. The highest pass was recorded as the value for the visual acuity.

## References

1. Al-Jomah, N., Mukololo, L., Anjum, A., Al Madadha, M., Patel, R., 2020. Pds5A and Pds5B Display Non-redundant Functions in Mitosis and Their Loss Triggers Chk1 Activation. Front Cell Dev Biol 8, 531.

2. Aref-Eshghi, E., Kerkhof, J., Pedro, V.P., Groupe, D.I.F., Barat-Houari, M., Ruiz-Pallares, N., Andrau, J.C., Lacombe, D., Van-Gils, J., Fergelot, P., Dubourg, C., Cormier-Daire, V., Rondeau, S., Lecoquierre, F., Saugier-Veber, P., Nicolas, G., Lesca, G., Chatron, N., Sanlaville, D., Vitobello, A., Faivre, L., Thauvin-Robinet, C., Laumonnier, F., Raynaud, M., Alders, M., Mannens, M., Henneman, P., Hennekam, R.C., Velasco, G., Francastel, C., Ulveling, D., Ciolfi, A., Pizzi, S., Tartaglia, M., Heide, S., Heron, D., Mignot, C., Keren, B., Whalen, S., Afenjar, A., Bienvenu, T., Campeau, P.M., Rousseau, J., Levy, M.A., Brick, L., Kozenko, M., Balci, T.B., Siu, V.M., Stuart, A., Kadour, M., Masters, J., Takano, K., Kleefstra, T., de Leeuw, N., Field, M., Shaw, M., Gecz, J., Ainsworth, P.J., Lin, H., Rodenhiser, D.I., Friez, M.J., Tedder, M., Lee, J.A., DuPont, B.R., Stevenson, R.E., Skinner, S.A., Schwartz, C.E., Genevieve, D., Sadikovic, B., 2020. Evaluation of DNA Methylation Episignatures for Diagnosis and Phenotype Correlations in 42 Mendelian Neurodevelopmental Disorders. Am J Hum Genet 106, 356–370.

3. Azidane, S., Gallego, X., Durham, L., Caceres, M., Guney, E., Perez-Cano, L., 2024. Identification of novel driver risk genes in CNV loci associated with neurodevelopmental disorders. HGG Adv 5, 100316.

4. Balciuniene, J., Feng, N., Iyadurai, K., Hirsch, B., Charnas, L., Bill, B.R., Easterday, M.C., Staaf, J., Oseth, L., Czapansky-Beilman, D., Avramopoulos, D., Thomas, G.H., Borg, A., Valle, D., Schimmenti, L.A., Selleck, S.B., 2007. Recurrent 10q22-q23 deletions: a genomic disorder on 10q associated with cognitive and behavioral abnormalities. Am J Hum Genet 80, 938–947.

5. Ball, A.R., Jr., Chen, Y.Y., Yokomori, K., 2014. Mechanisms of cohesin-mediated gene regulation and lessons learned from cohesinopathies. Biochim Biophys Acta 1839, 191–202.

6. Benedict, B., van Schie, J.J.M., Oostra, A.B., Balk, J.A., Wolthuis, R.M.F., Riele, H.T., de Lange, J., 2020. WAPL-Dependent Repair of Damaged DNA Replication Forks Underlies Oncogene-Induced Loss of Sister Chromatid Cohesion. Dev Cell 52, 683–698 e687.

7. Bolger, A.M., Lohse, M., Usadel, B., 2014. Trimmomatic: a flexible trimmer for Illumina sequence data. Bioinformatics 30, 2114–2120.

8. Boone, P.M., Paterson, S., Mohajeri, K., Zhu, W., Genetti, C.A., Tai, D.J.C., Nori, N., Agrawal, P.B., Bacino, C.A., Bi, W., Talkowski, M.E., Hogan, B.M., Rodan, L.H., 2020. Biallelic mutation of FBXL7 suggests a novel form of Hennekam syndrome. Am J Med Genet A 182, 189–194.

9. Brandes, N., Goldman, G., Wang, C.H., Ye, C.J., Ntranos, V., 2023. Genome-wide prediction of disease variant effects with a deep protein language model. Nat Genet 55, 1512–1522.

10. Breckpot, J., Tranchevent, L.C., Thienpont, B., Bauters, M., Troost, E., Gewillig, M., Vermeesch, J.R., Moreau, Y., Devriendt, K., Van Esch, H., 2012. BMPR1A is a candidate gene for congenital heart defects associated with the recurrent 10q22q23 deletion syndrome. Eur J Med Genet 55, 12–16.

11. Busslinger, G.A., Stocsits, R.R., van der Lelij, P., Axelsson, E., Tedeschi, A., Galjart, N., Peters, J.M., 2017. Cohesin is positioned in mammalian genomes by transcription, CTCF and Wapl. Nature 544, 503–507.

12. Buxbaum, J.D., Daly, M.J., Devlin, B., Lehner, T., Roeder, K., State, M.W., Autism Sequencing, C., 2012. The autism sequencing consortium: large-scale, high-throughput sequencing in autism spectrum disorders. Neuron 76, 1052–1056.

13. Carretero, M., Ruiz-Torres, M., Rodriguez-Corsino, M., Barthelemy, I., Losada, A., 2013. Pds5B is required for cohesion establishment and Aurora B accumulation at centromeres. EMBO J 32, 2938–2949.

14. Carvalho, C.M., Zhang, F., Lupski, J.R., 2010. Evolution in health and medicine Sackler colloquium: Genomic disorders: a window into human gene and genome evolution. Proc Natl Acad Sci U S A 107 Suppl 1, 1765–1771.

15. Chan, K.L., Gligoris, T., Upcher, W., Kato, Y., Shirahige, K., Nasmyth, K., Beckouet, F., 2013. Pds5 promotes and protects cohesin acetylation. Proc Natl Acad Sci U S A 110, 13020–13025.

16. Chan, K.L., Roig, M.B., Hu, B., Beckouet, F., Metson, J., Nasmyth, K., 2012. Cohesin’s DNA exit gate is distinct from its entrance gate and is regulated by acetylation. Cell 150, 961–974.

17. Chao, K.R., Wang, L., Panchal, R., Liao, C., Abderrazzaq, H., Ye, R., Schultz, P., Compitello, J., Grant, R.H., Kosmicki, J.A., Weisburd, B., Phu, W., Wilson, M.W., Laricchia, K.M., Goodrich, J.K., Goldstein, D., Goldstein, J.I., Vittal, C., Poterba, T., Baxter, S., Watts, N.A., Solomonson, M., gnom, A.D.C., Tiao, G., Rehm, H.L., Neale, B.M., Talkowski, M.E., MacArthur, D.G., O’Donnell-Luria, A., Karczewski, K.J., Radivojac, P., Daly, M.J., Samocha, K.E., 2024. The landscape of regional missense mutational intolerance quantified from 125,748 exomes. bioRxiv.

18. Cheng, J., Novati, G., Pan, J., Bycroft, C., Zemgulyte, A., Applebaum, T., Pritzel, A., Wong, L.H., Zielinski, M., Sargeant, T., Schneider, R.G., Senior, A.W., Jumper, J., Hassabis, D., Kohli, P., Avsec, Z., 2023. Accurate proteome-wide missense variant effect prediction with AlphaMissense. Science 381, eadg7492.

19. Ciosk, R., Shirayama, M., Shevchenko, A., Tanaka, T., Toth, A., Shevchenko, A., Nasmyth, K., 2000. Cohesin’s binding to chromosomes depends on a separate complex consisting of Scc2 and Scc4 proteins. Mol Cell 5, 243–254.

20. Coelho Molck, M., Simioni, M., Paiva Vieira, T., Paoli Monteiro, F., Gil-da-Silva-Lopes, V.L., 2017. A New Case of the Rare 10q22.3q23.2 Microdeletion Flanked by Low-Copy Repeats 3/4. Mol Syndromol 8, 161–167.

21. Collins, R.L., Glessner, J.T., Porcu, E., Lepamets, M., Brandon, R., Lauricella, C., Han, L., Morley, T., Niestroj, L.M., Ulirsch, J., Everett, S., Howrigan, D.P., Boone, P.M., Fu, J., Karczewski, K.J., Kellaris, G., Lowther, C., Lucente, D., Mohajeri, K., Noukas, M., Nuttle, X., Samocha, K.E., Trinh, M., Ullah, F., Vosa, U., Epi, C., Estonian Biobank Research, T., Hurles, M.E., Aradhya, S., Davis, E.E., Finucane, H., Gusella, J.F., Janze, A., Katsanis, N., Matyakhina, L., Neale, B.M., Sanders, D., Warren, S., Hodge, J.C., Lal, D., Ruderfer, D.M., Meck, J., Magi, R., Esko, T., Reymond, A., Kutalik, Z., Hakonarson, H., Sunyaev, S., Brand, H., Talkowski, M.E., 2022. A cross-disorder dosage sensitivity map of the human genome. Cell 185, 3041–3055 e3025.

22. Couturier, A.M., Fleury, H., Patenaude, A.M., Bentley, V.L., Rodrigue, A., Coulombe, Y., Niraj, J., Pauty, J., Berman, J.N., Dellaire, G., Di Noia, J.M., Mes-Masson, A.M., Masson, J.Y., 2016. Roles for APRIN (PDS5B) in homologous recombination and in ovarian cancer prediction. Nucleic Acids Res 44, 10879–10897.

23. Crawley, O., Barroso, C., Testori, S., Ferrandiz, N., Silva, N., Castellano-Pozo, M., Jaso-Tamame, A.L., Martinez-Perez, E., 2016. Cohesin-interacting protein WAPL-1 regulates meiotic chromosome structure and cohesion by antagonizing specific cohesin complexes. Elife 5, e10851.

24. Cuadrado, A., Gimenez-Llorente, D., De Koninck, M., Ruiz-Torres, M., Kojic, A., Rodriguez-Corsino, M., Losada, A., 2022. Contribution of variant subunits and associated factors to genome-wide distribution and dynamics of cohesin. Epigenetics Chromatin 15, 37.

25. Cuadrado, A., Gimenez-Llorente, D., Kojic, A., Rodriguez-Corsino, M., Cuartero, Y., Martin-Serrano, G., Gomez-Lopez, G., Marti-Renom, M.A., Losada, A., 2019. Specific Contributions of Cohesin-SA1 and Cohesin-SA2 to TADs and Polycomb Domains in Embryonic Stem Cells. Cell Rep 27, 3500–3510 e3504.

26. Dai, H.Q., Hu, H., Lou, J., Ye, A.Y., Ba, Z., Zhang, X., Zhang, Y., Zhao, L., Yoon, H.S., Chapdelaine-Williams, A.M., Kyritsis, N., Chen, H., Johnson, K., Lin, S., Conte, A., Casellas, R., Lee, C.S., Alt, F.W., 2021. Loop extrusion mediates physiological Igh locus contraction for RAG scanning. Nature 590, 338–343.

27. Davidson, I.F., Bauer, B., Goetz, D., Tang, W., Wutz, G., Peters, J.M., 2019. DNA loop extrusion by human cohesin. Science 366, 1338–1345.

28. Deardorff, M.A., Bando, M., Nakato, R., Watrin, E., Itoh, T., Minamino, M., Saitoh, K., Komata, M., Katou, Y., Clark, D., Cole, K.E., De Baere, E., Decroos, C., Di Donato, N., Ernst, S., Francey, L.J., Gyftodimou, Y., Hirashima, K., Hullings, M., Ishikawa, Y., Jaulin, C., Kaur, M., Kiyono, T., Lombardi, P.M., Magnaghi-Jaulin, L., Mortier, G.R., Nozaki, N., Petersen, M.B., Seimiya, H., Siu, V.M., Suzuki, Y., Takagaki, K., Wilde, J.J., Willems, P.J., Prigent, C., Gillessen-Kaesbach, G., Christianson, D.W., Kaiser, F.J., Jackson, L.G., Hirota, T., Krantz, I.D., Shirahige, K., 2012. HDAC8 mutations in Cornelia de Lange syndrome affect the cohesin acetylation cycle. Nature 489, 313–317.

29. Deciphering Developmental Disorders, S., 2015. Large-scale discovery of novel genetic causes of developmental disorders. Nature 519, 223–228.

30. DeLuca, D.S., Levin, J.Z., Sivachenko, A., Fennell, T., Nazaire, M.D., Williams, C., Reich, M., Winckler, W., Getz, G., 2012. RNA-SeQC: RNA-seq metrics for quality control and process optimization. Bioinformatics 28, 1530–1532.

31. Dobin, A., Davis, C.A., Schlesinger, F., Drenkow, J., Zaleski, C., Jha, S., Batut, P., Chaisson, M., Gingeras, T.R., 2013. STAR: ultrafast universal RNA-seq aligner. Bioinformatics 29, 15–21.

32. Epi25k Collaborative, 2024. Exome sequencing of 20,979 individuals with epilepsy reveals shared and distinct ultra-rare genetic risk across disorder subtypes. Nat Neurosci 27, 1864–1879.

33. Feather-Schussler, D.N., Ferguson, T.S., 2016. A Battery of Motor Tests in a Neonatal Mouse Model of Cerebral Palsy. J Vis Exp.

34. Firth, H.V., Richards, S.M., Bevan, A.P., Clayton, S., Corpas, M., Rajan, D., Van Vooren, S., Moreau, Y., Pettett, R.M., Carter, N.P., 2009. DECIPHER: Database of Chromosomal Imbalance and Phenotype in Humans Using Ensembl Resources. Am J Hum Genet 84, 524–533.

35. Fu, J.M., Satterstrom, F.K., Peng, M., Brand, H., Collins, R.L., Dong, S., Wamsley, B., Klei, L., Wang, L., Hao, S.P., Stevens, C.R., Cusick, C., Babadi, M., Banks, E., Collins, B., Dodge, S., Gabriel, S.B., Gauthier, L., Lee, S.K., Liang, L., Ljungdahl, A., Mahjani, B., Sloofman, L., Smirnov, A.N., Barbosa, M., Betancur, C., Brusco, A., Chung, B.H.Y., Cook, E.H., Cuccaro, M.L., Domenici, E., Ferrero, G.B., Gargus, J.J., Herman, G.E., Hertz-Picciotto, I., Maciel, P., Manoach, D.S., Passos-Bueno, M.R., Persico, A.M., Renieri, A., Sutcliffe, J.S., Tassone, F., Trabetti, E., Campos, G., Cardaropoli, S., Carli, D., Chan, M.C.Y., Fallerini, C., Giorgio, E., Girardi, A.C., Hansen-Kiss, E., Lee, S.L., Lintas, C., Ludena, Y., Nguyen, R., Pavinato, L., Pericak-Vance, M., Pessah, I.N., Schmidt, R.J., Smith, M., Costa, C.I.S., Trajkova, S., Wang, J.Y.T., Yu, M.H.C., Autism Sequencing, C., Broad Institute Center for Common Disease, G., i, P.-B.C., Cutler, D.J., De Rubeis, S., Buxbaum, J.D., Daly, M.J., Devlin, B., Roeder, K., Sanders, S.J., Talkowski, M.E., 2022. Rare coding variation provides insight into the genetic architecture and phenotypic context of autism. Nat Genet 54, 1320–1331.

36. Gabriele, M., Brandao, H.B., Grosse-Holz, S., Jha, A., Dailey, G.M., Cattoglio, C., Hsieh, T.S., Mirny, L., Zechner, C., Hansen, A.S., 2022. Dynamics of CTCF- and cohesin-mediated chromatin looping revealed by live-cell imaging. Science 376, 496–501.

37. Gandhi, R., Gillespie, P.J., Hirano, T., 2006. Human Wapl is a cohesin-binding protein that promotes sister-chromatid resolution in mitotic prophase. Curr Biol 16, 2406–2417.

38. Gao, H., Hamp, T., Ede, J., Schraiber, J.G., McRae, J., Singer-Berk, M., Yang, Y., Dietrich, A.S.D., Fiziev, P.P., Kuderna, L.F.K., Sundaram, L., Wu, Y., Adhikari, A., Field, Y., Chen, C., Batzoglou, S., Aguet, F., Lemire, G., Reimers, R., Balick, D., Janiak, M.C., Kuhlwilm, M., Orkin, J.D., Manu, S., Valenzuela, A., Bergman, J., Rousselle, M., Silva, F.E., Agueda, L., Blanc, J., Gut, M., de Vries, D., Goodhead, I., Harris, R.A., Raveendran, M., Jensen, A., Chuma, I.S., Horvath, J.E., Hvilsom, C., Juan, D., Frandsen, P., de Melo, F.R., Bertuol, F., Byrne, H., Sampaio, I., Farias, I., do Amaral, J.V., Messias, M., da Silva, M.N.F., Trivedi, M., Rossi, R., Hrbek, T., Andriaholinirina, N., Rabarivola, C.J., Zaramody, A., Jolly, C.J., Phillips-Conroy, J., Wilkerson, G., Abee, C., Simmons, J.H., Fernandez-Duque, E., Kanthaswamy, S., Shiferaw, F., Wu, D., Zhou, L., Shao, Y., Zhang, G., Keyyu, J.D., Knauf, S., Le, M.D., Lizano, E., Merker, S., Navarro, A., Bataillon, T., Nadler, T., Khor, C.C., Lee, J., Tan, P., Lim, W.K., Kitchener, A.C., Zinner, D., Gut, I., Melin, A., Guschanski, K., Schierup, M.H., Beck, R.M.D., Umapathy, G., Roos, C., Boubli, J.P., Lek, M., Sunyaev, S., O’Donnell-Luria, A., Rehm, H.L., Xu, J., Rogers, J., Marques-Bonet, T., Farh, K.K., 2023. The landscape of tolerated genetic variation in humans and primates. Science 380, eabn8153.

39. Garcia, P., Fernandez-Hernandez, R., Cuadrado, A., Coca, I., Gomez, A., Maqueda, M., Latorre-Pellicer, A., Puisac, B., Ramos, F.J., Sandoval, J., Esteller, M., Mosquera, J.L., Rodriguez, J., Pie, J., Losada, A., Queralt, E., 2021. Disruption of NIPBL/Scc2 in Cornelia de Lange Syndrome provokes cohesin genome-wide redistribution with an impact in the transcriptome. Nat Commun 12, 4551.

40. Gassler, J., Brandao, H.B., Imakaev, M., Flyamer, I.M., Ladstatter, S., Bickmore, W.A., Peters, J.M., Mirny, L.A., Tachibana, K., 2017. A mechanism of cohesin-dependent loop extrusion organizes zygotic genome architecture. EMBO J 36, 3600–3618.

41. Green, R.M., Fish, J.L., Young, N.M., Smith, F.J., Roberts, B., Dolan, K., Choi, I., Leach, C.L., Gordon, P., Cheverud, J.M., Roseman, C.C., Williams, T.J., Marcucio, R.S., Hallgrimsson, B., 2017. Developmental nonlinearity drives phenotypic robustness. Nat Commun 8, 1970.

42. Haarhuis, J.H.I., van der Weide, R.H., Blomen, V.A., Yanez-Cuna, J.O., Amendola, M., van Ruiten, M.S., Krijger, P.H.L., Teunissen, H., Medema, R.H., van Steensel, B., Brummelkamp, T.R., de Wit, E., Rowland, B.D., 2017. The Cohesin Release Factor WAPL Restricts Chromatin Loop Extension. Cell 169, 693–707 e614.

43. Hafner, A., Boettiger, A., 2023. The spatial organization of transcriptional control. Nat Rev Genet 24, 53–68.

44. Hara, K., Zheng, G., Qu, Q., Liu, H., Ouyang, Z., Chen, Z., Tomchick, D.R., Yu, H., 2014. Structure of cohesin subcomplex pinpoints direct shugoshin-Wapl antagonism in centromeric cohesion. Nat Struct Mol Biol 21, 864–870.

45. He, X., Sanders, S.J., Liu, L., De Rubeis, S., Lim, E.T., Sutcliffe, J.S., Schellenberg, G.D., Gibbs, R.A., Daly, M.J., Buxbaum, J.D., State, M.W., Devlin, B., Roeder, K., 2013. Integrated model of de novo and inherited genetic variants yields greater power to identify risk genes. PLoS Genet 9, e1003671.

46. Hill, L., Ebert, A., Jaritz, M., Wutz, G., Nagasaka, K., Tagoh, H., Kostanova-Poliakova, D., Schindler, K., Sun, Q., Bonelt, P., Fischer, M., Peters, J.M., Busslinger, M., 2020. Wapl repression by Pax5 promotes V gene recombination by Igh loop extrusion. Nature 584, 142–147.

47. Houseman, E.A., Accomando, W.P., Koestler, D.C., Christensen, B.C., Marsit, C.J., Nelson, H.H., Wiencke, J.K., Kelsey, K.T., 2012. DNA methylation arrays as surrogate measures of cell mixture distribution. BMC Bioinformatics 13, 86.

48. Huis in ’t Veld, P.J., Herzog, F., Ladurner, R., Davidson, I.F., Piric, S., Kreidl, E., Bhaskara, V., Aebersold, R., Peters, J.M., 2014. Characterization of a DNA exit gate in the human cohesin ring. Science 346, 968–972.

49. Kaplanis, J., Samocha, K.E., Wiel, L., Zhang, Z., Arvai, K.J., Eberhardt, R.Y., Gallone, G., Lelieveld, S.H., Martin, H.C., McRae, J.F., Short, P.J., Torene, R.I., de Boer, E., Danecek, P., Gardner, E.J., Huang, N., Lord, J., Martincorena, I., Pfundt, R., Reijnders, M.R.F., Yeung, A., Yntema, H.G., Deciphering Developmental Disorders, S., Vissers, L., Juusola, J., Wright, C.F., Brunner, H.G., Firth, H.V., FitzPatrick, D.R., Barrett, J.C., Hurles, M.E., Gilissen, C., Retterer, K., 2020. Evidence for 28 genetic disorders discovered by combining healthcare and research data. Nature 586, 757–762.

50. Karczewski, K.J., Francioli, L.C., Tiao, G., Cummings, B.B., Alfoldi, J., Wang, Q., Collins, R.L., Laricchia, K.M., Ganna, A., Birnbaum, D.P., Gauthier, L.D., Brand, H., Solomonson, M., Watts, N.A., Rhodes, D., Singer-Berk, M., England, E.M., Seaby, E.G., Kosmicki, J.A., Walters, R.K., Tashman, K., Farjoun, Y., Banks, E., Poterba, T., Wang, A., Seed, C., Whiffin, N., Chong, J.X., Samocha, K.E., Pierce-Hoffman, E., Zappala, Z., O’Donnell-Luria, A.H., Minikel, E.V., Weisburd, B., Lek, M., Ware, J.S., Vittal, C., Armean, I.M., Bergelson, L., Cibulskis, K., Connolly, K.M., Covarrubias, M., Donnelly, S., Ferriera, S., Gabriel, S., Gentry, J., Gupta, N., Jeandet, T., Kaplan, D., Llanwarne, C., Munshi, R., Novod, S., Petrillo, N., Roazen, D., Ruano-Rubio, V., Saltzman, A., Schleicher, M., Soto, J., Tibbetts, K., Tolonen, C., Wade, G., Talkowski, M.E., Genome Aggregation Database, C., Neale, B.M., Daly, M.J., MacArthur, D.G., 2020. The mutational constraint spectrum quantified from variation in 141,456 humans. Nature 581, 434–443.

51. Kean, C.M., Tracy, C.J., Mitra, A., Rahat, B., Van Winkle, M.T., Gebert, C.M., Noeker, J.A., Calof, A.L., Lander, A.D., Kassis, J.A., Pfeifer, K., 2022. Decreasing Wapl dosage partially corrects embryonic growth and brain transcriptome phenotypes in Nipbl(+/-) embryos. Sci Adv 8, eadd4136.

52. Kiefer, L., Chiosso, A., Langen, J., Buckley, A., Gaudin, S., Rajkumar, S.M., Servito, G.I.F., Cha, E.S., Vijay, A., Yeung, A., Horta, A., Mui, M.H., Canzio, D., 2023. WAPL functions as a rheostat of Protocadherin isoform diversity that controls neural wiring. Science 380, eadf8440.

53. Kueng, S., Hegemann, B., Peters, B.H., Lipp, J.J., Schleiffer, A., Mechtler, K., Peters, J.M., 2006. Wapl controls the dynamic association of cohesin with chromatin. Cell 127, 955–967.

54. Kwon, S., Safer, J., Nguyen, D.T., Hoksza, D., May, P., Arbesfeld, J.A., Rubin, A.F., Campbell, A.J., Burgin, A., Iqbal, S., 2024. Genomics 2 Proteins portal: a resource and discovery tool for linking genetic screening outputs to protein sequences and structures. Nat Methods 21, 1947–1957.

55. Leek, J.T., Johnson, W.E., Parker, H.S., Jaffe, A.E., Storey, J.D., 2012. The sva package for removing batch effects and other unwanted variation in high-throughput experiments. Bioinformatics 28, 882–883.

56. Levy, M.A., McConkey, H., Kerkhof, J., Barat-Houari, M., Bargiacchi, S., Biamino, E., Bralo, M.P., Cappuccio, G., Ciolfi, A., Clarke, A., DuPont, B.R., Elting, M.W., Faivre, L., Fee, T., Fletcher, R.S., Cherik, F., Foroutan, A., Friez, M.J., Gervasini, C., Haghshenas, S., Hilton, B.A., Jenkins, Z., Kaur, S., Lewis, S., Louie, R.J., Maitz, S., Milani, D., Morgan, A.T., Oegema, R., Ostergaard, E., Pallares, N.R., Piccione, M., Pizzi, S., Plomp, A.S., Poulton, C., Reilly, J., Relator, R., Rius, R., Robertson, S., Rooney, K., Rousseau, J., Santen, G.W.E., Santos-Simarro, F., Schijns, J., Squeo, G.M., St John, M., Thauvin-Robinet, C., Traficante, G., van der Sluijs, P.J., Vergano, S.A., Vos, N., Walden, K.K., Azmanov, D., Balci, T., Banka, S., Gecz, J., Henneman, P., Lee, J.A., Mannens, M., Roscioli, T., Siu, V., Amor, D.J., Baynam, G., Bend, E.G., Boycott, K., Brunetti-Pierri, N., Campeau, P.M., Christodoulou, J., Dyment, D., Esber, N., Fahrner, J.A., Fleming, M.D., Genevieve, D., Kerrnohan, K.D., McNeill, A., Menke, L.A., Merla, G., Prontera, P., Rockman-Greenberg, C., Schwartz, C., Skinner, S.A., Stevenson, R.E., Vitobello, A., Tartaglia, M., Alders, M., Tedder, M.L., Sadikovic, B., 2022. Novel diagnostic DNA methylation episignatures expand and refine the epigenetic landscapes of Mendelian disorders. HGG Adv 3, 100075.

57. Li, H., Handsaker, B., Wysoker, A., Fennell, T., Ruan, J., Homer, N., Marth, G., Abecasis, G., Durbin, R., Genome Project Data Processing, S., 2009. The Sequence Alignment/Map format and SAMtools. Bioinformatics 25, 2078–2079.

58. Liberzon, A., Birger, C., Thorvaldsdottir, H., Ghandi, M., Mesirov, J.P., Tamayo, P., 2015. The Molecular Signatures Database (MSigDB) hallmark gene set collection. Cell Syst 1, 417–425.

59. Liu, H., Tsai, H., Yang, M., Li, G., Bian, Q., Ding, G., Wu, D., Dai, J., 2023. Three-dimensional genome structure and function. MedComm (2020) 4, e326.

60. Liu, J., Zhang, Z., Bando, M., Itoh, T., Deardorff, M.A., Clark, D., Kaur, M., Tandy, S., Kondoh, T., Rappaport, E., Spinner, N.B., Vega, H., Jackson, L.G., Shirahige, K., Krantz, I.D., 2009. Transcriptional dysregulation in NIPBL and cohesin mutant human cells. PLoS Biol 7, e1000119.

61. Liu, N.Q., Maresca, M., van den Brand, T., Braccioli, L., Schijns, M., Teunissen, H., Bruneau, B.G., Nora, E.P., de Wit, E., 2021. WAPL maintains a cohesin loading cycle to preserve cell-type-specific distal gene regulation. Nat Genet 53, 100–109.

62. Livak, K.J., Schmittgen, T.D., 2001. Analysis of relative gene expression data using real-time quantitative PCR and the 2(-Delta Delta C(T)) Method. Methods 25, 402–408.

63. Losada, A., Yokochi, T., Hirano, T., 2005. Functional contribution of Pds5 to cohesin-mediated cohesion in human cells and Xenopus egg extracts. J Cell Sci 118, 2133–2141.

64. Love, M.I., Huber, W., Anders, S., 2014. Moderated estimation of fold change and dispersion for RNA-seq data with DESeq2. Genome Biol 15, 550.

65. Luppino, J.M., Field, A., Nguyen, S.C., Park, D.S., Shah, P.P., Abdill, R.J., Lan, Y., Yunker, R., Jain, R., Adelman, K., Joyce, E.F., 2022. Co-depletion of NIPBL and WAPL balance cohesin activity to correct gene misexpression. PLoS Genet 18, e1010528.

66. Mach, P., Kos, P.I., Zhan, Y., Cramard, J., Gaudin, S., Tunnermann, J., Marchi, E., Eglinger, J., Zuin, J., Kryzhanovska, M., Smallwood, S., Gelman, L., Roth, G., Nora, E.P., Tiana, G., Giorgetti, L., 2022. Cohesin and CTCF control the dynamics of chromosome folding. Nat Genet 54, 1907–1918.

67. McLaren, W., Gil, L., Hunt, S.E., Riat, H.S., Ritchie, G.R., Thormann, A., Flicek, P., Cunningham, F., 2016. The Ensembl Variant Effect Predictor. Genome Biol 17, 122.

68. Meisenberg, C., Pinder, S.I., Hopkins, S.R., Wooller, S.K., Benstead-Hume, G., Pearl, F.M.G., Jeggo, P.A., Downs, J.A., 2019. Repression of Transcription at DNA Breaks Requires Cohesin throughout Interphase and Prevents Genome Instability. Mol Cell 73, 212–223 e217.

69. Misteli, T., 2020. The Self-Organizing Genome: Principles of Genome Architecture and Function. Cell 183, 28–45.

70. Misulovin, Z., Pherson, M., Gause, M., Dorsett, D., 2018. Brca2, Pds5 and Wapl differentially control cohesin chromosome association and function. PLoS Genet 14, e1007225.

71. Morales, C., Ruiz-Torres, M., Rodriguez-Acebes, S., Lafarga, V., Rodriguez-Corsino, M., Megias, D., Cisneros, D.A., Peters, J.M., Mendez, J., Losada, A., 2020. PDS5 proteins are required for proper cohesin dynamics and participate in replication fork protection. J Biol Chem 295, 146–157.

72. Murayama, Y., Uhlmann, F., 2014. Biochemical reconstitution of topological DNA binding by the cohesin ring. Nature 505, 367–371.

73. Nadig, A., Replogle, J.M., Pogson, A.N., Murthy, M., McCarroll, S.A., Weissman, J.S., Robinson, E.B., O’Connor, L.J., 2025. Transcriptome-wide analysis of differential expression in perturbation atlases. Nat Genet 57, 1228–1237.

74. Nishiyama, T., Ladurner, R., Schmitz, J., Kreidl, E., Schleiffer, A., Bhaskara, V., Bando, M., Shirahige, K., Hyman, A.A., Mechtler, K., Peters, J.M., 2010. Sororin mediates sister chromatid cohesion by antagonizing Wapl. Cell 143, 737–749.

75. Nuttle, X., Burt, N.D., Currall, B., Moyses-Oliveira, M., Mohajeri, K., Bhavsar, R., Lucente, D., Yadav, R., Tai, D.J.C., Gusella, J.F., Talkowski, M.E., 2024. Parallelized engineering of mutational models using piggyBac transposon delivery of CRISPR libraries. Cell Rep Methods 4, 100672.

76. Oikawa, K., Ohbayashi, T., Kiyono, T., Nishi, H., Isaka, K., Umezawa, A., Kuroda, M., Mukai, K., 2004. Expression of a novel human gene, human wings apart-like (hWAPL), is associated with cervical carcinogenesis and tumor progression. Cancer Res 64, 3545–3549.

77. Ouyang, Z., Zheng, G., Song, J., Borek, D.M., Otwinowski, Z., Brautigam, C.A., Tomchick, D.R., Rankin, S., Yu, H., 2013. Structure of the human cohesin inhibitor Wapl. Proc Natl Acad Sci U S A 110, 11355–11360.

78. Ouyang, Z., Zheng, G., Tomchick, D.R., Luo, X., Yu, H., 2016. Structural Basis and IP6 Requirement for Pds5-Dependent Cohesin Dynamics. Mol Cell 62, 248–259.

79. Palmer, D.S., Howrigan, D.P., Chapman, S.B., Adolfsson, R., Bass, N., Blackwood, D., Boks, M.P.M., Chen, C.Y., Churchhouse, C., Corvin, A.P., Craddock, N., Curtis, D., Di Florio, A., Dickerson, F., Freimer, N.B., Goes, F.S., Jia, X., Jones, I., Jones, L., Jonsson, L., Kahn, R.S., Landen, M., Locke, A.E., McIntosh, A.M., McQuillin, A., Morris, D.W., O’Donovan, M.C., Ophoff, R.A., Owen, M.J., Pedersen, N.L., Posthuma, D., Reif, A., Risch, N., Schaefer, C., Scott, L., Singh, T., Smoller, J.W., Solomonson, M., Clair, D.S., Stahl, E.A., Vreeker, A., Walters, J.T.R., Wang, W., Watts, N.A., Yolken, R., Zandi, P.P., Neale, B.M., 2022. Exome sequencing in bipolar disorder identifies AKAP11 as a risk gene shared with schizophrenia. Nat Genet 54, 541–547.

80. Park, K.S., Rahat, B., Lee, H.C., Yu, Z.X., Noeker, J., Mitra, A., Kean, C.M., Knutsen, R.H., Springer, D., Gebert, C.M., Kozel, B.A., Pfeifer, K., 2021. Cardiac pathologies in mouse loss of imprinting models are due to misexpression of H19 long noncoding RNA. Elife 10.

81. Petela, N.J., Gligoris, T.G., Metson, J., Lee, B.G., Voulgaris, M., Hu, B., Kikuchi, S., Chapard, C., Chen, W., Rajendra, E., Srinivisan, M., Yu, H., Lowe, J., Nasmyth, K.A., 2018. Scc2 Is a Potent Activator of Cohesin’s ATPase that Promotes Loading by Binding Scc1 without Pds5. Mol Cell 70, 1134–1148 e1137.

82. Pfaffl, M.W., 2001. A new mathematical model for relative quantification in real-time RT-PCR. Nucleic Acids Res 29, e45.

83. Philippakis, A.A., Azzariti, D.R., Beltran, S., Brookes, A.J., Brownstein, C.A., Brudno, M., Brunner, H.G., Buske, O.J., Carey, K., Doll, C., Dumitriu, S., Dyke, S.O., den Dunnen, J.T., Firth, H.V., Gibbs, R.A., Girdea, M., Gonzalez, M., Haendel, M.A., Hamosh, A., Holm, I.A., Huang, L., Hurles, M.E., Hutton, B., Krier, J.B., Misyura, A., Mungall, C.J., Paschall, J., Paten, B., Robinson, P.N., Schiettecatte, F., Sobreira, N.L., Swaminathan, G.J., Taschner, P.E., Terry, S.F., Washington, N.L., Zuchner, S., Boycott, K.M., Rehm, H.L., 2015. The Matchmaker Exchange: a platform for rare disease gene discovery. Hum Mutat 36, 915–921.

84. Piche, J., Van Vliet, P.P., Puceat, M., Andelfinger, G., 2019. The expanding phenotypes of cohesinopathies: one ring to rule them all! Cell Cycle 18, 2828–2848.

85. Ritchie, M.E., Phipson, B., Wu, D., Hu, Y., Law, C.W., Shi, W., Smyth, G.K., 2015. limma powers differential expression analyses for RNA-sequencing and microarray studies. Nucleic Acids Res 43, e47.

86. Rowley, M.J., Corces, V.G., 2018. Organizational principles of 3D genome architecture. Nat Rev Genet 19, 789–800.

87. Saito, H., Yamamura, K., Suzuki, N., 2012. Reduced bone morphogenetic protein receptor type 1A signaling in neural-crest-derived cells causes facial dysmorphism. Dis Model Mech 5, 948–955.

88. Sakata, T., Tei, S., Izumi, K., Krantz, I.D., Bando, M., Shirahige, K., 2025. A common molecular mechanism underlying Cornelia de Lange and CHOPS syndromes. Curr Biol 35, 1353–1363 e1355.

89. Sanborn, A.L., Rao, S.S., Huang, S.C., Durand, N.C., Huntley, M.H., Jewett, A.I., Bochkov, I.D., Chinnappan, D., Cutkosky, A., Li, J., Geeting, K.P., Gnirke, A., Melnikov, A., McKenna, D., Stamenova, E.K., Lander, E.S., Aiden, E.L., 2015. Chromatin extrusion explains key features of loop and domain formation in wild-type and engineered genomes. Proc Natl Acad Sci U S A 112, E6456–6465.

90. Sheridan, S.D., Theriault, K.M., Reis, S.A., Zhou, F., Madison, J.M., Daheron, L., Loring, J.F., Haggarty, S.J., 2011. Epigenetic characterization of the FMR1 gene and aberrant neurodevelopment in human induced pluripotent stem cell models of fragile X syndrome. PLoS One 6, e26203.

91. Shi, Y., Kirwan, P., Livesey, F.J., 2012. Directed differentiation of human pluripotent stem cells to cerebral cortex neurons and neural networks. Nat Protoc 7, 1836–1846.

92. Silva, M.C.C., Powell, S., Ladstatter, S., Gassler, J., Stocsits, R., Tedeschi, A., Peters, J.M., Tachibana, K., 2020. Wapl releases Scc1-cohesin and regulates chromosome structure and segregation in mouse oocytes. J Cell Biol 219.

93. Singh, T., Poterba, T., Curtis, D., Akil, H., Al Eissa, M., Barchas, J.D., Bass, N., Bigdeli, T.B., Breen, G., Bromet, E.J., Buckley, P.F., Bunney, W.E., Bybjerg-Grauholm, J., Byerley, W.F., Chapman, S.B., Chen, W.J., Churchhouse, C., Craddock, N., Cusick, C.M., DeLisi, L., Dodge, S., Escamilla, M.A., Eskelinen, S., Fanous, A.H., Faraone, S.V., Fiorentino, A., Francioli, L., Gabriel, S.B., Gage, D., Gagliano Taliun, S.A., Ganna, A., Genovese, G., Glahn, D.C., Grove, J., Hall, M.H., Hamalainen, E., Heyne, H.O., Holi, M., Hougaard, D.M., Howrigan, D.P., Huang, H., Hwu, H.G., Kahn, R.S., Kang, H.M., Karczewski, K.J., Kirov, G., Knowles, J.A., Lee, F.S., Lehrer, D.S., Lescai, F., Malaspina, D., Marder, S.R., McCarroll, S.A., McIntosh, A.M., Medeiros, H., Milani, L., Morley, C.P., Morris, D.W., Mortensen, P.B., Myers, R.M., Nordentoft, M., O’Brien, N.L., Olivares, A.M., Ongur, D., Ouwehand, W.H., Palmer, D.S., Paunio, T., Quested, D., Rapaport, M.H., Rees, E., Rollins, B., Satterstrom, F.K., Schatzberg, A., Scolnick, E., Scott, L.J., Sharp, S.I., Sklar, P., Smoller, J.W., Sobell, J.L., Solomonson, M., Stahl, E.A., Stevens, C.R., Suvisaari, J., Tiao, G., Watson, S.J., Watts, N.A., Blackwood, D.H., Borglum, A.D., Cohen, B.M., Corvin, A.P., Esko, T., Freimer, N.B., Glatt, S.J., Hultman, C.M., McQuillin, A., Palotie, A., Pato, C.N., Pato, M.T., Pulver, A.E., St Clair, D., Tsuang, M.T., Vawter, M.P., Walters, J.T., Werge, T.M., Ophoff, R.A., Sullivan, P.F., Owen, M.J., Boehnke, M., O’Donovan, M.C., Neale, B.M., Daly, M.J., 2022. Rare coding variants in ten genes confer substantial risk for schizophrenia. Nature 604, 509–516.

94. Sobreira, N., Schiettecatte, F., Valle, D., Hamosh, A., 2015. GeneMatcher: a matching tool for connecting investigators with an interest in the same gene. Hum Mutat 36, 928–930.

95. Tai, D.J., Ragavendran, A., Manavalan, P., Stortchevoi, A., Seabra, C.M., Erdin, S., Collins, R.L., Blumenthal, I., Chen, X., Shen, Y., Sahin, M., Zhang, C., Lee, C., Gusella, J.F., Talkowski, M.E., 2016. Engineering microdeletions and microduplications by targeting segmental duplications with CRISPR. Nat Neurosci 19, 517–522.

96. Tai, D.J.C., Razaz, P., Erdin, S., Gao, D., Wang, J., Nuttle, X., de Esch, C.E., Collins, R.L., Currall, B.B., O’Keefe, K., Burt, N.D., Yadav, R., Wang, L., Mohajeri, K., Aneichyk, T., Ragavendran, A., Stortchevoi, A., Morini, E., Ma, W., Lucente, D., Hastie, A., Kelleher, R.J., Perlis, R.H., Talkowski, M.E., Gusella, J.F., 2022. Tissue-and cell-type-specific molecular and functional signatures of 16p11.2 reciprocal genomic disorder across mouse brain and human neuronal models. Am J Hum Genet 109, 1789–1813.

97. Tedeschi, A., Wutz, G., Huet, S., Jaritz, M., Wuensche, A., Schirghuber, E., Davidson, I.F., Tang, W., Cisneros, D.A., Bhaskara, V., Nishiyama, T., Vaziri, A., Wutz, A., Ellenberg, J., Peters, J.M., 2013. Wapl is an essential regulator of chromatin structure and chromosome segregation. Nature 501, 564–568.

98. van Ruiten, M.S., van Gent, D., Sedeno Cacciatore, A., Fauster, A., Willems, L., Hekkelman, M.L., Hoekman, L., Altelaar, M., Haarhuis, J.H.I., Brummelkamp, T.R., de Wit, E., Rowland, B.D., 2022. The cohesin acetylation cycle controls chromatin loop length through a PDS5A brake mechanism. Nat Struct Mol Biol 29, 586–591.

99. Waizenegger, I.C., Hauf, S., Meinke, A., Peters, J.M., 2000. Two distinct pathways remove mammalian cohesin from chromosome arms in prophase and from centromeres in anaphase. Cell 103, 399–410.

100. Wang, L., Wang, S., Li, W., 2012. RSeQC: quality control of RNA-seq experiments. Bioinformatics 28, 2184–2185.

101. Weiss, F.D., Calderon, L., Wang, Y.F., Georgieva, R., Guo, Y., Cvetesic, N., Kaur, M., Dharmalingam, G., Krantz, I.D., Lenhard, B., Fisher, A.G., Merkenschlager, M., 2021. Neuronal genes deregulated in Cornelia de Lange Syndrome respond to removal and re-expression of cohesin. Nat Commun 12, 2919.

102. Wendt, K.S., Yoshida, K., Itoh, T., Bando, M., Koch, B., Schirghuber, E., Tsutsumi, S., Nagae, G., Ishihara, K., Mishiro, T., Yahata, K., Imamoto, F., Aburatani, H., Nakao, M., Imamoto, N., Maeshima, K., Shirahige, K., Peters, J.M., 2008. Cohesin mediates transcriptional insulation by CCCTC-binding factor. Nature 451, 796–801.

103. Wutz, G., Varnai, C., Nagasaka, K., Cisneros, D.A., Stocsits, R.R., Tang, W., Schoenfelder, S., Jessberger, G., Muhar, M., Hossain, M.J., Walther, N., Koch, B., Kueblbeck, M., Ellenberg, J., Zuber, J., Fraser, P., Peters, J.M., 2017. Topologically associating domains and chromatin loops depend on cohesin and are regulated by CTCF, WAPL, and PDS5 proteins. EMBO J 36, 3573–3599.

104. Yan, J., Enge, M., Whitington, T., Dave, K., Liu, J., Sur, I., Schmierer, B., Jolma, A., Kivioja, T., Taipale, M., Taipale, J., 2013. Transcription factor binding in human cells occurs in dense clusters formed around cohesin anchor sites. Cell 154, 801–813.

105. Yan, J., Zhang, F., Brundage, E., Scheuerle, A., Lanpher, B., Erickson, R.P., Powis, Z., Robinson, H.B., Trapane, P.L., Stachiw-Hietpas, D., Keppler-Noreuil, K.M., Lalani, S.R., Sahoo, T., Chinault, A.C., Patel, A., Cheung, S.W., Lupski, J.R., 2009. Genomic duplication resulting in increased copy number of genes encoding the sister chromatid cohesion complex conveys clinical consequences distinct from Cornelia de Lange. J Med Genet 46, 626–634.

106. Yuan, B., Neira, J., Pehlivan, D., Santiago-Sim, T., Song, X., Rosenfeld, J., Posey, J.E., Patel, V., Jin, W., Adam, M.P., Baple, E.L., Dean, J., Fong, C.T., Hickey, S.E., Hudgins, L., Leon, E., Madan-Khetarpal, S., Rawlins, L., Rustad, C.F., Stray-Pedersen, A., Tveten, K., Wenger, O., Diaz, J., Jenkins, L., Martin, L., McGuire, M., Pietryga, M., Ramsdell, L., Slattery, L., Study, D.D.D., Abid, F., Bertuch, A.A., Grange, D., Immken, L., Schaaf, C.P., Van Esch, H., Bi, W., Cheung, S.W., Breman, A.M., Smith, J.L., Shaw, C., Crosby, A.H., Eng, C., Yang, Y., Lupski, J.R., Xiao, R., Liu, P., 2019. Clinical exome sequencing reveals locus heterogeneity and phenotypic variability of cohesinopathies. Genet Med 21, 663–675.

107. Zhang, N., Coutinho, L.E., Pati, D., 2021. PDS5A and PDS5B in Cohesin Function and Human Disease. Int J Mol Sci 22.

108. Zhang, Y., Pak, C., Han, Y., Ahlenius, H., Zhang, Z., Chanda, S., Marro, S., Patzke, C., Acuna, C., Covy, J., Xu, W., Yang, N., Danko, T., Chen, L., Wernig, M., Sudhof, T.C., 2013. Rapid single-step induction of functional neurons from human pluripotent stem cells. Neuron 78, 785–798.

109. Zhou, W., Triche, T.J., Jr., Laird, P.W., Shen, H., 2018. SeSAMe: reducing artifactual detection of DNA methylation by Infinium BeadChips in genomic deletions. Nucleic Acids Res 46, e123.

