## Supplemental Figures for "Clinical, *in vitro,* and *in vivo* evidence of *WAPL* as a novel cohesinopathy gene and phenotypic driver of 10q22.3q23.2 genomic disorder"

**List of Supplemental Tables**

Table S1. Constraint metrics of human cohesin genes

Table S2. Cohort-based analyses of disease association of cohesin release factor genes

Table S3. Databases queried to identify WAPL, PDS5A, PDS5B, and 10q22q23 cases

Table S4. *WAPL*, *PDS5A*, and *PDS5B* cases details

Table S5. 10q22q23 known disease genes and del-associated phenotypes

Table S6. Cases with DNA methylation analysis

Table S7. Cell line genotypes

Table S8. DEG statistics

Table S9. DEGs

Table S10. DEGs within and flanking 10q dels and dups

Table S11. Analyses of heart function in wt and *Wapl*$^{+/-}$ heterozygotes

Table S12. Guide RNAs, primers, ddPCR assay, antibodies

Table S13. CNVs overlapping 10q GD region

### Supplemental Figures

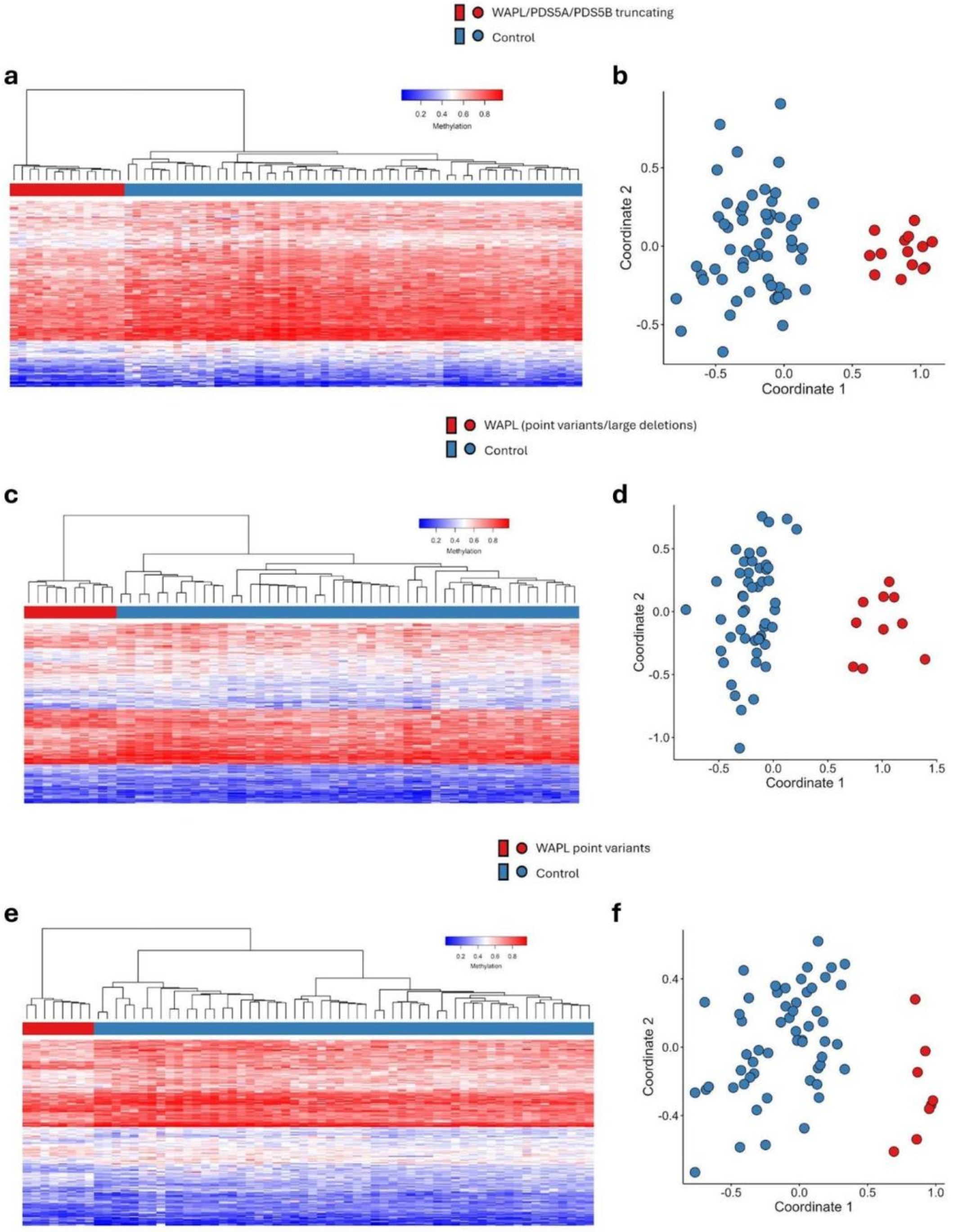


**Figure S1. Verification of the methylation episignature probes. a.** Hierarchical clustering, based on the probes selected in Analysis 1, with rows of the heatmap representing probes and columns representing samples. The heatmap shows methylation levels, ranging from blue (0, no methylation) to red (1, full methylation). Case and control samples are shown in red and blue, respectively, in the bar above the heatmap. **b.** Multidimensional scaling (MDS) plot for Analysis 1, using the same case/control color coding as in panel (a). **c, d.** Hierarchical clustering heatmap (c) and MDS plot (d) for Analysis. **e, f.** Hierarchical clustering heatmap (e) and MDS plot (f) for Analysis 3. In all three analyses, the heatmap and MDS plot demonstrates a clear distinction between case and control samples, supporting the reliability of the selected probes.

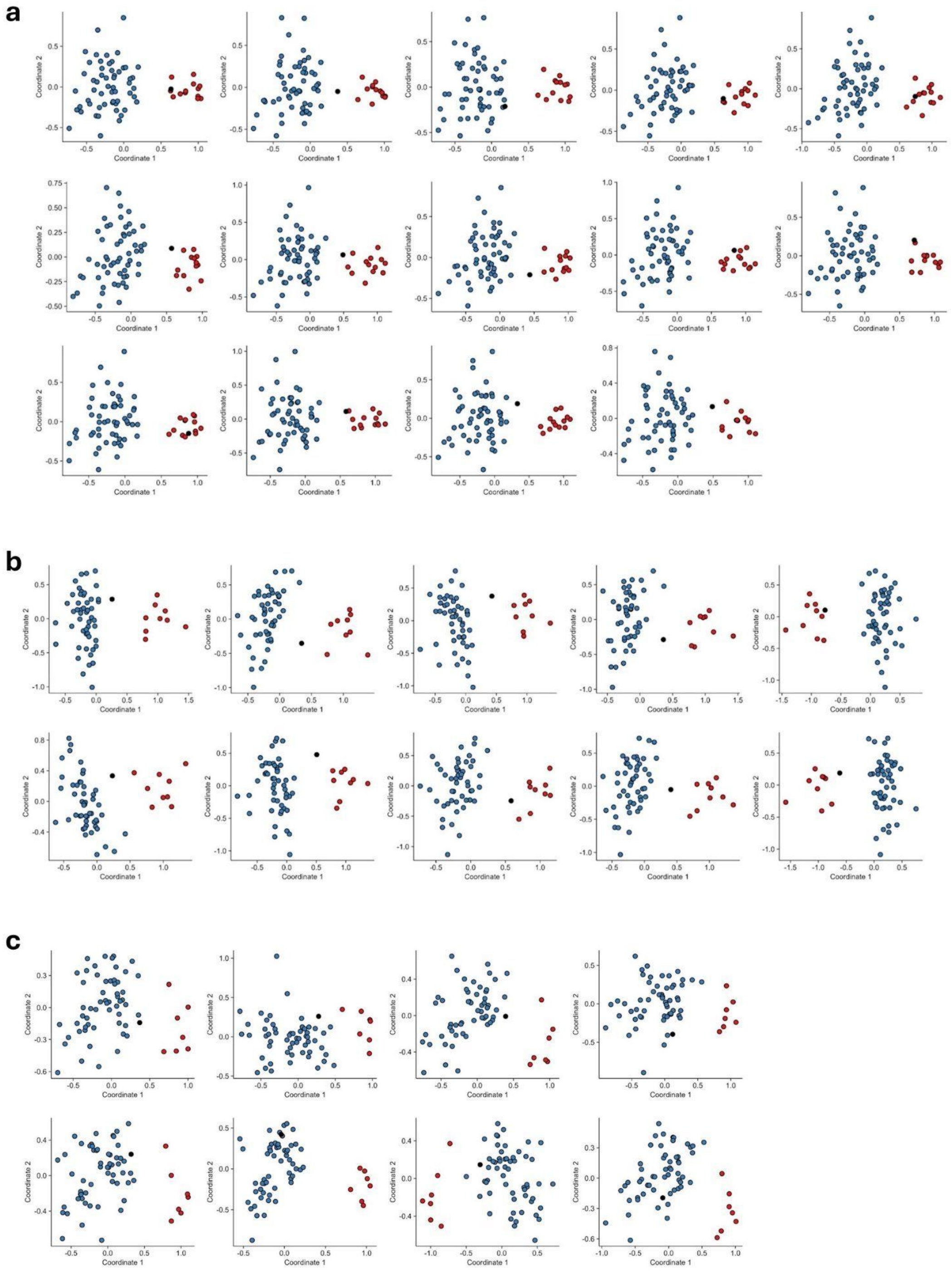


**Figure S2. Leave-one-out cross-validation (LOOCV) of methylation analyses. a-c.** MDS plots generated to assess whether the left-out sample (black) clusters with the other case samples (red) versus control samples (blue). The left-out case sample clustered with the other case samples in almost all iterations of Analysis 1 (a), for the majority of iterations of Analysis 2 (b), but never in Analysis 3 (c).

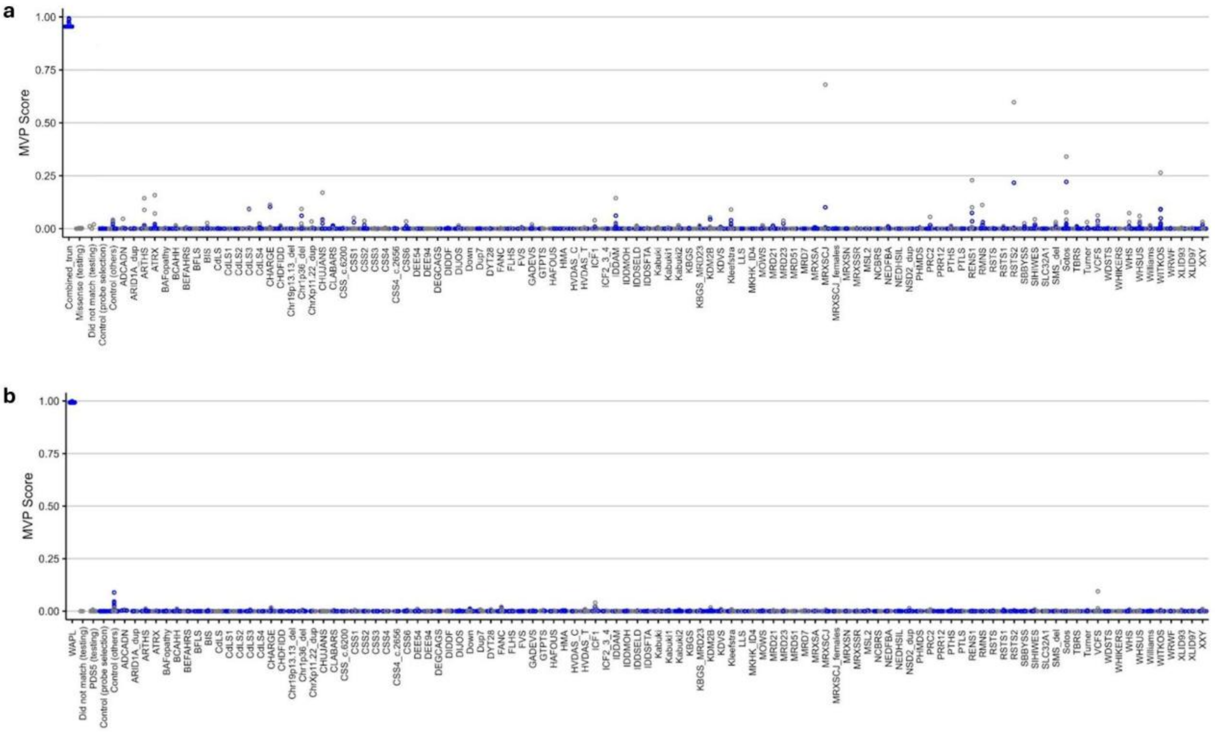


**Figure S3. Methylation variant pathogenicity scores. a.** Visualization of the MVP scores produced by the SVM for Analysis 1. Blue circles indicate training samples, while grey circles represent testing samples. **b.** MVP scores corresponding to Analysis 2.

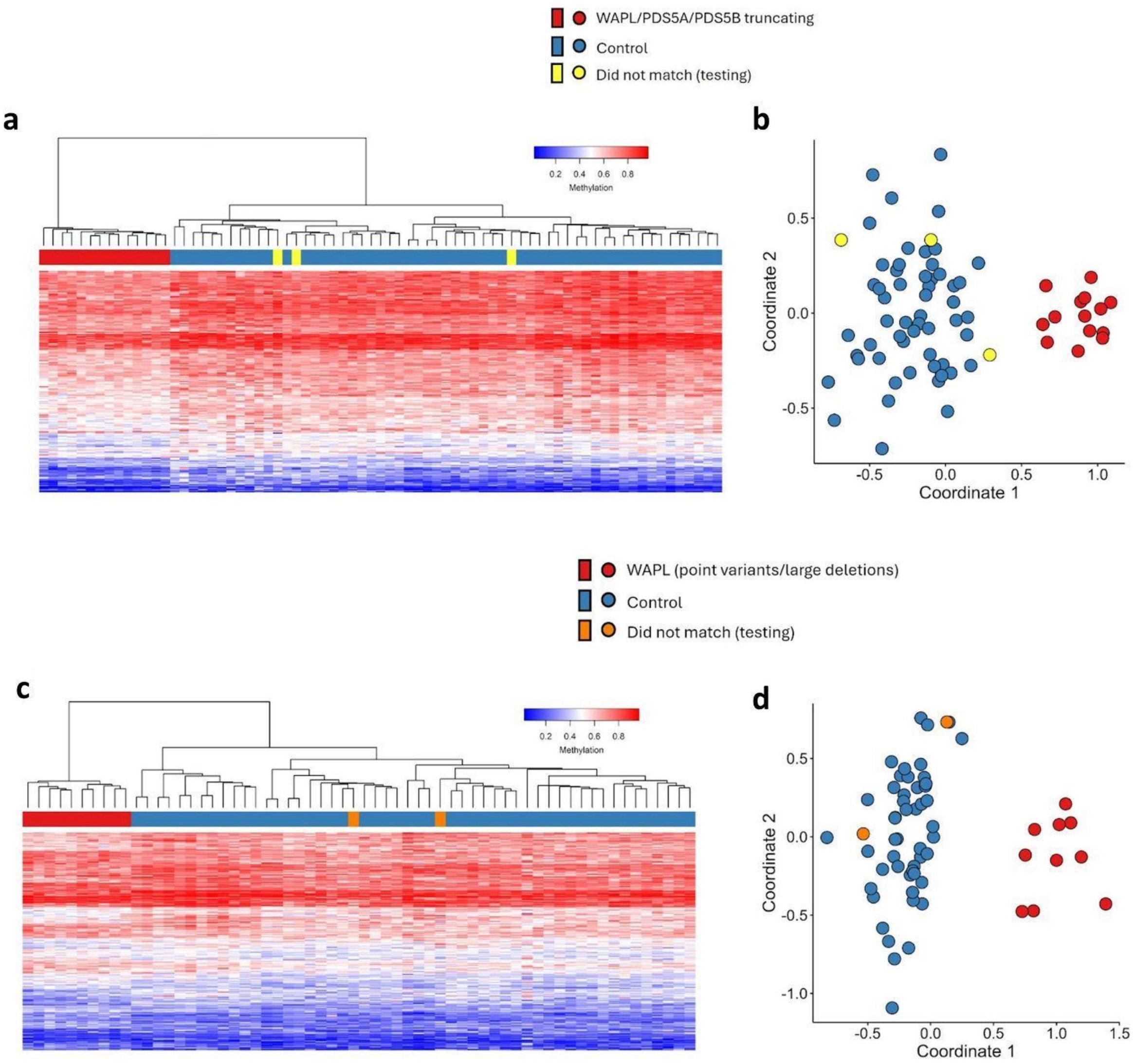


**Figure S4. Methylation episignature, testing additional study samples using the identified episignatures.** Test study samples were plotted along with the case and control samples used for probe selection. **a, b.** Hierarchical clustering and MDS plots showing the results of testing samples with truncating variants that were not used for probe selection or model training in Analysis 1. **c, d.** Testing of samples with *WAPL* point variants and large dels encompassing *WAPL* that were not included in probe selection or model construction in Analysis 2. In neither Analysis do the additional (test) samples plot with the original case samples.

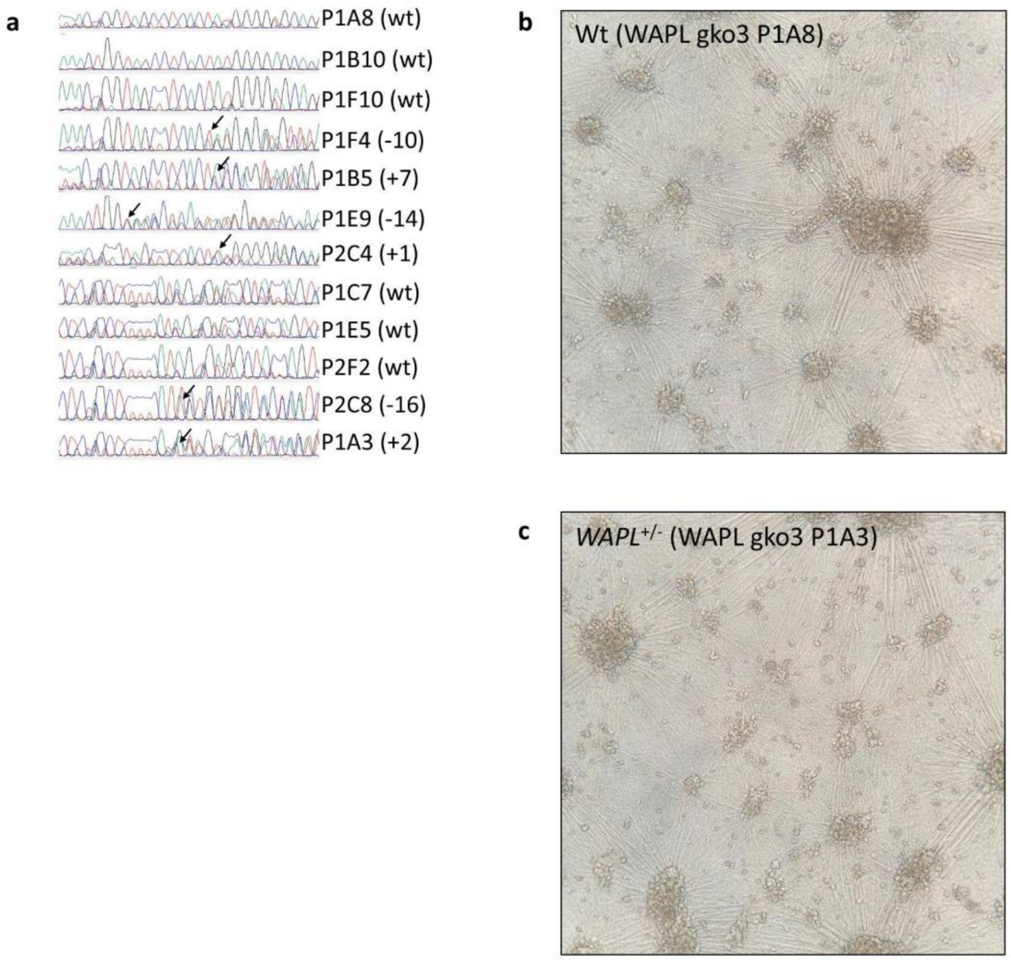


**Figure S5. CRISPR cell line genotyping and morphology. a.** Sanger sequencing of *WAPL* gko3 clones. Arrow indicates position of heterozygous fs indels. The number of bases added or deleted is indicated in parentheses. Genotyping of 10q del and dup lines by ddPCR is not shown. **b-c.** Example morphology of day 24 induced neurons (iNs).

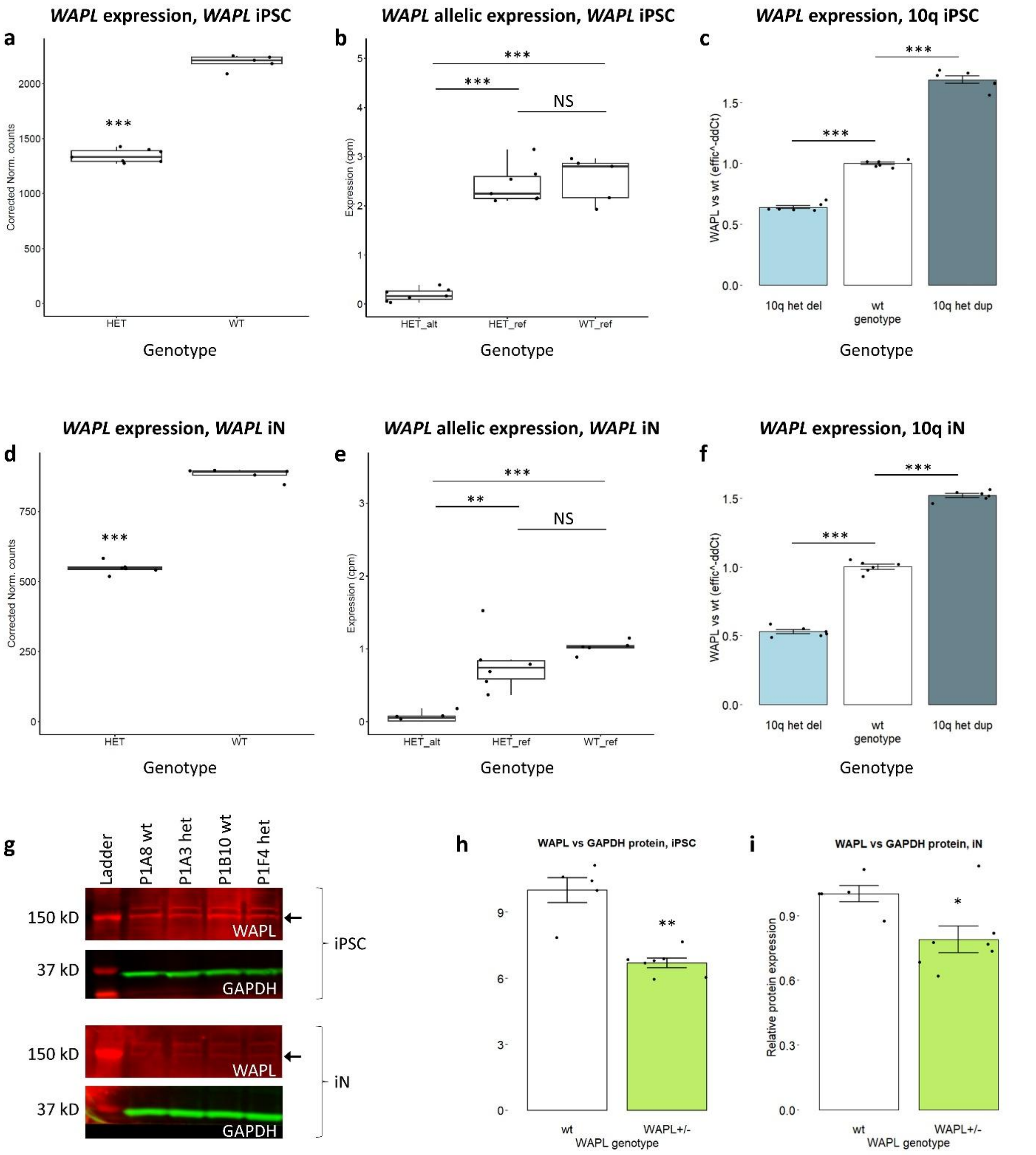


**Figure S6.** ***WAPL*** **expression is expectedly dosage-responsive in** $\boldsymbol{WAPL^{+/-}}$**, het 10q del, and het 10q dup CRISPR-edited cell lines.** All data are from RNA-seq. **a-c.** iPSCs. **d-f.** iNs. **a, d.** *WAPL* expression (SVA-corrected normalized read counts) in $WAPL^{+/-}$ and wt iPSCs (a) (p = 2.37e-65) and iNs (d) (p = 5.88e-26). Counts are via DESeq2. Statistics are via two-tailed t-test. **b, e.** Normalized allelic expression of *WAPL*, by genotype and allele measured. Expression of the reference (ref) allele in control samples was divided by two before plotting (Methods). The alternate (alt) allele in het lines is diminished compared to the ref allele in het lines (iPSCs p = 1.03e-6, iNs p = 5.72e-3) and compared to the ref allele in wt lines (iPSCs p = 2.16e-4, iNs p = 1.97e-7). The ref allele in het lines is not differentially expressed compared to the ref allele in wt lines, in either cell type (p=6.55e-1 in iPSCs and p=2.23e-1 in iNs). This demonstrates that the overall decrease of *WAPL* in *WAPL*$^{+/-}$ lines is attributable to a decrease in the alternate (alt) allele. Statistics are via two-tailed t test. **c, f.** qPCR of *WAPL* in 10q del, wt, and 10q dup iPSCs and iNs, normalized to wt lines. Control gene is *POLR2A*. Statistics are via two-tailed t-test. **g.** Example Western blots, staining for WAPL and normalization protein GAPDH, on protein lysates from iPSCs (top) and iNs (bottom). The lower molecular weight band (arrow, ~150 kD) for WAPL was quantitated. WAPL expression is lower in neurons than in iPSCs. **h-i.** Quantitation of Western blot results of WAPL protein normalized to GAPDH. Each data point represents the average of technical duplicates of one clonal line. N = 5 wt and 7 het fs. **h.** Quantitation of Western blots. iPSCs show a decrease of 33% (p=2.51e-3 by t-test) **i.** iNs show a decrease of 21% (p=1.69e-2 by t-test). * $p<0.05$, ** $p<0.005$, *** $p<0.0005$.

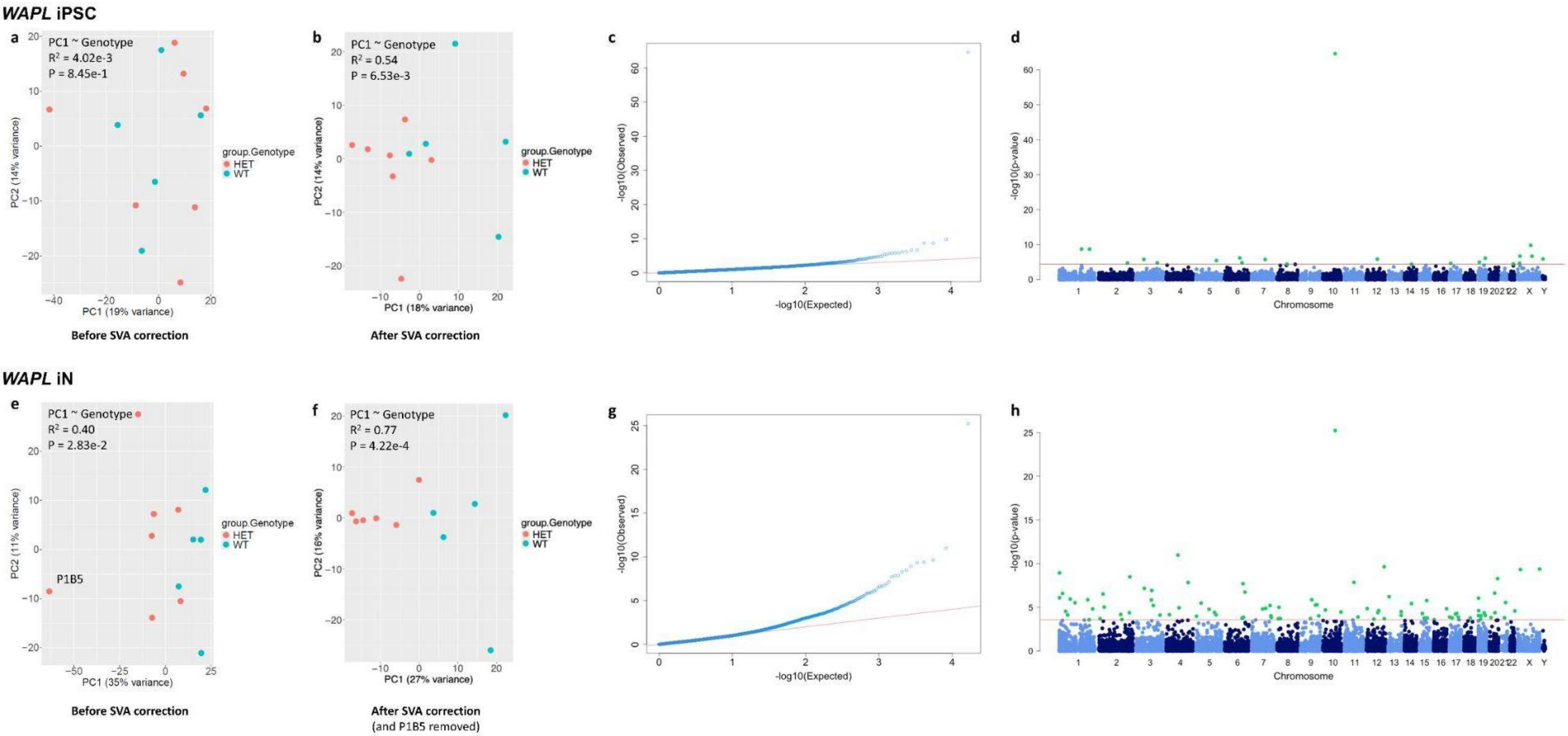


**Figure S7. RNA-Seq QC, *WAPL*. a-d.** *WAPL* iPSC. **e-h.** *WAPL* iN. **a, e.** Principal components analysis (PCA) plot by genotype, before correction by surrogate variable analysis (SVA). **b, f.** PCA plot by genotype, after correction by SVA. **c, g.** q-q plots. **d, h.** Manhattan plots of differentially expressed genes.

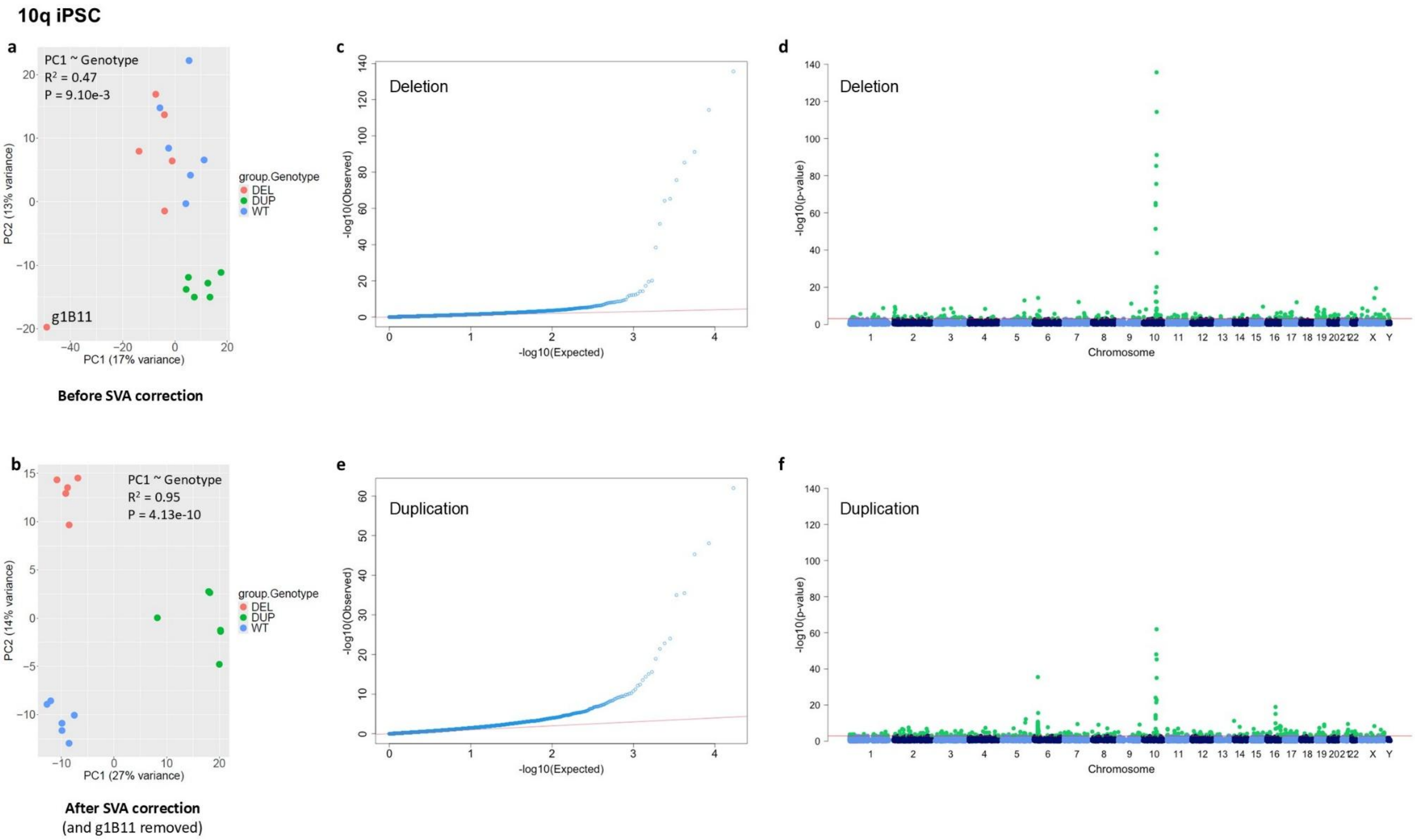


**Figure S8. RNA-Seq QC, 10q iPSC. a.** Principal components analysis (PCA) plot by genotype, before correction by surrogate variable analysis (SVA). **b.** PCA plot by genotype, after correction by SVA. **c, e.** q-q plots of del and dup, respectively. **d, f.** Manhattan plots of differentially expressed genes in del and dup, respectively.

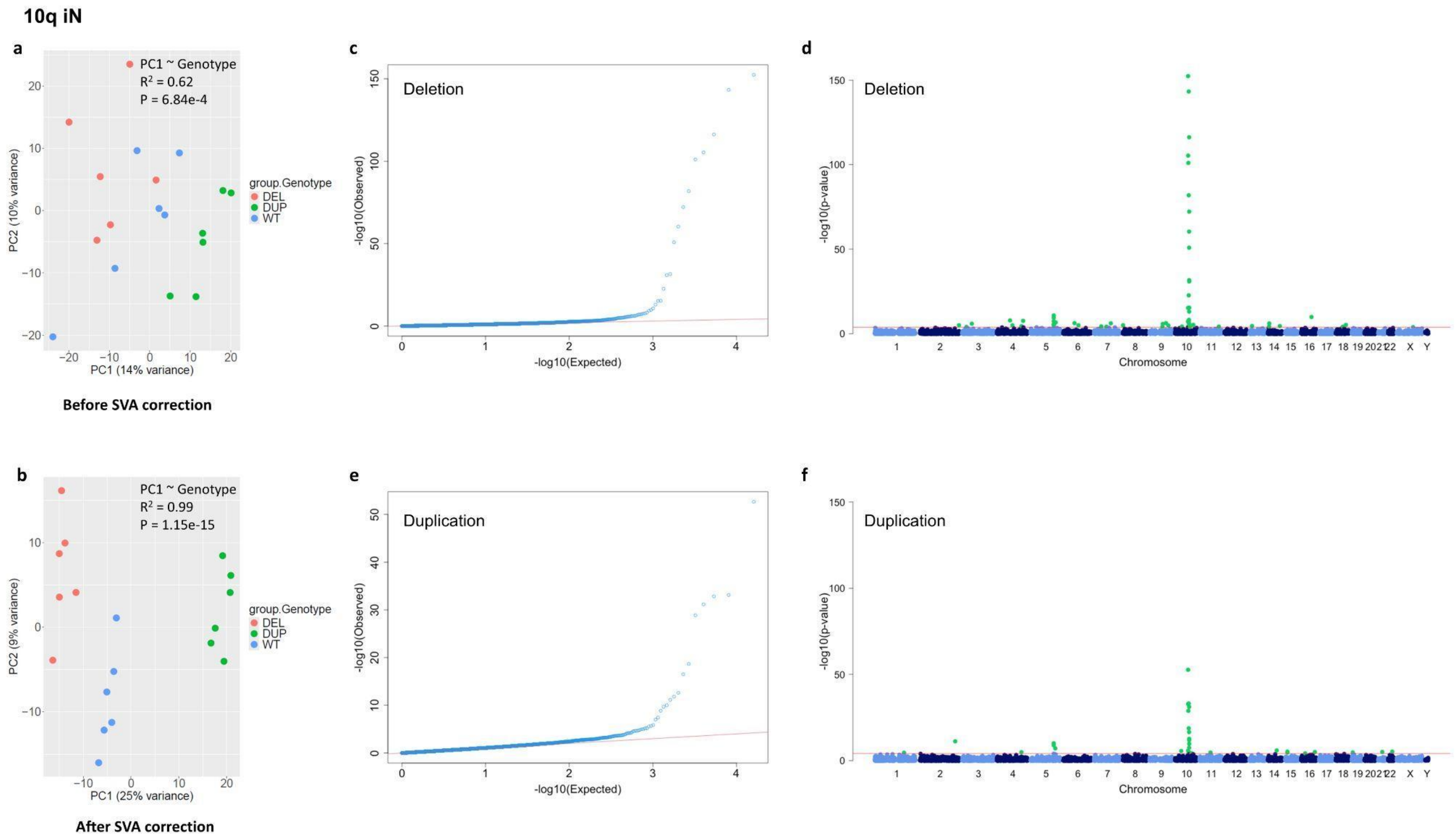


**Figure S9. RNA-Seq QC, 10q iN. a.** Principal components analysis (PCA) plot by genotype, before correction by surrogate variable analysis (SVA). **b.** PCA plot by genotype, after correction by SVA. **c, e.** q-q plots of del and dup, respectively. **d, f.** Manhattan plots of differentially expressed genes of del and dup, respectively.

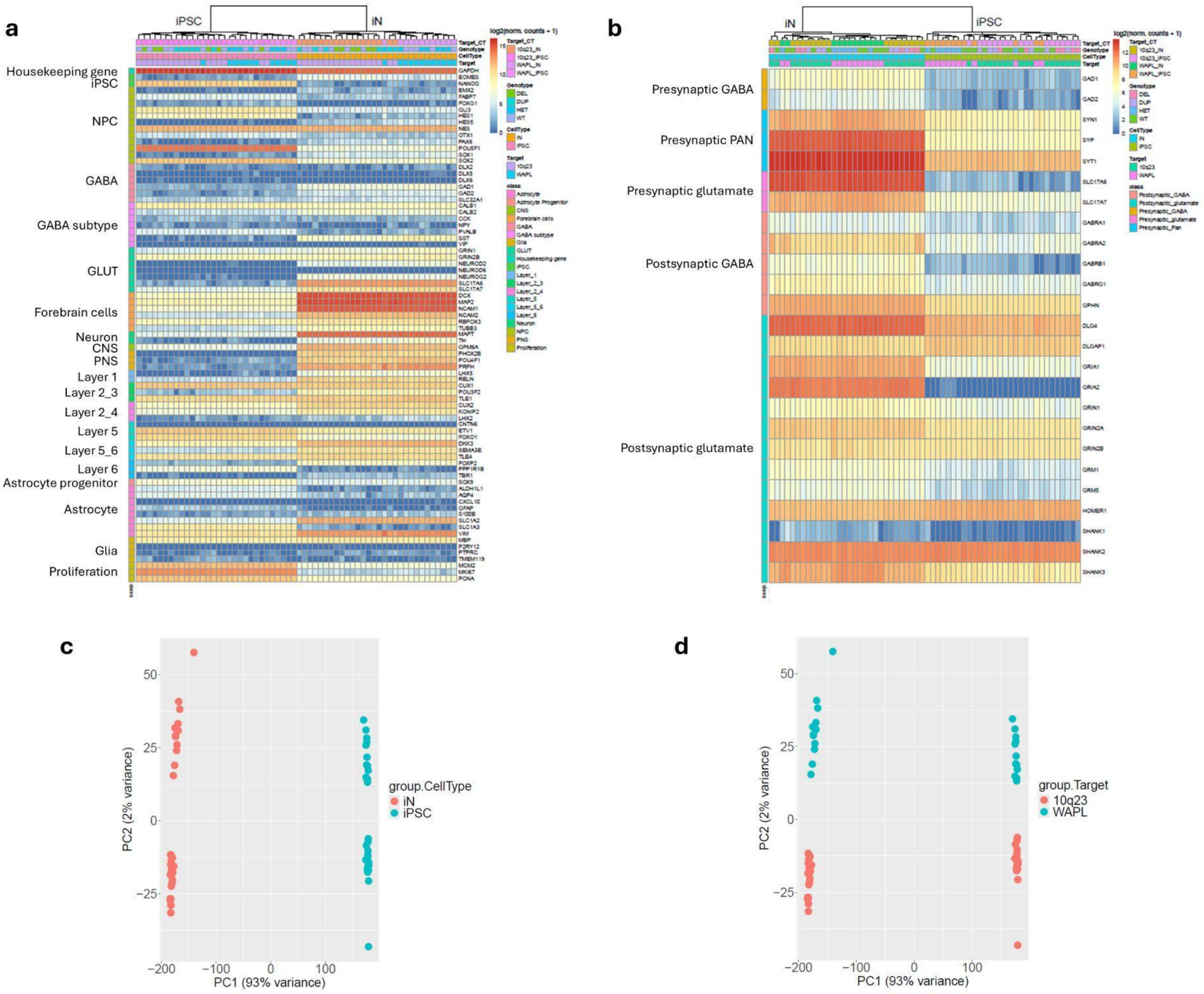


**Figure S10. RNA-Seq QC, cell type identity as identified by gene expression. a-b.** Heat maps reveal that iPSCs are comparatively enriched for stemness and proliferation genes, while iNs are comparatively enriched for neuronal genes, particularly forebrain. **c.** Principal components analysis (PCA) separates by cell type to an extreme degree. **d.** By PCA, the 10q experiment's lines (del, dup, wt) are separated from the *WAPL* experiment's lines (del and wt).

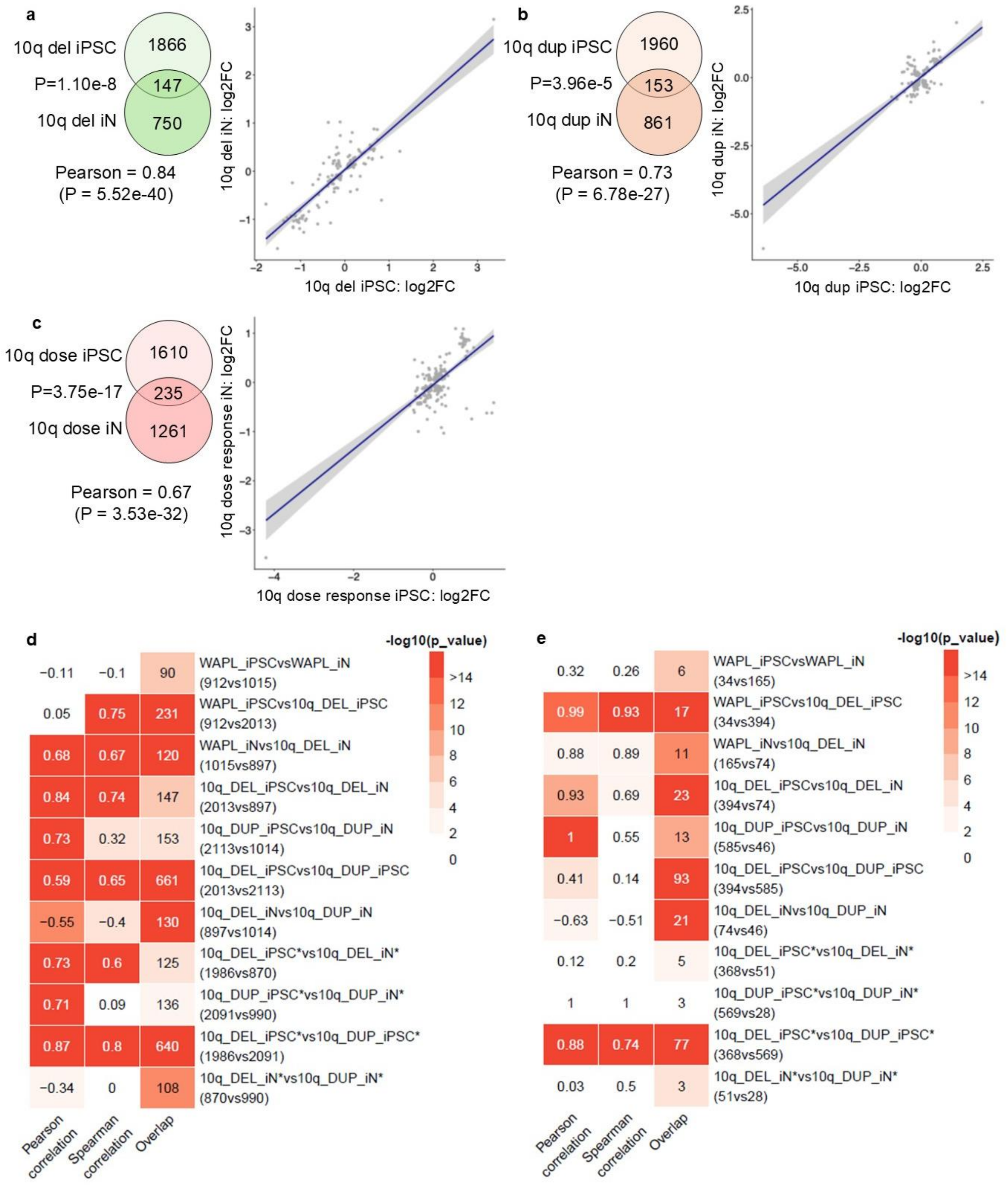


**Figure S11. Supplement to Fig. 4. a-c.** Similar plots to Fig. 4b-d, respectively, but with genes within the 10q deleted or duplicated region retained. dose = dose responsive. **c-d.** Heatmaps of DEG overlaps between the stated cell lines, from DEGs at the P<0.05 level (c) or FDR <0.1 level (d). * indicates that the DEG set has 10q22.3q23.2 region genes removed.

**Figure S12. *PDS5B* subject photographs.** Case-level photographs are unable to be displayed, per medRxiv regulations.

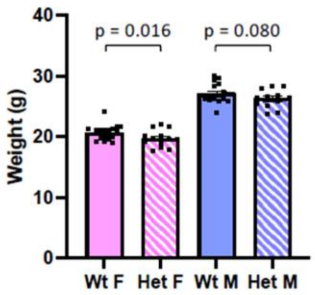


**Figure S13. Adult *Wapl*$^{+/-}$ mice are smaller than their wt cohorts.** Mice were weighed on days 120-134. *Wapl*$^{+/-}$ females and males are 4.6% and 4.3% smaller, respectively. Bar graphs depict mean + standard error of the mean. P values were calculated using Student's t-test. N = 72: 27 wt females; 19 het females; 15 wt males; 11 het males.

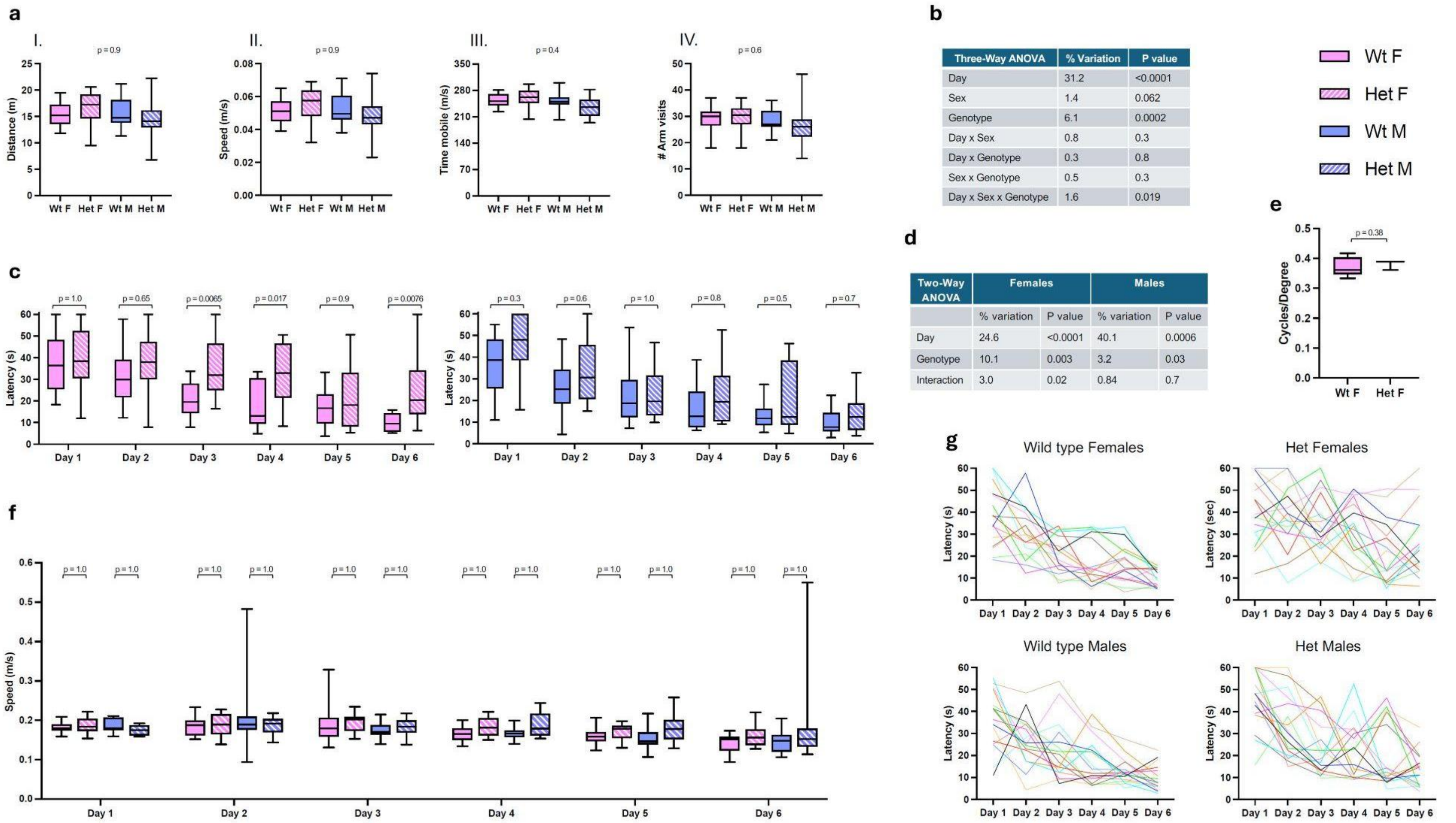


| Three-Way ANOVA | % Variation | P value |
|---|---|---|
| Day | 31.2 | <0.0001 |
| Sex | 1.4 | 0.062 |
| Genotype | 6.1 | 0.0002 |
| Day x Sex | 0.8 | 0.3 |
| Day x Genotype | 0.3 | 0.8 |
| Sex x Genotype | 0.5 | 0.3 |
| Day x Sex x Genotype | 1.6 | 0.019 |

| Two-Way ANOVA | Females | | Males | |
|---|---|---|---|---|
| | % variation | P value | % variation | P value |
| Day | 24.6 | <0.0001 | 40.1 | 0.0006 |
| Genotype | 10.1 | 0.003 | 3.2 | 0.03 |
| Interaction | 3.0 | 0.02 | 0.84 | 0.7 |

**Figure S14. Supplement to adult mouse behavior figure (Fig. 6). a.** Supplement for Fig. 6b (Y-Maze). Genotype does not significantly affect movement in the Y-Maze. i. Total distance traveled. ii. Average speed. iii. Time mobile. iv. Number of arm visits. In all panels, data are presented as median, the 25-75% quartiles, and max/min values. Statistical significance of genotype was computed using 2-way ANOVA. N = 64 (16 adults per group). **b-g.** Supplement for Fig. 6c (Morris Water Maze, MWM). **b.** Summary of 3-way ANOVA analysis of MWM data presented in Fig. 6c. N = 64, with 16 mice in each group. **c-d.** Poor performance in the MWM in female heterozygotes. Times to reach the hidden platform (latency) are analyzed separately for each sex by 2-way ANOVA and post-hoc Sidak's multiple comparison test to determine pairwise comparisons. Note that genotype explains a larger part of the variation in female mice. **e.** Diminished visual acuity does not explain MWM performance deficiencies in heterozygous mice. Visual acuity was assessed using OptoDrum. Data are presented as median, the 25-75% quartiles, and max/min values. Statistical significance was computed using a Student's t-test. N = 26 (15 wt and 11 *Wapl*$^{+/-}$ females). **f.** Impaired swimming abilities do not explain deficient MWM performances of heterozygous mice. Average daily swim speeds for each group analyzed in Fig. 6c. Data were analyzed by 3-way ANOVA followed by post-hoc Sidak's test to determine pairwise comparisons. **g.** Performances of individual mice in the MWM. Each colored line represents the behavior of one mouse over the 6-day testing period. These data emphasize the highly variable performance of heterozygous mice, especially heterozygous females.

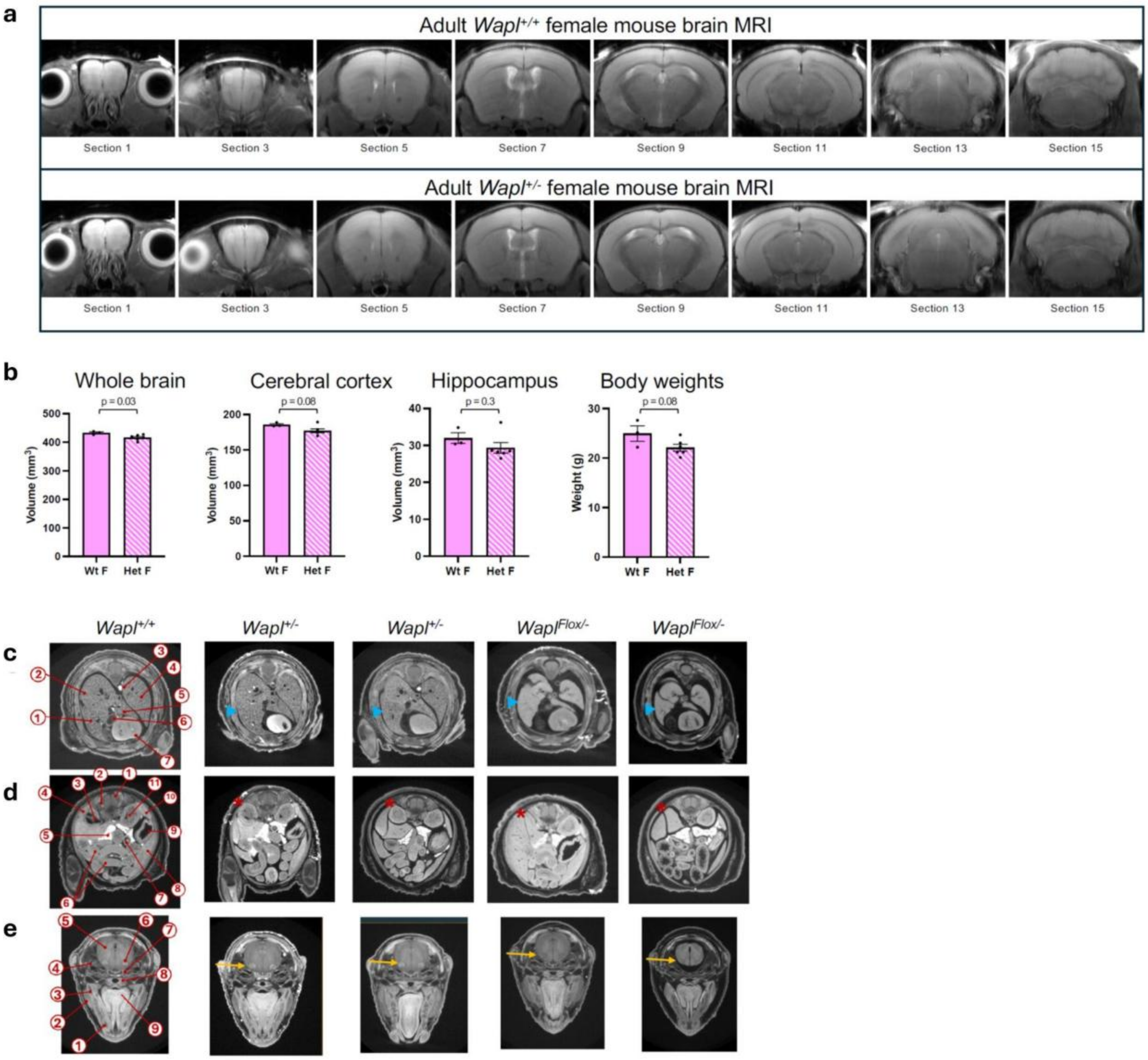


**Figure S15. Mouse anatomic phenotyping. a-b.** Whole brain, cerebral cortex and hippocampal morphologies and volumes. **a.** Representative MRI images of one wt and one adult *Wapl*$^{+/-}$ mouse. For each animal, 16 sections were collected and 8 of these are depicted here. We could not distinguish *Wapl*$^{+/+}$ and *Wapl*$^{+/-}$ samples. **b.** Referencing the Allen Brain Atlas, images were segmented for whole brain, cerebral cortex, and hippocampus and then volumes calculated. The small decreases in brain volumes are consistent with the decreased body mass in heterozygotes. N = 9 (3 wt and 6 *Wapl*$^{+/-}$ females; age 8-9 mo). **c-e.** Mouse embryo micro-CT reveals a dosage liability threshold for birth defects between 25% and 50% of wt *Wapl* level. Embryonic development in *Wapl*$^{+/-}$ (N = 7) and in *Wapl*$^{\mathrm{Flox}/-}$ (N = 6) e18.5 embryos was assessed by micro-CT scanning. Representative transverse sections are displayed. *Wapl*$^{+/-}$ embryos (50% Wapl levels) are small but at the appropriate developmental stage and no dysmorphologies were identified. *Wapl*$^{\mathrm{Flox}/-}$ embryos (25% Wapl levels) are small. Overall, *Wapl*$^{\mathrm{Flox}/-}$ embryos are at the appropriate developmental stage, however, three discrete phenotypes were observed: (c) 5 of 6 *Wapl*$^{\mathrm{Flox}/-}$ embryos displayed small lungs and immature alveolar development (blue arrowhead) (p = 0.005), (d) 3 of 6 *Wapl*$^{\mathrm{Flox}/-}$ embryos displayed missing or a severely reduced right kidney (red asterisks) (p = 0.07); and (e) 3 of 6 embryos displayed absent or a severely reduced facial motor nucleus (yellow arrows) (p = 0.07). **c.** 1, right lung middle lobe; 2, right lung cranial lobe; 3, dorsal thoracic aorta; 4, lung left lobe; 5, right lung accessory lobe; 6, superior vena cava; 7, heart. **d.** 1, spinal cord; 2, sympathetic ganglia; 3, right kidney pelvis; 4, right kidney cortex; 5, pancreas; 6, small intestine; 7, hindgut; 8, liver; 9, stomach; 10, spleen; 11, left kidney medulla. **e.** 1, incisor; 2, masseter muscle; 3, mandible ramus; 4, exoccipital bone with stapes; 5, medulla oblongata; 6, facial motor nucleus; 7, basosphenoid bone; 8, pituitary with Rathke's pouch; 9, tongue. P values were calculated using Fisher's exact test.

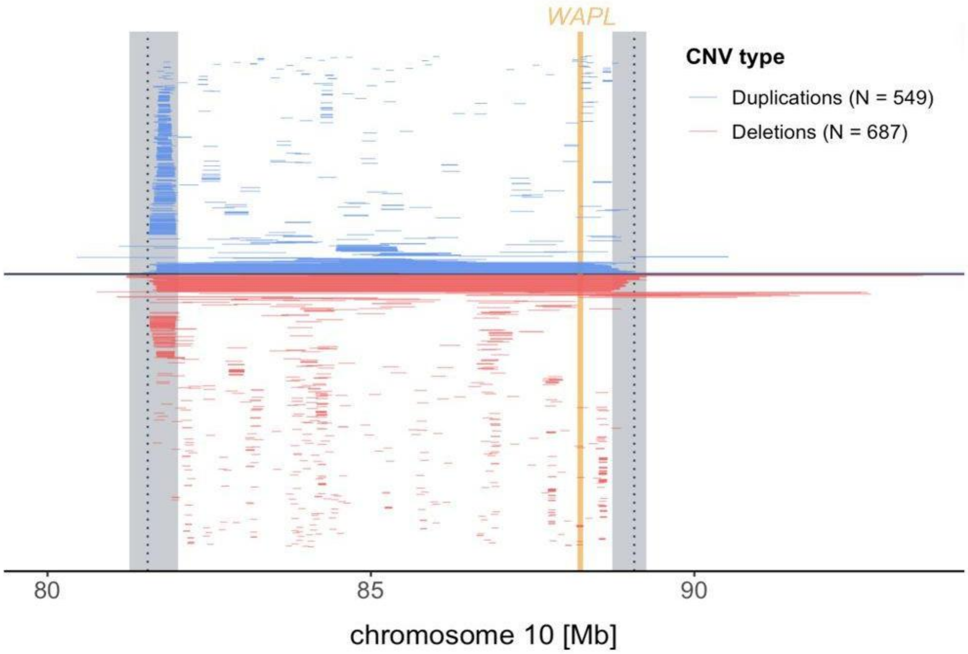


**Figure S16. All rCNVs with any overlap of the 10q22.3q23.2 GD region.** Flanking segmental duplications are in grey. CRISPR breakpoints are indicated by dotted lines.

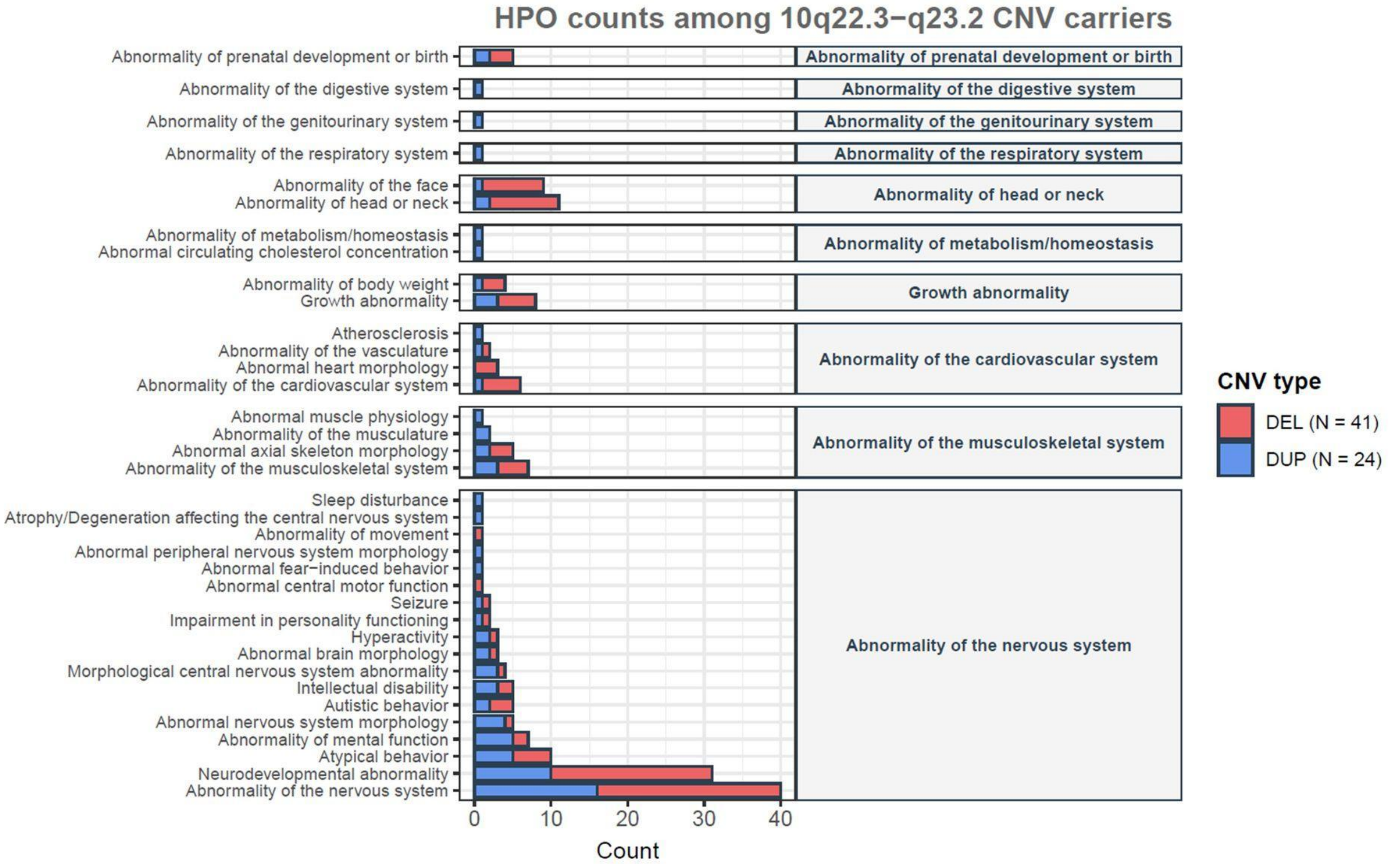


**Figure S17. Phenotypes of carriers of recurrent 10q22.3q23.2 deletions and duplications.** HPO ‘metaphenotypes’ are counted for individuals with CNVs with at least 80% overlap of the recurrent 10q22.3q23.2 reciprocal genomic disorder region. These include 39 affected deletion carriers, two unaffected deletion carriers, 18 affected duplication carriers, and six unaffected duplication carriers. Metaphenotypes are derived from clustering of HPO terms to derive ≥ 3rd degree ancestor terms (see Methods).
